# Comparison of the Efficacy and Safety of Myasthenia Gravis Treatments: A Bayesian Network Meta-Analysis

**DOI:** 10.64898/2025.12.04.25341653

**Authors:** Nilay P. McLaren, Matteo Rosati, Wayne Zhong, Sheena Hussain, Melissa C. Funaro, Richard J. Nowak, Bhaskar Roy

## Abstract

**Importance:** Many novel targeted therapies have shown promise in treating generalized myasthenia gravis (MG); however, no head-to-head prospective studies have compared efficacy and safety between one another, nor to existing therapies.

**Objective:** We conducted a nuanced comparative analysis of MG treatments that accounts for differences in trial designs and populations.

**Data Sources and Study Selection:** We searched Scopus, Embase, Web of Science, CENTRAL, and MEDLINE up to May 2, 2025, for randomized trials for MG. We included prospective multi-arm studies of adults (18 or older) with generalized MG that reported mean treatment difference, with uncertainty, in MG-Activities of Daily Living (MG-ADL) or Quantitative MG (QMG) score change from baseline. Of 4,823 eligible studies, 32 studies (27 placebo-controlled) met these criteria.

**Data Extraction and Synthesis:** Two independent reviewers extracted data in accordance with PRISMA guidelines and assessed risk of bias. Data was pooled with a Bayesian random-effects model.

**Main Outcomes and Measures:** The primary outcome was the difference in MG-ADL or QMG (patient-reported and physician-reported outcome measures, respectively) change from baseline between treatment arms.

**Results:** Twenty-seven trials (placebo n=1,086; treatment n=1,232) were included that targeted B cells, neonatal Fc receptors (FcRn), complement activity, CD40, and interleukin-6 signaling as well as immunoglobulin G and broad immunosuppression therapy. QMG scores decreased by an average of -3.6 (95% Credible Interval: [-4.40, -2.72]) points more in patients treated with FcRn inhibitors than those treated with placebo and standard of care, followed by -2.59 [-3.93, -1.27] points for C5 complement inhibitors (C5i), and -2.50 [-5.15, 0.13] points for CD19+ B-cell depletion therapy (BCDT). Sensitivity analyses detected potential confounding of treatment effects by age, sex, and baseline MG-ADL and QMG scores. Therefore, differences in trial populations may mask the treatment effect of CD20+ BCDT. Patients treated with FcRn inhibitors had greater odds of treatment-related adverse events (Odds Ratio: 2.20 [1.52, 3.38]) while C5i and CD19+ BCDT showed comparable odds to placebo and standard of care.

**Conclusion and Relevance:** FcRn inhibitors, C5i, and CD19+ BCDT have comparable efficacies in treating MG. However, our findings highlight how differences between trials may confound the interpretation of study outcomes, warranting further prospective comparative studies.

**Key Points:** **Question:** How do the different therapies for myasthenia gravis (MG) compare in terms of their efficacy and safety?

**Findings:** In this Bayesian network meta-analysis, we find comparable efficacies between C5 complement inhibitors, FcRn inhibitors, and CD19+ B-cell depletion therapy. We also demonstrate how confounders can mask or amplify treatment effects.

**Meaning:** This study serves as a cautionary tale against uncontextualized comparisons of treatment efficacy and warrants prospective studies that are better suited for determining optimal MG treatment strategies.

## Introduction

Myasthenia gravis (MG) is an autoimmune disorder mediated by autoantibodies against antigens at the neuromuscular junction. These autoantibodies disrupt synaptic transmission and consequently lead to muscle weakness. About 75-80% of patients with MG have autoantibodies against the acetylcholine receptor (AChR), and approximately 5-10% have autoantibodies against muscle specific kinase (MuSK).^1,2^ Even fewer may harbor autoantibodies against low-density lipoprotein receptor-related protein 4 (LRP4) or agrin, and some may have no detectable autoantibodies (classified as seronegative).^2,3^ Historically, acetylcholinesterase inhibitors (pyridostigmine), acute immunomodulatory therapies (intravenous immunoglobulin [IVIg] and plasma exchange [PLEX]), corticosteroids (prednisone), non-steroidal immunosuppressive therapies (IST; azathioprine, mycophenolate mofetil, methotrexate, cyclosporine, and tacrolimus), and thymectomy have been used to treat MG. More recently, targeted therapies against C5 complement activation (eculizumab, ravulizumab, and zilucoplan), B cells (rituximab, belimumab, and inebiluzumab), T cells (iscalimab), neonatal Fc receptor (FcRn; efgartigimod, rozanolixizumab, nipocalimab, and batoclimab), Interleukin-6 (IL-6) signaling (satralizumab and tocilizumab), and B-cell maturation antigen (BCMA; Chimeric Antigen Receptor [CAR] T-cell therapies) have been developed, with some now approved by the FDA for the treatment of generalized MG.^4,5^

Despite the wide range of treatment options, no prospective studies have compared their efficacy and safety. As a viable alternative, researchers have conducted both frequentist and Bayesian meta-analyses, but these studies have limited clinical relevance because they do not account for differences in trial designs and populations.^6–12^ For example, some clinical trials for MG included only AChR+ patients, while others included a mix of AChR+, MuSK+, LRP4+, and seronegative patients.^13–19^ These patients respond to treatment differently and existing meta-analyses have not accounted for this.^2,6–12^

Furthermore, consider how the year of a study can act as a confounder to mask or amplify a treatment effect. Older trials may demonstrate a greater treatment effect than newer trials because the standard of care has improved over time, thus reducing the difference in outcome measure reduction between treatment and placebo arms. Thus, study-wide variables can confound treatment effects, even in randomized trials that balance covariates across study arms.^20^ Therefore, valid comparisons of treatment effects require a thorough analysis of the effects of confounders. There is an unmet need to assess the relative efficacy and safety of MG treatments, and this paper provides such a nuanced comparison by accounting for differences in trial designs and populations.

## Methods

### Screening and Inclusion/Exclusion Criteria

We performed a systematic search of Scopus, Embase, Web of Science, CENTRAL, and MEDLINE up to May 2, 2025 for randomized trials for MG (**e**Table 1). Search results were uploaded to Covidence to remove duplicates and for screening. Two reviewers (NM & MR) performed two rounds of review, independently. In the first, we screened abstracts and titles for relevance (n=2474 irrelevant studies excluded). In the second, we conducted a full text review for studies that met the following inclusion criteria: 1) prospective multi-arm studies, 2) adults (18 or older) with generalized MG, and 3) reported mean treatment difference, with uncertainty (SE, SD, or confidence interval), in MG-ADL or QMG change from baseline. In both rounds, a third reviewer (BR) was consulted to resolve conflicts. Overall, 32 studies met these criteria, 27 of which were placebo-controlled randomized trials. Preferred Reporting Items for Systematic reviews and Meta-Analyses (PRISMA) flow chart of the screening process is shown in **eFigure 1**. This study was prospectively registered in PROSPERO and reported according to PRISMA-NMA guidelines.^21,22^

**Figure 1.**
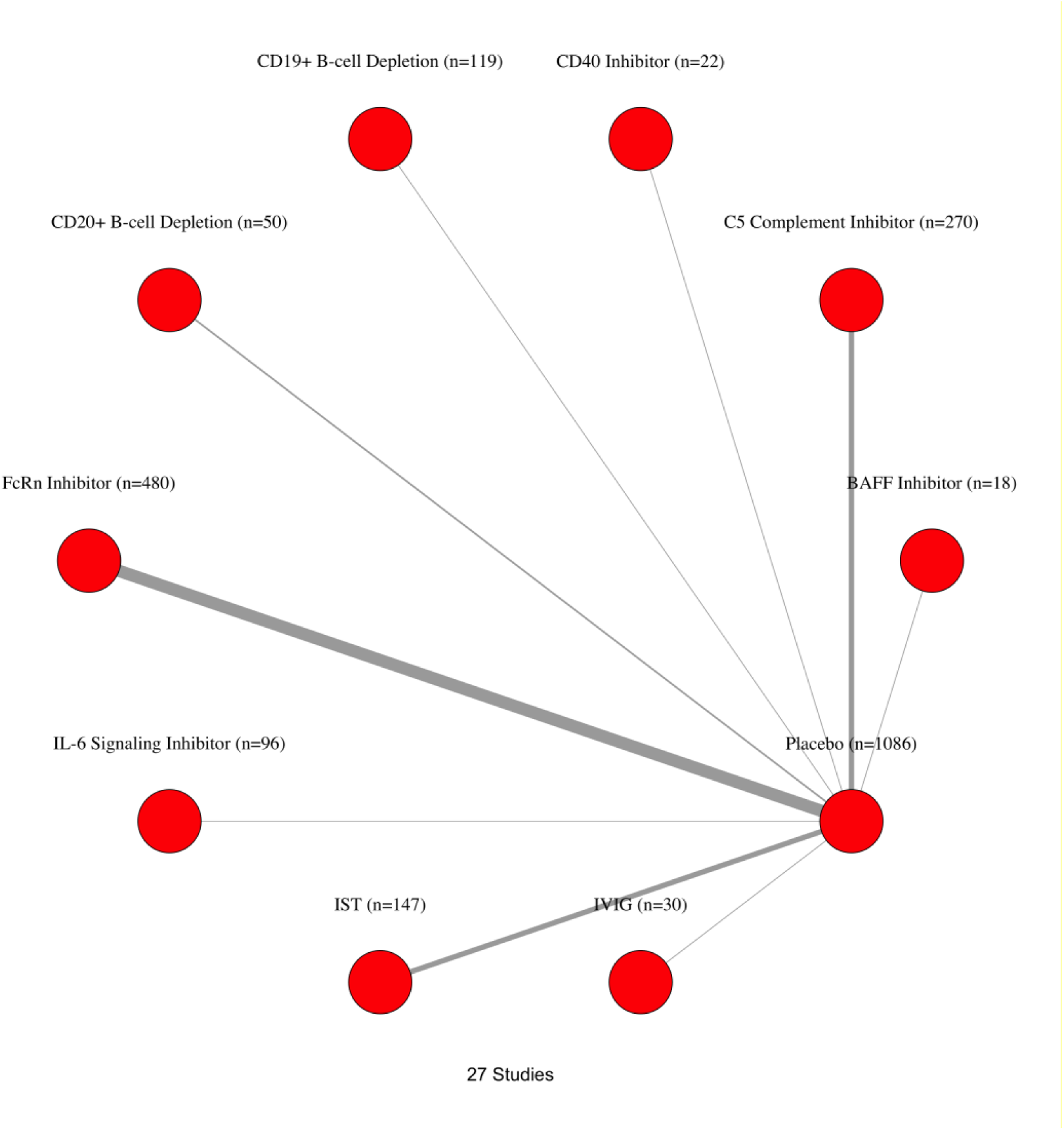
Network plot of included placebo-controlled studies grouped by mechanism of action.

**Table 1.**
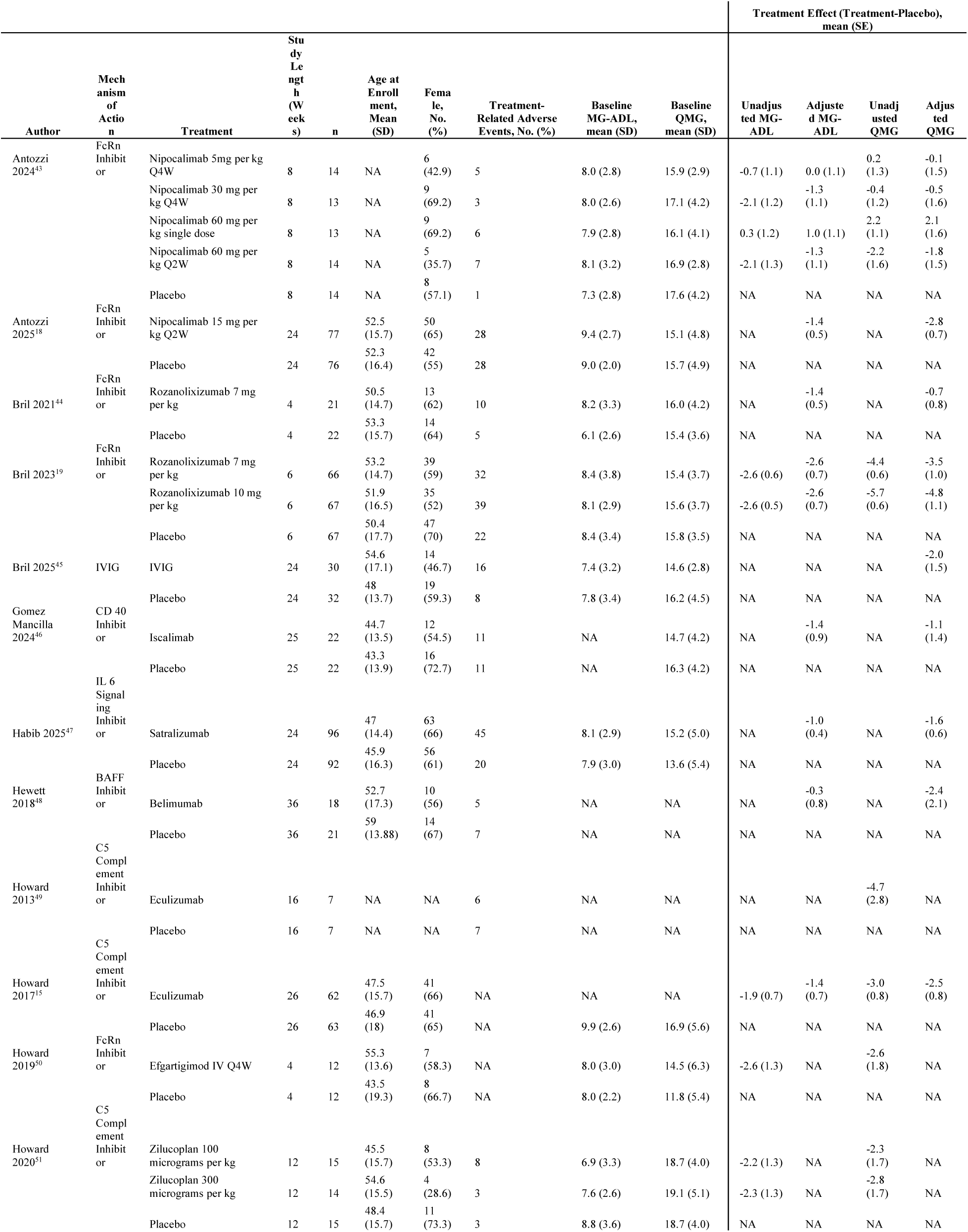
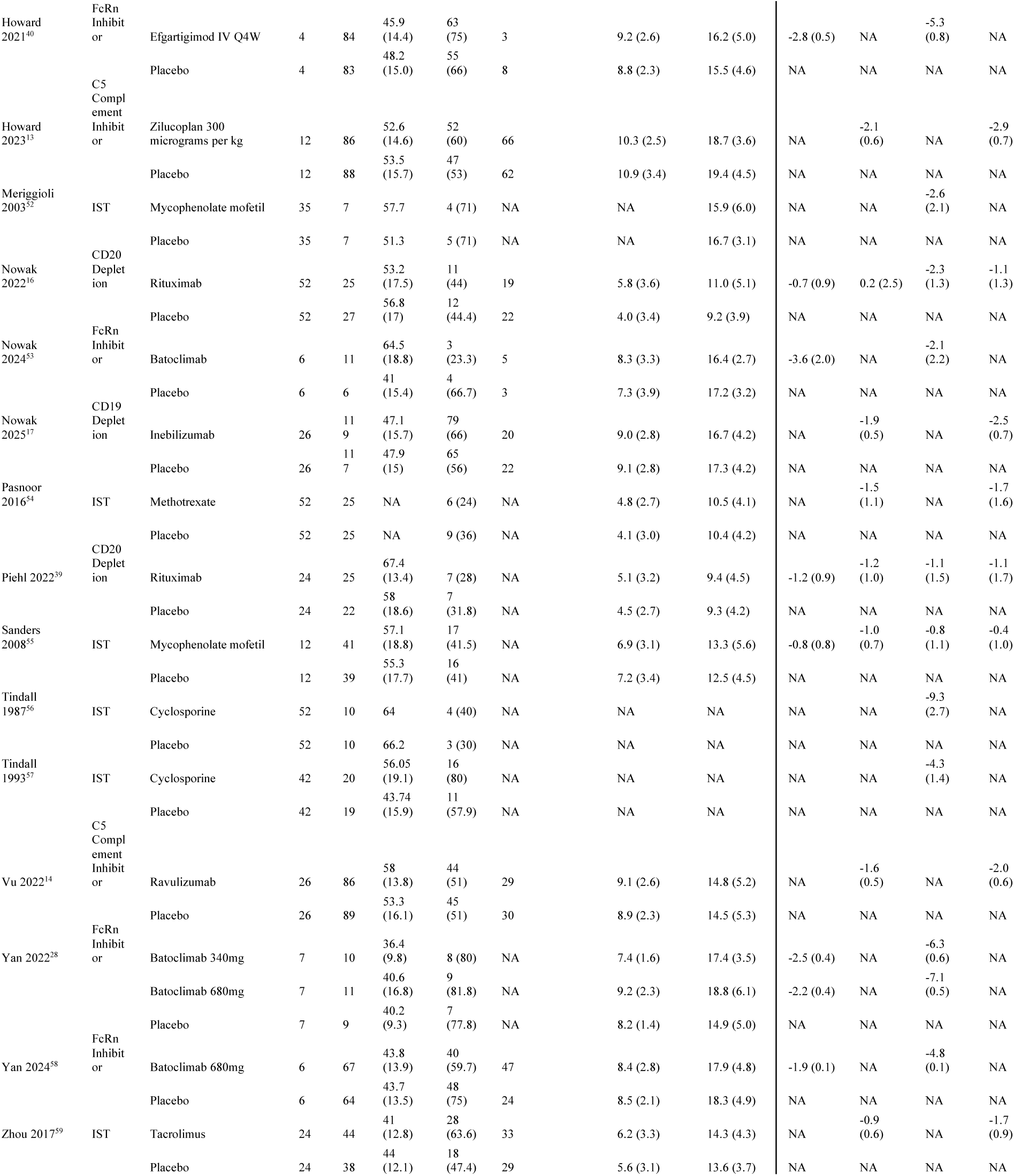
Summary statistics of the 27 included placebo-controlled trials.

### Risk of Bias

We used the revised Cochrane risk of bias 2 tool to assess the quality of the randomized trials. Two independent reviewers (WZ & SH) assessed risk of bias, and a third reviewer (NM) resolved conflicts. Results are displayed in **eFigures 2** **&** 3.

**Figure 2.**
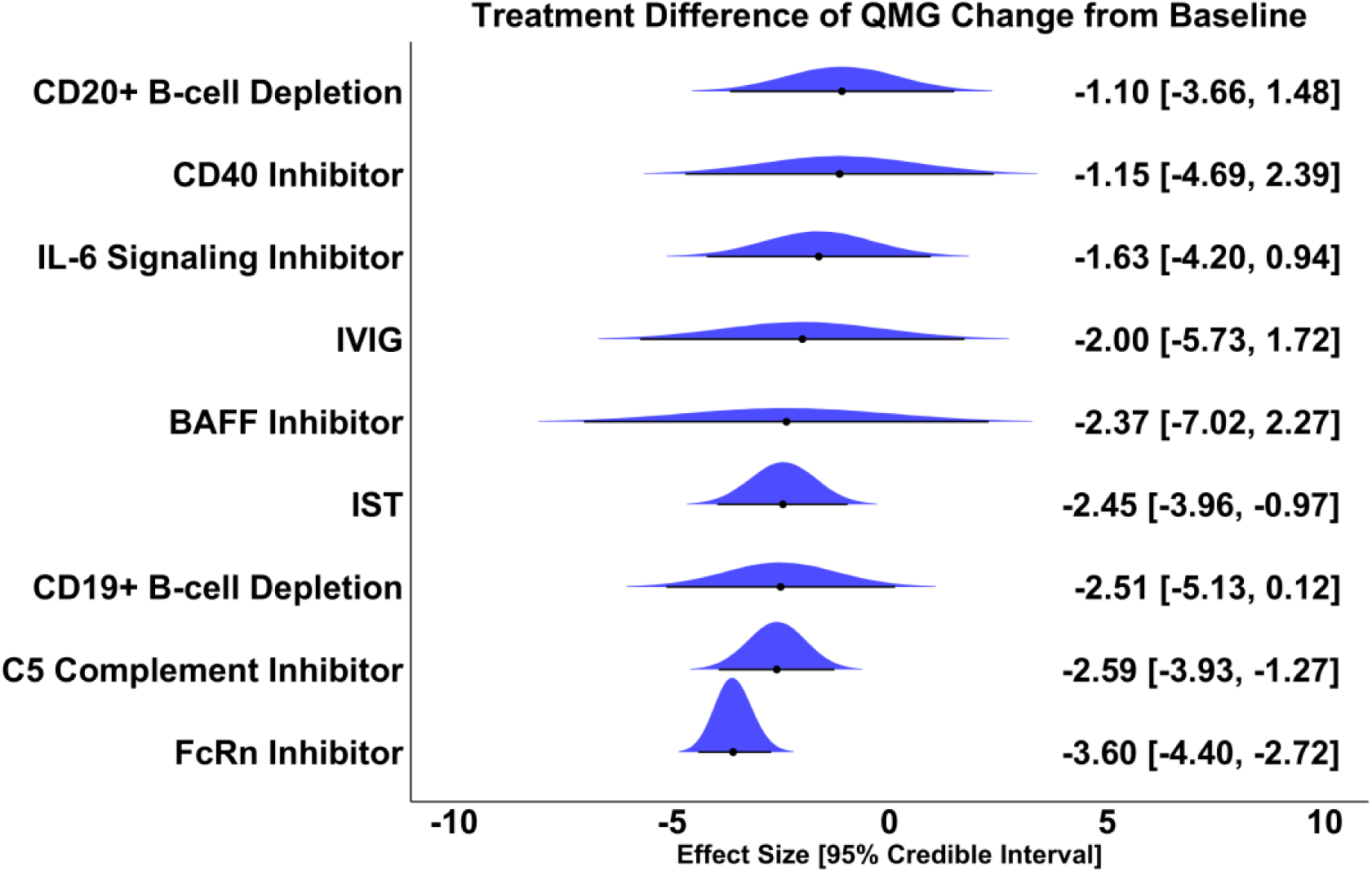
Effect size, 95% Credible Interval, and overlayed posterior distribution of QMG change from baseline compared to placebo. A negative effect size of x is interpreted as: the treatment reduces QMG x points more than placebo and standard of care.

**Figure 3.**
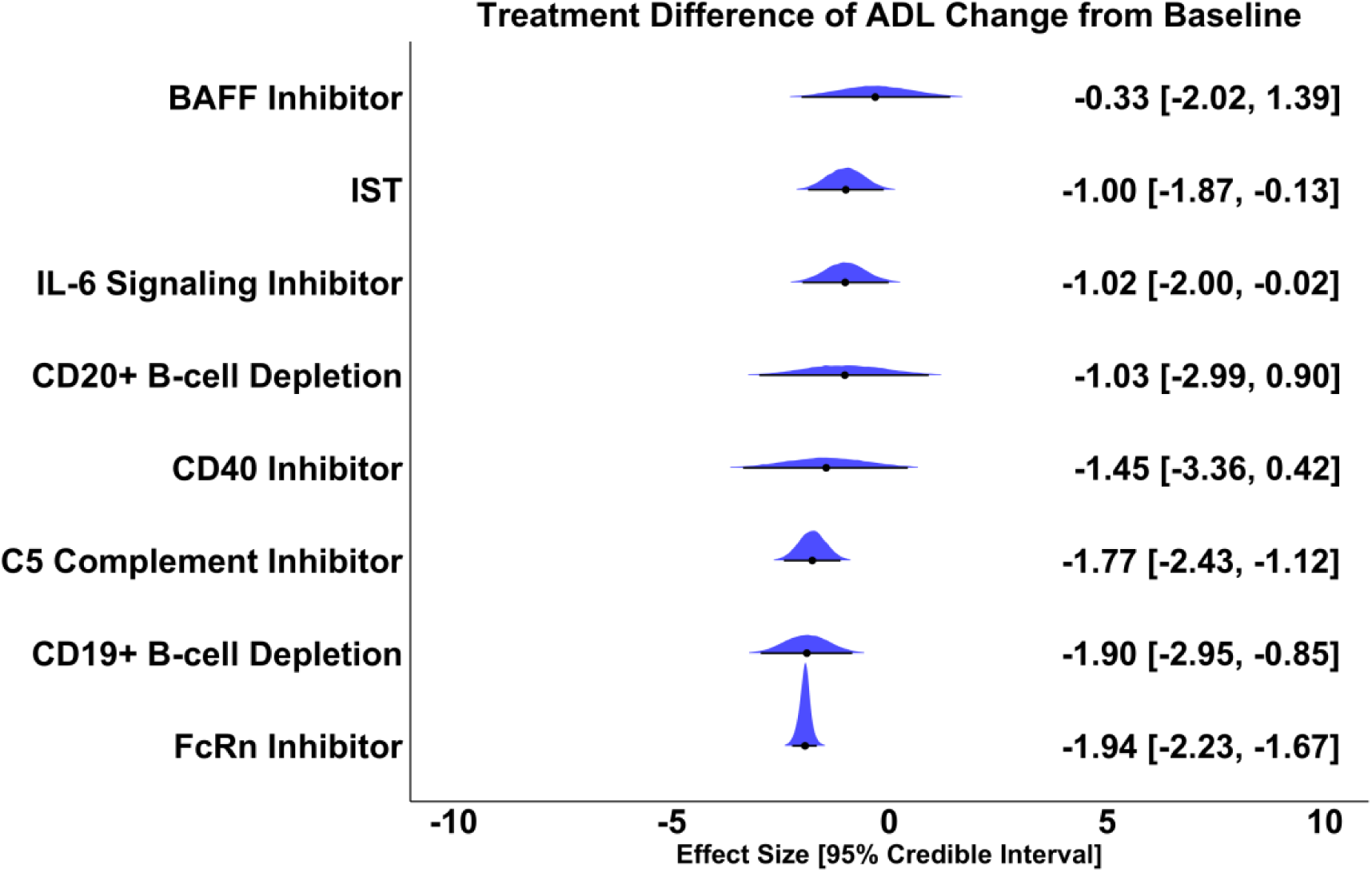
Effect size, 95% Credible Interval, and overlayed posterior distribution of MG-ADL change from baseline compared to placebo. A negative effect size of x is interpreted as: the treatment reduces MG-ADL x points more than placebo and standard of care.

### Statistical Analysis

The following data was collected from the placebo-controlled studies that met the inclusion criteria: mean and standard error of difference in Quantitative MG (QMG) and MG-Activities of Daily Living (MG-ADL) score change from baseline between treatment and placebo arms, length of study, enrollment age, percentage of female study participants, baseline MG-ADL and QMG score, and number of treatment-related adverse events (AE). For studies that reported only unadjusted means and standard errors of individual arms instead of treatment differences, we used the data from individual arms to calculate treatment differences and standard errors. For studies that reported both unadjusted and adjusted treatment differences, we used the adjusted treatment differences if available. For studies that did not explicitly state MG-ADL or QMG change from baseline but instead presented it in a figure, we extracted the relevant data using an online tool (graphreader).^23^ We conducted a Bayesian network meta-analysis using the ‘gemtc’ package in R to estimate treatment effects according to the following hierarchical structure:^24^

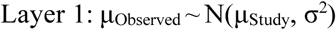

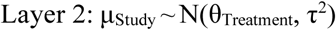

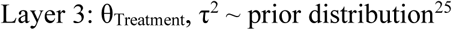

The continuous outcomes (QMG and MG-ADL change from baseline treatment difference) and binary outcome (AE) were modeled using random effects, and parameters were estimated using Markov Chain Monte Carlo (MCMC). We placed an informative log-normal (µ = -2.13, σ^2^ = 1.58^2^) prior on heterogeneity in the AE model. This value was empirically determined from a study of 14,886 meta-analyses that examined the distribution of heterogeneity in meta-analyses of placebo-controlled trials that analyzed subjective binary outcomes (such as AE).^26^ We placed a weakly informative normal (µ = 0, σ^2^ = 10^2^) prior on the heterogeneity for these models. Model convergence was assessed using the Potential Scale Reduction Factor (PSRF), with PSRF <1.05 indicating convergence.

The effect sizes of continuous outcomes (QMG and MG-ADL) are reported as mean difference and 95% Credible Interval (CrI) while the effect sizes of the binary outcome (AE) are reported as an odds ratio and 95% CrI. To further compare treatments, we performed large-scale (∼100,000) simulations in which treatment effects are sampled from their respective posterior distributions to rank treatments based on Surface Under the Cumulative Ranking (SUCRA) scores. These are interpreted as the probability that each treatment has the greatest effect on patients.^27^

We then performed a sensitivity analysis adjusting for the following study-wide covariates: average age at enrollment, percentage of female patients, and baseline QMG or MG-ADL score. We assumed the treatment effect to be distributed according to N(μ + βx, τ^2^) and then estimated the β coefficient separately from μ to obtain a “raw” estimate of the average treatment effect. We also compared treatment efficacy with the following secondary analyses: AChR+ patients only, MuSK+ patients only, phase 3 trials, leave-one-out analysis excluding studies with outlier treatment effects, and using recently reported 52-week data for CD19+ B-cell depletion. We then compared treatment dosages using five additional trials that were prospective, but not placebo controlled.

We used Covidence for study screening and R version 4.1.1 for analysis.

## Results

A total of 27 placebo-controlled randomized trials (publication date: 1987-2025) for 17 different treatments were included in this study (patients in placebo arms: 1086; patients in treatment arms: 1232) (**Figure 1)**. We classified the treatments into nine categories based on mechanism of action: CD20+ B-cell depletion (rituximab), CD19+ B-cell depletion (inebilizumab), C5 complement inhibitors (C5i; eculizumab, ravulizumab, and zilucoplan), intravenous immunoglobulin (IVIG), FcRn inhibitors (nipocalimab, efgartigimod, rozanolixizumab, and batoclimab), CD40 inhibitor (iscalimab), IL-6 signaling inhibitor (satralizumab), B-cell activating factor inhibitor (BAFF; belimumab), and immunosuppressive therapy/steroid sparing agents (mycophenolate mofetil, tacrolimus, cyclosporine, and methotrexate). Individual study data is presented in **Table 1**.

Our model estimates that patients treated with FcRn inhibitors will experience -3.6 (95% Credible Interval: [-4.40, -2.72]) greater reduction in QMG score than those treated with placebo and standard of care; -2.59 [-3.93, -1.27] point reduction for those treated with C5 complement inhibitors, and -2.50 [-5.15, 0.13] point reduction for those treated with CD19+ B-cell depletion. Patients treated with FcRn inhibitors can expect MG-ADL scores to decrease by -1.94 [-2.23, -1.67] points more than placebo and standard of care, -1.89 [-2.95, -0.83] for CD19+ B-cell depletion, and -1.78 [-2.43, -1.13] for C5 complement inhibitors (**Figure 2** **&** 3). We calculated the relative effects of each treatment’s ability to reduce QMG and MG-ADL scores (**eFigure 4** **&** 5). In 100,000 simulations, the probability of FcRn inhibitors having the greatest improvement in QMG and MG-ADL compared to all other treatments was approximately 0.9 (**eTable 2**). We observed that a higher percentage of female patients in a study was associated with greater treatment effect magnitude (Beta = -3.16; 95% CrI = [-4.45, -1.76]) (eTable 2). We then calculated covariate-adjusted treatment effects which estimate the effect of each treatment if all studies had the same average age, female patient percentage, and baseline QMG or MG-ADL scores and estimate a greater treatment effect for CD20+ B-cell depletion under consistent trial conditions (**eTable 2, eFigure 6 & 7**).

**Figure 4.**
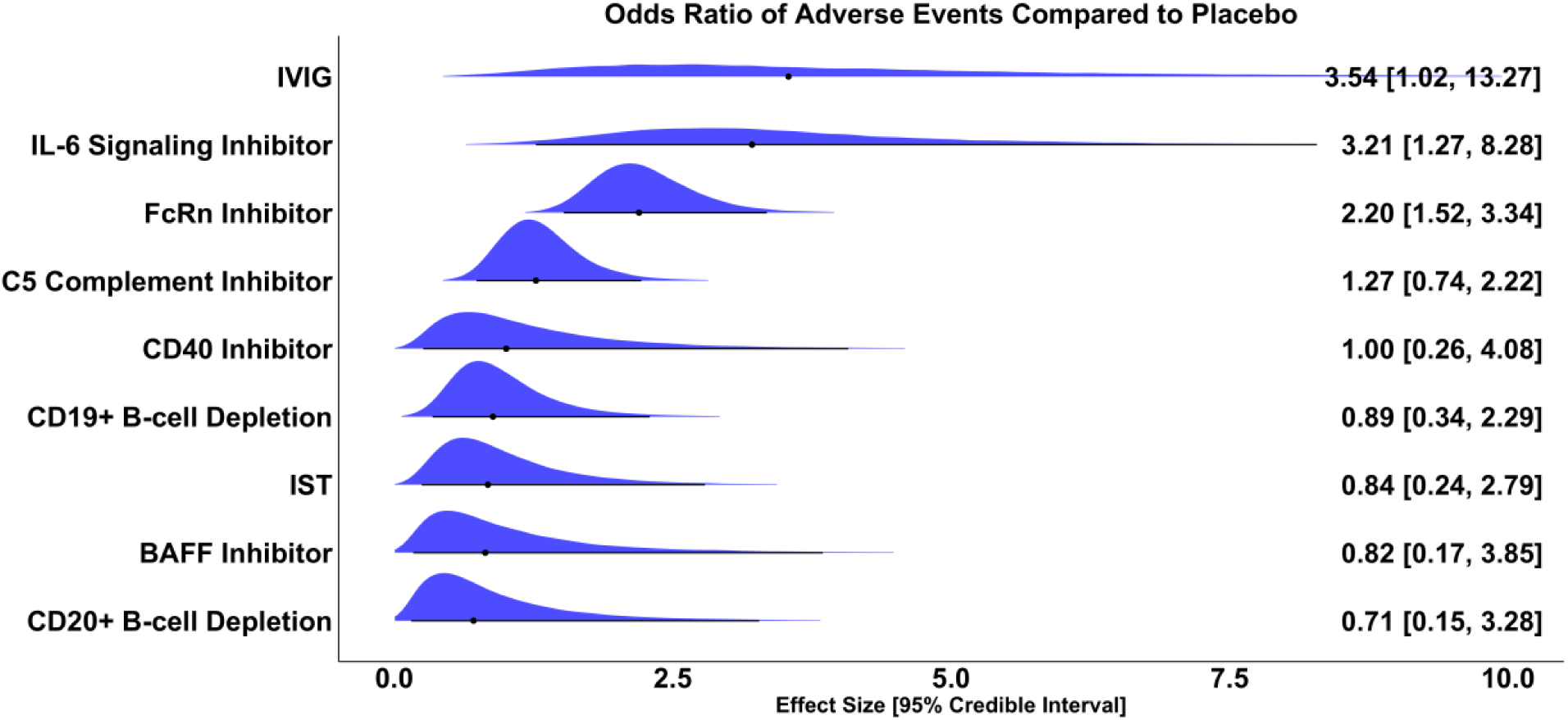
Odds ratio, 95% Credible Interval, and overlayed posterior distribution of odds of treatment-related adverse events compared to placebo. An odds ratio greater than 1 is interpreted as: patients have greater odds of treatment-related adverse events compared to placebo.

### Sensitivity Analysis

We conducted multiple sensitivity analyses to confirm these results. First, we estimated the treatment effects using 52-week data for CD19+ B-cell depletion therapy and found that the QMG treatment effect improved to -4.3 [-7.05, -1.54] and the MG-ADL treatment effect improved to -2.79 [-3.99, -1.61], the highest among all treatment types (**eFigure 8 & 9**). In simulations, the probability of CD19+ B-cell depletion having the greatest QMG and MG-ADL treatment effect became 0.868 and 0.955, respectively (**eTable 3**). Notably, one phase 2 trial of batoclimab had one of the highest treatment effects across all studies.^28^ We estimated the QMG treatment effect of FcRn inhibitors without this trial to be -2.93 [-3.72, - 2.05] and the MG-ADL effect to be -1.87 [-2.16, -1.51] (**eFigure 10 & 11**). Third, we compared data from only phase 3 trials for novel targeted therapies (FcRn inhibitors, C5 complement inhibitors, and CD19+ B-cell depletion therapy). FcRn inhibitors and CD19+ B-cell depletion demonstrated similar treatment effects for both QMG and MG-ADL reduction (**eFigure 12 & 13**).

### AChR Antibody Positive Patient Population

We then examined the efficacy of treatments in AChR+ patients. Nineteen studies (n=798 patients in treatment arms, n=787 patients in placebo arms) had QMG data and 15 studies (n=844 patients in treatment arms, n=771 patients in placebo arms) had MG-ADL data (**eFigure 14 & 15)**. FcRn inhibitors showed the greatest treatment effect with patients treated likely, on average, to see a -3.80 [-4.87, -2.72] point improvement in QMG and a -2.20 [-2.73, -1.67] point improvement in MG-ADL compared to those treated with placebo and standard of care. CD19+ B-cell depletion therapy and C5 complement inhibitors had slightly lower QMG treatment effects (CD19+ B-cell depletion: -2.51, 95% CrI = [-4.14, -0.87]; C5 complement inhibitors: -2.49, 95% CrI = [-3.35, -1.66]) and MG-ADL treatment effects (CD19+ B-cell depletion: -1.79, 95% CrI = [-2.99, -0.61]; C5 complement inhibitors: -1.77, 95% CrI = [-2.42, -1.12]) than FcRn inhibitors but showed comparable efficacy (**eFigure 16 & 17**). Estimates for the improvement in QMG and MG-ADL that AChR+ patients will experience on one treatment compared to any other are shown in league tables (**eFigure 18 & 19**). We provide calculated probabilities of each treatment having the greatest efficacy (**eTable 4**).

### MuSK Antibody Positive Patients

Five different treatment regimens from four studies reported the treatment difference in MG-ADL change from baseline for MuSK+ patients (n=59 patients in treatment arms; n=45 patients in placebo arm) (**eFigure 20**). Rozanolixizumab 7mg/kg showed the greatest efficacy in reducing MG-ADL compared to placebo and standard of care (-9.42, 95% CrI [-15.07, -3.67]) (**eFigure 21**). We estimated the relative effects of each treatment on MG-ADL reductions in MuSK+ patients and report these findings in a league table (**eFigure 22**).

### Dosage

During screening, we identified 5 additional prospective studies that were not placebo controlled, but compared two treatments.^29–33^ We leveraged the ability of the Bayesian network meta-analysis to estimate indirect treatment effects and produced a league table of all pairs of treatment dosages. Network plots of included dosages are shown in **eFigure 23 & 24** and league tables for QMG and MG-ADL are shown in **eFigure 25 & 26**, respectively.

### Safety

We calculated the odds ratio relative to placebo of treatment-related adverse events (TRAE) for studies with the available data (n=18 studies; n=1,009 patients in treatment arms; n=880 patients in placebo arms) (**eFigure 27**). CD20+ B-cell depletion (Odds Ratio: 0.70; 95% CrI: [0.15, 3.35]), the BAFF inhibitor trial (Odds Ratio: 0.81; 95% CrI: [0.16, 3.82]), and IST (Odds Ratio: 0.85; 95% CrI: [0.25, 2.85]) had the lowest odds of TRAE compared to placebo while IVIG (Odds Ratio: 3.57; 95% CrI: [1.01, 13.51]), IL-6 Signaling Inhibitors (Odds Ratio: 3.54; 95% CrI: [1.02, 13.19]), and FcRn Inhibitors (Odds Ratio: 2.20; 95% CrI: [1.52, 3.38]) had higher odds of TRAE compared to placebo (**Figure 4**). We have calculated the odds of TRAE between treatments and presented these values in a league table (**eFigure 28**).

SUCRA score rankings indicate that CD20+ B-cell depletion has the highest probability (0.735) of having the lowest odds of TRAE followed by IST (0.690). However, while IST odds ratios remained consistently below 1 after covariate adjustment and SUCRA probabilities remained around 0.7, in sensitivity analyses there were fluctuations in TRAE odds for CD20+ B-cell depletion. **(eFigure 29 & eTable 5**).

## Discussion

Existing meta-analyses have repeatedly reported superiority of FcRn inhibitors, but we show that after accounting for differences in study populations and designs, FcRn inhibitors, C5 complement inhibitors, and CD19+ B-cell depletion therapy have comparable efficacy.^6,8–11^

Interestingly, the effect of CD20+ B-cell depletion therapy improved after adjusting for age, sex, and baseline MG-ADL and QMG score. We examined the trend in treatment effect against each of these covariates and observed that trials with younger patient populations and a higher proportion of women demonstrated greater treatment effects. Conversely, treatments tended to perform similarly to placebo in trials with older populations and greater proportions of men. These relationships may be explained by the higher frequency of treatment refractory MG in younger patients and women, and further supported by recent literature that has confirmed pathomechanistic differences between early and late onset MG.^34–38^ Refractory patients would fare worse treated with standard of care in placebo arms, consequently amplifying the treatment effect. The two rituximab trials had a lower percentage of female patients, an older patient population, and lower baseline QMG and MG-ADL scores compared to the averages across studies. For reference, the RINOMAX trial was 29.8% women, had an average age of 63 years, average baseline QMG score of 9.35, and an average baseline MG-ADL score of 4.8 compared to the study wide averages in the mixed autoantibody cohort of 60.6% women, 48.8 years old, average QMG of 15.8, and average MG-ADL score of 7.8, respectively.^39^ The BeatMG study population was 44% women, had an average age of 55.1 years, average baseline QMG score of 10.1, and average baseline MG-ADL score of 4.9 compared to the study wide averages in the AChR+ trials of 54.5% women, average age of 51.9 years, average QMG score of 15.3, and average MG-ADL score of 7.8.^16^

This is further supported by the significant reduction in QMG score in the placebo arm of the RINOMAX trial compared to the non-rituximab trials (RINOMAX: -5.8; non-rituximab trials: -1.42).^39^ However, this fails to explain the effect, if any, that this vastly different patient population had on the BeatMG study because the placebo arm shows comparable reduction in QMG to other trials (BeatMG: -1.7; non-rituximab trials: -1.42). Lack of available patient-level data prevented a sensitivity analysis between early and late onset MG in our study. Nevertheless, this finding suggests that under similar experimental conditions to other trials, rituximab may show comparable efficacy to novel targeted therapies; however, without a definitive prospective study this would be speculative.

Furthermore, differences in mechanisms of action result in temporal variability of treatment response. The cyclical pattern of rapid symptomatic improvement followed by relapse that is characteristic of FcRn inhibitors starkly contrasts with the sustained benefit of CD19+ B-cell depletion, which persists to 52 weeks.^17,40^ We partially addressed this by conducting a sensitivity analysis using the 52-week randomized controlled period data of CD19+ B-cell depletion and by conducting leave-one-out sensitivity analyses of short-term FcRn inhibitor trials that were driving treatment effects to better represent their efficacy. After these sensitivity analyses, the difference between FcRn inhibitor, C5 complement inhibitors, and CD19+ B-cell depletion treatment effects attenuated.

Despite our efforts to conduct a valid comparison of MG treatments, our analysis still has several limitations. Notably absent from this meta-analysis are the azathioprine and thymectomy randomized controlled trials, two widely used immunosuppressive therapies in the treatment of MG, which were excluded because they reported median, rather than mean, change from baseline in QMG and MG-ADL.^41,42^ We used the PROMISE-MG trial, a prospective comparative study of azathioprine and mycophenolate mofetil, to indirectly estimate azathioprine’s efficacy. The results showed comparable efficacy to placebo, but this should not be misinterpreted as the treatment having no benefit. Rather, azathioprine likely shows similar efficacy to placebo because patients in placebo arms were treated with standard of care therapies such as azathioprine. Second, some studies did not report adjusted least squares mean treatment differences. In these cases, we used the unadjusted treatment difference. Third, Bayesian hierarchical models are computationally expensive thus limiting the meta-regression complexity to simple linear regression. We would like to emphasize that the covariate-adjusted treatment effects are model-based predictions rather than subgroup analyses and should not be misinterpreted as Class I evidence sourced from randomized trials. Lastly, some features of trial designs remain unaddressed such as corticosteroid tapering regimens.

Through the identification of confounding covariates and sensitivity analyses that account for differences in trial designs, we show that the reported superiority of FcRn inhibitors in previous meta-analyses may instead be a consequence of differences in trial designs and populations.^6,8–11^ Rather, FcRn inhibitors, C5 complement inhibitors, and CD19+ B-cell depletion have comparable efficacies. Our findings of associations between treatment effects and variables related to a trial’s patient population warrant future prospective studies that can further explicate the causes of subtle variations in treatment response, thus helping clinicians determine optimal treatment strategies for a given patient. Lastly, a longitudinal, prospective, comparative effectiveness study is needed to allow the collection, not only of traditional trial endpoints (i.e., MG-ADL, QMG), but also of data on disease course, burden of disease/treatment, and long-term impact of therapy on outcomes.

## Data Availability

All data produced in the present work are contained in the manuscript

## Acknowledgments

NM, MR, WZ, SH, and MF report no disclosures.

BR reports no disclosures relevant to the manuscript. He has served as a consultant for Alexion Pharmaceuticals (now part of AstraZeneca), Takeda Pharmaceuticals, Sanofi, and argenx. He has received research support from the National Institutes of Health, Takeda Pharmaceuticals, argenx, Abcuro, and Immunovant. BR also reports minor stocks in Cabaletta bio, CAVA, Pfizer.

RJN has received research support from the National Institutes of Health, Genentech, Inc., Alexion Pharmaceuticals, Inc., argenx, Annexon Biosciences, Inc., Ra Pharmaceuticals, Inc. (now UCB S.A.), the Myasthenia Gravis Foundation of America, Inc., Momenta Pharmaceuticals, Inc. (now Janssen), Immunovant, Inc., Grifols, S.A., and Viela Bio, Inc. (Horizon Therapeutics, now Amgen Inc.). He has served as a consultant/advisor for Alexion Pharmaceuticals, Inc., argenx, Cabaletta Bio, Inc., Cour Pharmaceuticals, Ra Pharmaceuticals, Inc. (now UCB S.A.), Immunovant, Inc., Momenta Pharmaceuticals, Inc. (now Janssen), and Viela Bio, Inc. (Horizon Therapeutics, now Amgen Inc.). He reports being a principal or sub-investigator on numerous trials for myasthenia gravis.

## Author Contributions

NM: Writing – Original Draft Preparation, Methodology, Data Curation, Formal Analysis, Conceptualization

MR: Data curation, Writing – Review & Editing WZ: Data curation, Writing – Review & Editing SH: Data curation, Writing – Review & Editing MF: Data curation, Writing – Review & Editing

BR: Conceptualization, Project Administration, Supervision, Writing – Review & Editing RJN: Conceptualization, Supervision, Writing – Review & Editing

