## Supplement 1 for "Comparison of the Efficacy and Safety of Myasthenia Gravis Treatments: A Bayesian Network Meta-Analysis"

Online Only Supplement

### **eTable 1.** Search strategy used to identify randomized trials.

| ID | Keywords | Results |
| --- | --- | --- |
| Ovid MEDLINE(R) ALL <1946 to May 02, 2025> |  |  |
| 1 | exp Myasthenia Gravis/ or myasthenia gravis.tw,kw. | 22288 |
| 2 | randomized controlled trial.pt. | 638558 |
| 3 | controlled clinical trial.pt. | 95692 |
| 4 | random$.ab. | 1579826 |
| 5 | (double adj2 blind).ti,ab. | 166310 |
| 6 | placebo.ti,ab. | 265639 |
| 7 | or/2-6 | 1874867 |
| 8 | 1 and 7 | 685 |
| Embase <1974 to 2025 May 02> |  |  |
| 1 | exp myasthenia gravis/ or myasthenia gravis.tw. | 30704 |
| 2 | exp randomized controlled trial/ | 881656 |
| 3 | exp controlled clinical trial/ | 1064815 |
| 4 | random*.ti,ab. | 2200974 |
| 5 | (double adj2 blind).ti,ab. | 237184 |
| 6 | placebo.ti,ab. | 392195 |
| 7 | or/2-6 | 2611151 |
| 8 | 1 and 7 | 1481 |
| Scopus |  |  |
| 1 | TITLE-ABS-KEY ( {Myasthenia gravis} ) AND TITLE-ABS-KEY ( {randomized controlled} OR {controlled clinical} OR {double blind} OR placebo OR random* ) | 1538 |
| Web of Science Core Collection (Clarivate) |  |  |
| 3 | #1 AND #2 | 1,133 |
| 2 | TS=("randomized controlled" OR "controlled clinical" OR "double blind" OR placebo OR random*) | 2,957,815 |
| 1 | TS=("Myasthenia gravis") | 21,067 |
| CENTRAL |  |  |
| 1 | "Myasthenia gravis" | 1024 |

### **eTable 2.** Unadjusted and covariate-adjusted SUCRA scores and Beta estimates for covariates across all 27 placebo-controlled studies.

| **QMG** | | | | | | | | | | |
| --- | --- | --- | --- | --- | --- | --- | --- | --- | --- | --- |
| **Unadjusted** | | **Age** | | | | **Percentage Female** | | | **Baseline Score** | |
| **Treatment** | **SUCRA** | **Treatment** | | | **SUCRA** | **Treatment** | | **SUCRA** | **Treatment** | **SUCRA** |
| Placebo | 0.100 | Placebo | | | 0.096 | Placebo | | 0.101 | Placebo | 0.086 |
| CD20+ B-cell depletion | 0.328 | CD40 Inhibitor | | | 0.168 | CD40 Inhibitor | | 0.211 | CD40 Inhibitor | 0.362 |
| CD40 Inhibitor | 0.356 | IL-6 Signaling Inhibitor | | | 0.278 | IL-6 Signaling Inhibitor | | 0.263 | IST | 0.489 |
| IL-6 Signaling Inhibitor | 0.429 | CD19+ B-cell depletion | | | 0.493 | BAFF Inhibitor | | 0.476 | IL-6 Signaling Inhibitor | 0.496 |
| IVIG | 0.506 | IVIG | | | 0.527 | CD19+ B-cell depletion | | 0.497 | IVIG | 0.522 |
| BAFF Inhibitor | 0.564 | IST | | | 0.572 | C5 Complement Inhibitor | | 0.600 | C5 Complement Inhibitor | 0.543 |
| IST | 0.602 | CD20+ B-cell depletion | | | 0.590 | IVIG | | 0.602 | CD19+ B-cell depletion | 0.562 |
| CD19+ B-cell depletion | 0.608 | C5 Complement Inhibitor | | | 0.664 | FcRn Inhibitor | | 0.680 | CD20+ B-cell depletion | 0.575 |
| C5 Complement Inhibitor | 0.635 | BAFF Inhibitor | | | 0.711 | IST | | 0.772 | FcRn Inhibitor | 0.865 |
| FcRn Inhibitor | 0.873 | FcRn Inhibitor | | | 0.903 | CD20+ B-cell depletion | | 0.797 | NA | NA |
| **MG-ADL** | | |  | | | | | | | |
| **Unadjusted** | | **Age** | | | | **Percentage Female** | | | **Baseline Score** | |
| **Treatment** | **SUCRA** | **Treatment** | | | **SUCRA** | **Treatment** | | **SUCRA** | **Treatment** | **SUCRA** |
| Placebo | 0.077 | Placebo | | | 0.076 | Placebo | | 0.074 | Placebo | 0.026 |
| BAFF Inhibitor | 0.222 | BAFF Inhibitor | | | 0.250 | BAFF Inhibitor | | 0.182 | IL-6 Signaling Inhibitor | 0.348 |
| IST | 0.413 | IST | | | 0.375 | IL-6 Signaling Inhibitor | | 0.343 | IST | 0.487 |
| IL-6 Signaling Inhibitor | 0.418 | IL-6 Signaling Inhibitor | | | 0.408 | IST | | 0.512 | CD20+ B-cell depletion | 0.554 |
| CD20+ B-cell depletion | 0.430 | CD20+ B-cell depletion | | | 0.497 | CD40 Inhibitor | | 0.528 | C5 Complement Inhibitor | 0.633 |
| CD40 Inhibitor | 0.588 | CD40 Inhibitor | | | 0.557 | CD20+ B-cell depletion | | 0.661 | CD19+ B-cell depletion | 0.669 |
| C5 Complement Inhibitor | 0.740 | C5 Complement Inhibitor | | | 0.753 | FcRn Inhibitor | | 0.694 | FcRn Inhibitor | 0.784 |
| CD19+ B-cell depletion | 0.778 | CD19+ B-cell depletion | | | 0.758 | CD19+ B-cell depletion | | 0.740 | NA |  |
| FcRn Inhibitor | 0.834 | FcRn Inhibitor | | | 0.827 | C5 Complement Inhibitor | | 0.765 | NA |  |
| **Beta Estimates** | | | |  | | | | | | |
|  |  | **QMG** | | | | **MG-ADL** | | | |  |
| **Covariate** | | **Beta** | | | **95% CrI** | **Beta** | **95% CrI** | |  | |
| Enrollment Age | | 2.12 | | | (0.83, 3.29) | 0.18 | (-0.49, 0.84) | |  | |
| Percentage Female | | -3.16 | | | (-4.45, -1.76) | -0.68 | (-1.48, 0.14) | |  | |
| Baseline Score | | -1.02 | | | (-3.26, 1.25) | -0.45 | (-1.65, 0.7) | |  | |

**eTable 3.** SUCRA scores for each treatment type, grouped by mechanism of action, using 52-week data for CD19+ B-cell depletion therapy.

| **QMG** | | **MG-ADL** | |
| --- | --- | --- | --- |
| **Treatment** | **SUCRA** | **Treatment** | **SUCRA** |
| **Placebo** | 0.097 | **Placebo** | 0.077 |
| **CD20+ B-cell depletion** | 0.307 | **BAFF Inhibitor** | 0.215 |
| **CD40 Inhibitor** | 0.336 | **IST** | 0.399 |
| **IL-6 Signaling Inhibitor** | 0.403 | **IL-6 Signaling Inhibitor** | 0.408 |
| **IVIG** | 0.479 | **CD20+ B-cell depletion** | 0.420 |
| **BAFF Inhibitor** | 0.536 | **CD40 Inhibitor** | 0.557 |
| **IST** | 0.561 | **C5 Complement Inhibitor** | 0.695 |
| **C5 Complement Inhibitor** | 0.591 | **FcRn Inhibitor** | 0.775 |
| **FcRn Inhibitor** | 0.821 | **CD19+ B-cell depletion** | 0.955 |
| **CD19+ B-cell depletion** | 0.868 | NA | NA |

### **eTable 4.** Unadjusted and covariate-adjusted SUCRA scores and Beta estimates for covariates in AChR+ patients.

| **Unadjusted** | | **Age** | | | | **Percentage Female** | | | **Baseline Score** | |
| --- | --- | --- | --- | --- | --- | --- | --- | --- | --- | --- |
| **Treatment** | **SUCRA** | **Treatment** | | | **SUCRA** | **Treatment** | | **SUCRA** | **Treatment** | **SUCRA** |
| Placebo | 0.065 | Placebo | | | 0.088 | Placebo | | 0.074 | Placebo | 0.069 |
| CD20+ B-cell+ B-cell depletion | 0.321 | CD40 Inhibitor | | | 0.258 | CD40 Inhibitor | | 0.243 | CD40 Inhibitor | 0.345 |
| CD40 Inhibitor | 0.337 | CD20+ B-cell depletion | | | 0.353 | IL-6 Signaling Inhibitor | | 0.313 | IST | 0.414 |
| IL-6 Signaling Inhibitor | 0.403 | IVIG | | | 0.498 | CD20+ B-cell depletion | | 0.464 | IVIG | 0.514 |
| IVIG | 0.520 | CD19+ B-cell depletion | | | 0.568 | IVIG | | 0.544 | CD20+ B-cell depletion | 0.549 |
| IST | 0.613 | C5 Complement Inhibitor | | | 0.607 | C5 Complement Inhibitor | | 0.604 | C5 Complement Inhibitor | 0.567 |
| CD19+ B-cell depletion | 0.643 | IST | | | 0.667 | CD19+ B-cell depletion | | 0.607 | CD19+ B-cell depletion | 0.570 |
| C5 Complement Inhibitor | 0.650 | FcRn Inhibitor | | | 0.961 | IST | | 0.726 | FcRn Inhibitor | 0.972 |
| FcRn Inhibitor | 0.949 | NA | | | NA | FcRn Inhibitor | | 0.924 | NA | NA |
| **MG-ADL** | | |  | | | | | | | |
| **Unadjusted** | | **Age** | | | | **Percentage Female** | | | **Baseline Score** | |
| **Treatment** | **SUCRA** | **Treatment** | | | **SUCRA** | **Treatment** | | **SUCRA** | **Treatment** | **SUCRA** |
| Placebo | 0.097 | Placebo | | | 0.132 | Placebo | | 0.114 | Placebo | 0.127 |
| CD20+ B-cell depletion | 0.261 | CD20+ B-cell depletion | | | 0.264 | CD20+ B-cell depletion | | 0.267 | CD20+ B-cell depletion | 0.269 |
| IL-6 Signaling Inhibitor | 0.391 | IST | | | 0.376 | IST | | 0.389 | IST | 0.444 |
| IST | 0.443 | CD40 Inhibitor | | | 0.552 | IL-6 Signaling Inhibitor | | 0.420 | CD19+ B-cell depletion | 0.605 |
| CD40 Inhibitor | 0.565 | C5 Complement Inhibitor | | | 0.619 | CD40 Inhibitor | | 0.565 | C5 Complement Inhibitor | 0.630 |
| C5 Complement Inhibitor | 0.684 | CD19+ B-cell depletion | | | 0.633 | C5 Complement Inhibitor | | 0.651 | FcRn Inhibitor | 0.925 |
| CD19+ B-cell depletion | 0.688 | FcRn Inhibitor | | | 0.925 | CD19+ B-cell depletion | | 0.655 | NA | NA |
| FcRn Inhibitor | 0.872 | NA | | | NA | FcRn Inhibitor | | 0.938 | NA | NA |
| **Beta Estimates** | | | |  | | | | | | |
|  |  | **QMG** | | | | **MG-ADL** | | | |  |
| **Covariate** | | **Beta** | | | **95% CrI** | **Beta** | **95% CrI** | |  | |
| Enrollment Age | | 0.57 | | | (-1.37, 2.49) | -0.13 | (-1.58, 1.32) | |  | |
| Percentage Female | | -1.47 | | | (-3.07, 0.16) | 0.34 | (-1.45, 2.12) | |  | |
| Baseline Score | | -1.1 | | | (-3.18, 0.99) | 0.08 | (-2.02, 2.17) | |  | |

### **eTable 5.** Unadjusted and covariate-adjusted SUCRA scores and Beta estimates for odds of treatment-related adverse events.

| **Unadjusted** | | **Study Length** | | **Age** | | **Female** | |
| --- | --- | --- | --- | --- | --- | --- | --- |
| **Treatment** | **SUCRA** | **Treatment** | **SUCRA** | **Treatment** | **SUCRA** | **Treatment** | **SUCRA** |
| IL-6 Signaling Inhibitor | 0.119 | IVIG | 0.235 | IVIG | 0.135 | IL-6 Signaling Inhibitor | 0.041 |
| IVIG | 0.119 | CD40 Inhibitor | 0.249 | IL-6 Signaling Inhibitor | 0.183 | FcRn Inhibitor | 0.216 |
| FcRn Inhibitor | 0.225 | IL-6 Signaling Inhibitor | 0.257 | FcRn Inhibitor | 0.317 | IVIG | 0.237 |
| C5 Complement Inhibitor | 0.495 | CD19+ B-cell depletion | 0.274 | C5 Complement Inhibitor | 0.386 | CD19+ B-cell depletion | 0.365 |
| CD40 Inhibitor | 0.608 | CD20+ B-cell depletion | 0.377 | BAFF Inhibitor | 0.461 | C5 Complement Inhibitor | 0.412 |
| Placebo | 0.645 | FcRn Inhibitor | 0.592 | CD20+ B-cell depletion | 0.569 | Placebo | 0.652 |
| BAFF Inhibitor | 0.68 | BAFF Inhibitor | 0.603 | Placebo | 0.661 | CD40 Inhibitor | 0.711 |
| CD19+ B-cell depletion | 0.683 | C5 Complement Inhibitor | 0.741 | CD19+ B-cell depletion | 0.733 | IST | 0.762 |
| IST | 0.69 | IST | 0.808 | CD40 Inhibitor | 0.748 | BAFF Inhibitor | 0.78 |
| CD20+ B-cell depletion | 0.735 | Placebo | 0.864 | IST | 0.807 | CD20+ B-cell depletion | 0.826 |
| **Beta Estimates** | | | | | | |  |
| **Covariate** | | **Beta** | | **95% CrI** | |  | |
| Study Length | | -0.58 | | (-1.5, 0.41) | |  | |
| Enrollment Age | | -1.45 | | (-2.66, -0.26) | |  | |
| Percentage Female | | -0.92 | | (-1.78, -0.1) | |  | |

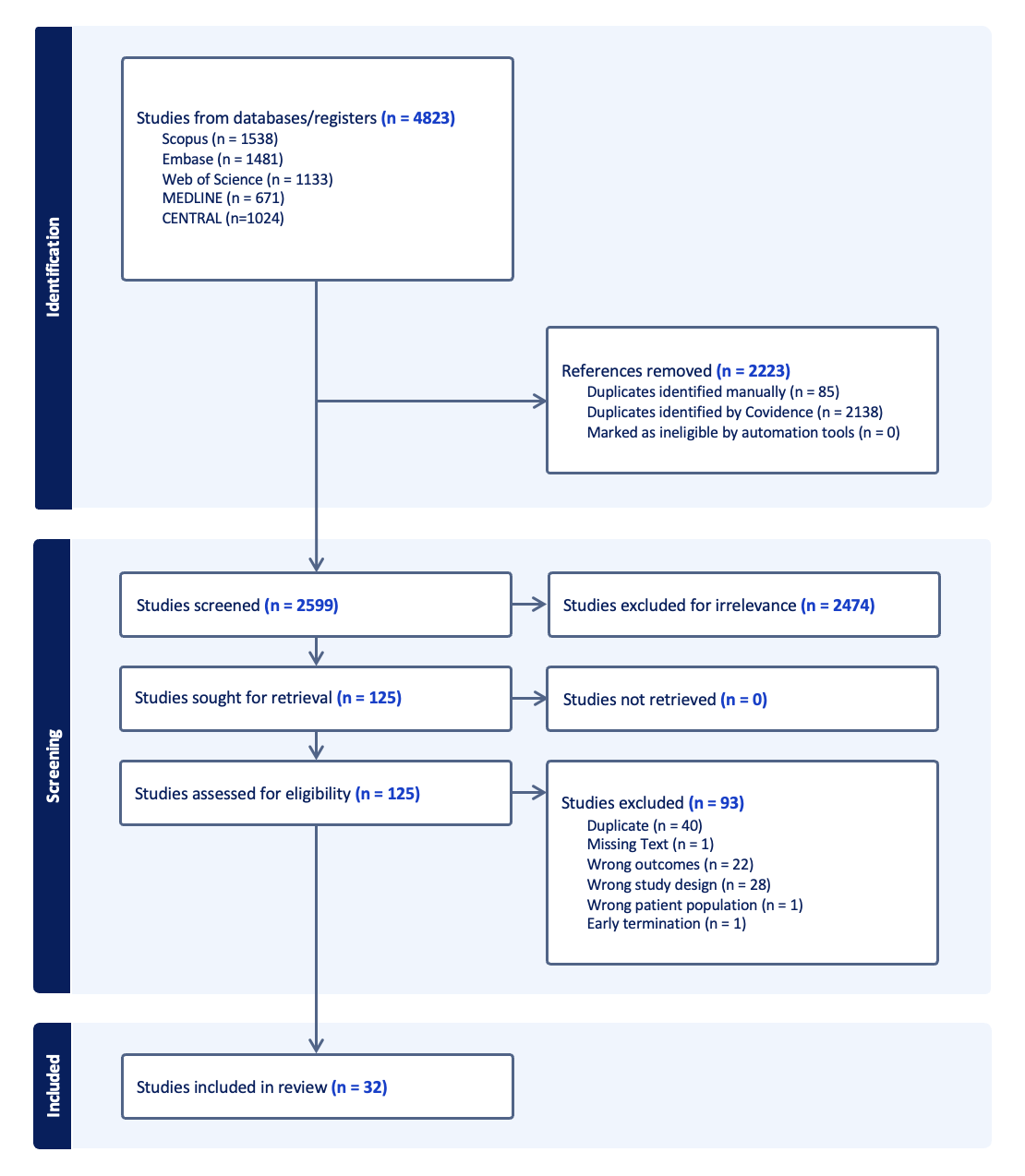

### **eFigure 1.** Preferred Reporting Items for Systematic reviews and Meta-Analyses (PRISMA) flowchart of the screening process.

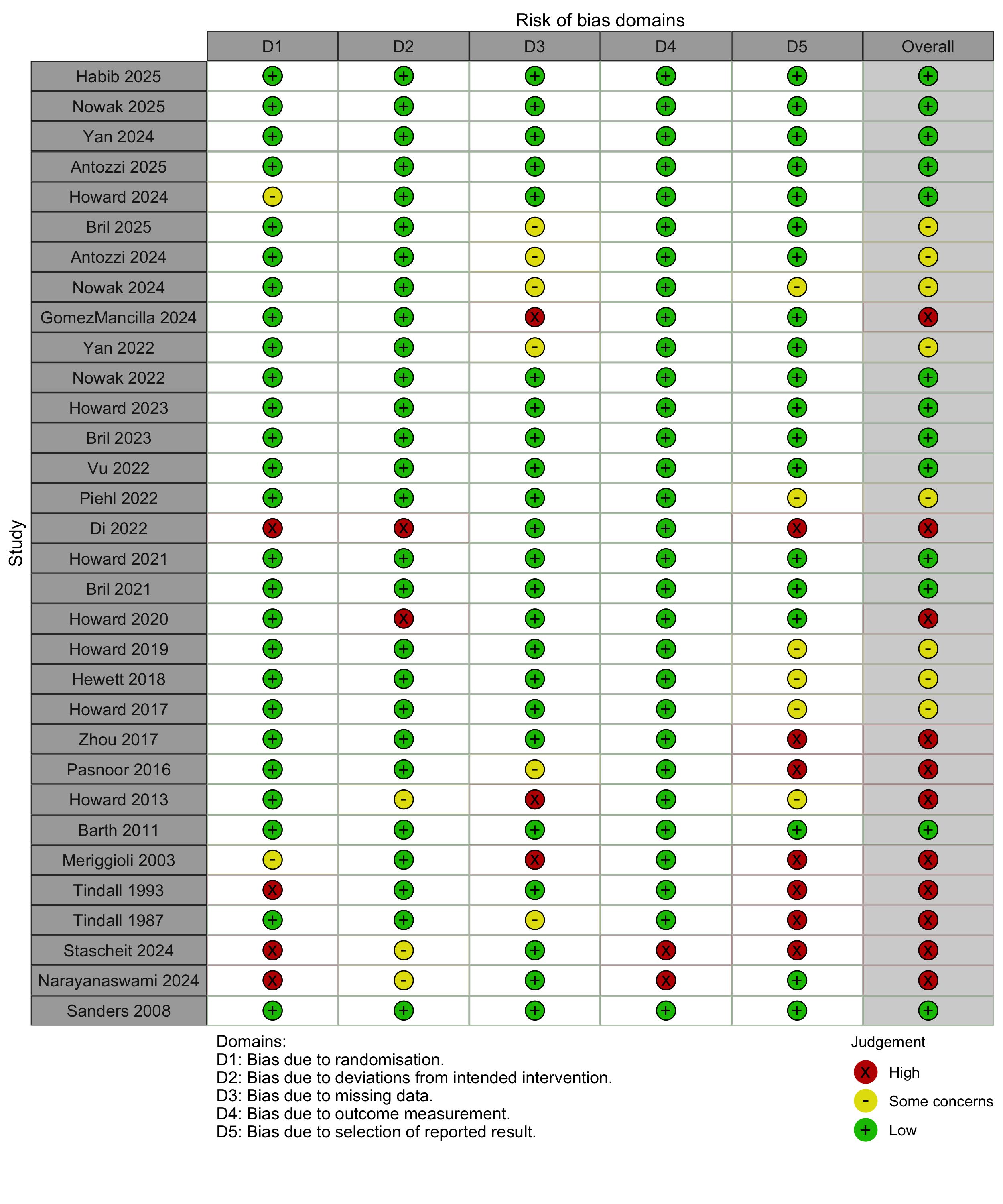
**eFigure 2.** Individual risk of bias assessments for included studies.

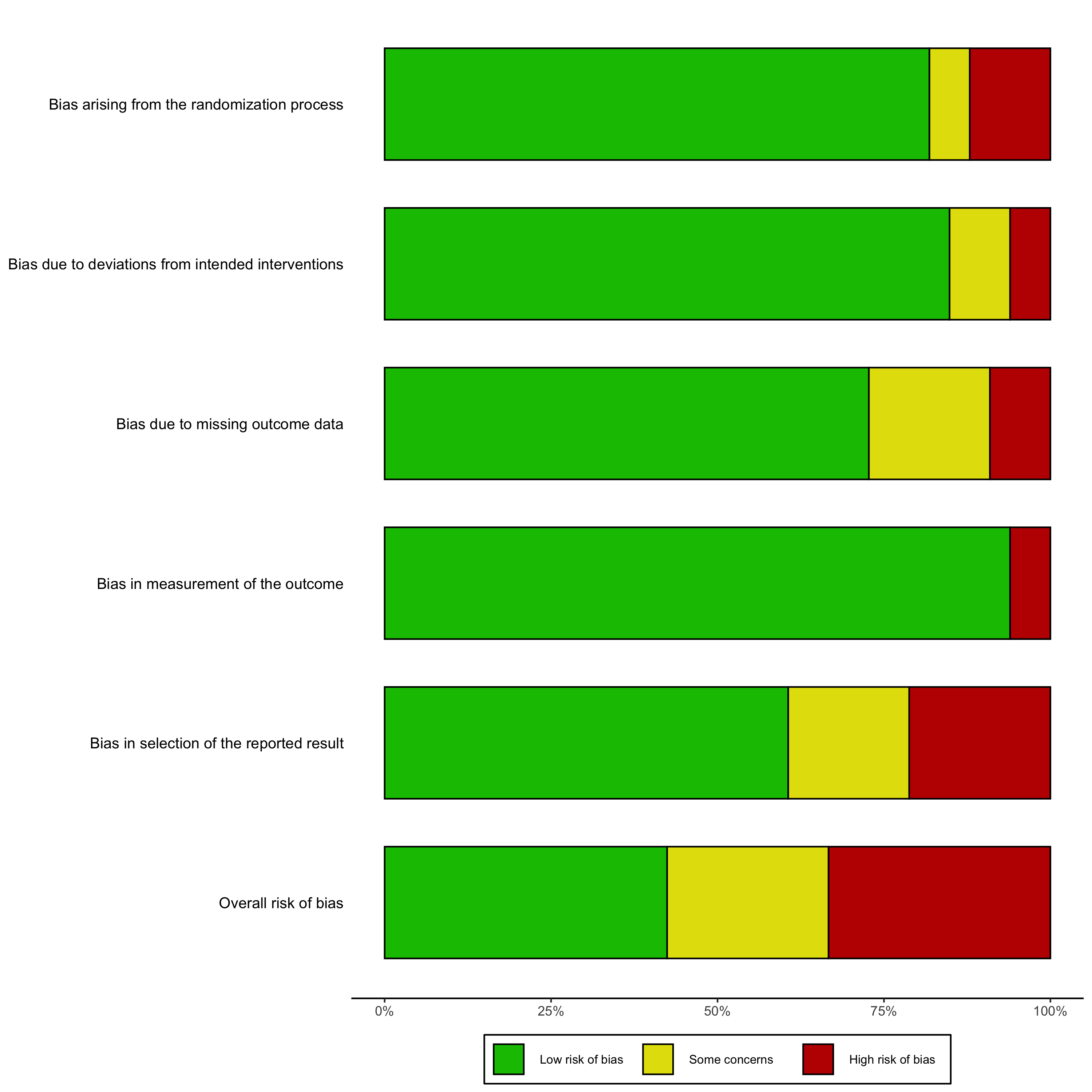

### **eFigure 3.** Summary plot of risk of bias across all studies.

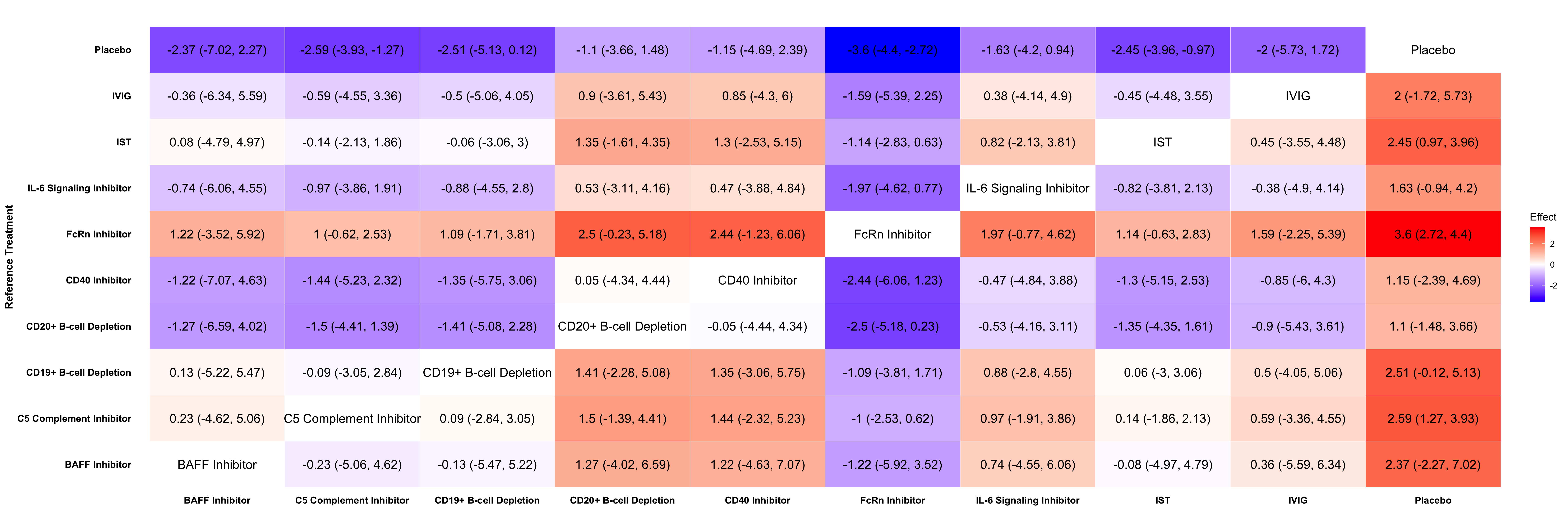

### **eFigure 4.** League table of relative treatment effect of QMG change from baseline across all pairs of treatments using data from all 27 placebo-controlled trials. Reference treatments are on the rows. A treatment effect of -1 in row A and column B is interpreted as: treatment B reduces the outcome measure by one more point than treatment A.

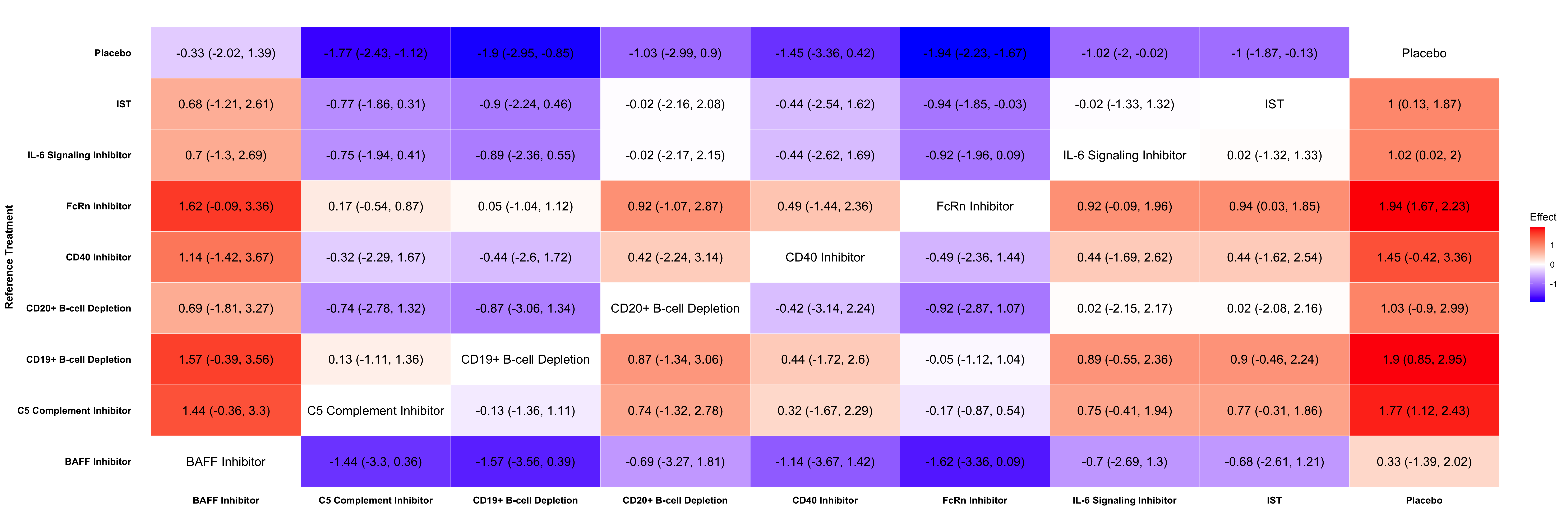

### **eFigure 5.** League table of relative treatment effect of MG-ADL change from baseline across all pairs of treatments using data from all 27 placebo-controlled trials. Reference treatments are on the rows. A treatment effect of -1 in row A and column B is interpreted as: treatment B reduces the outcome measure by one more point than treatment A.

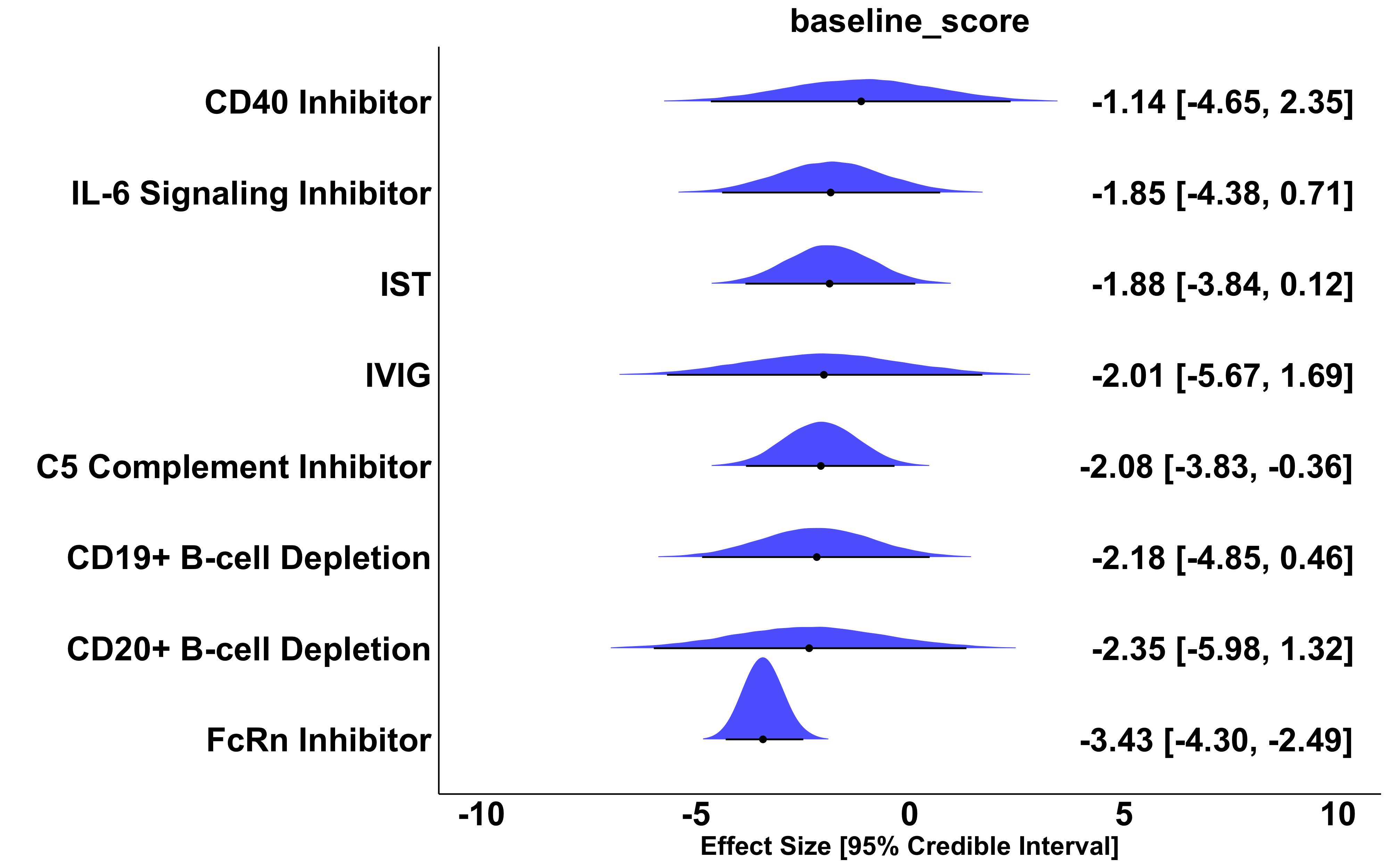

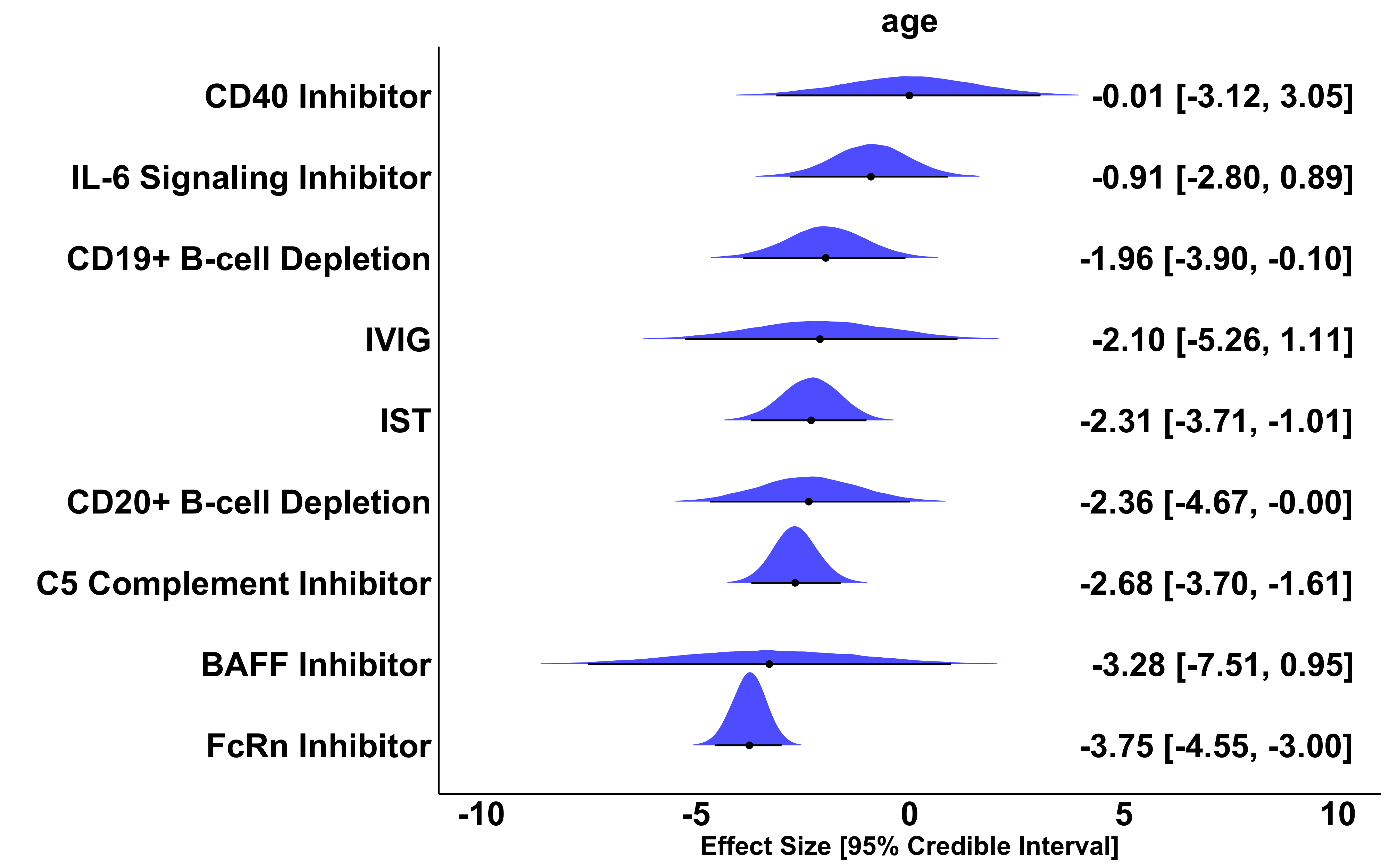

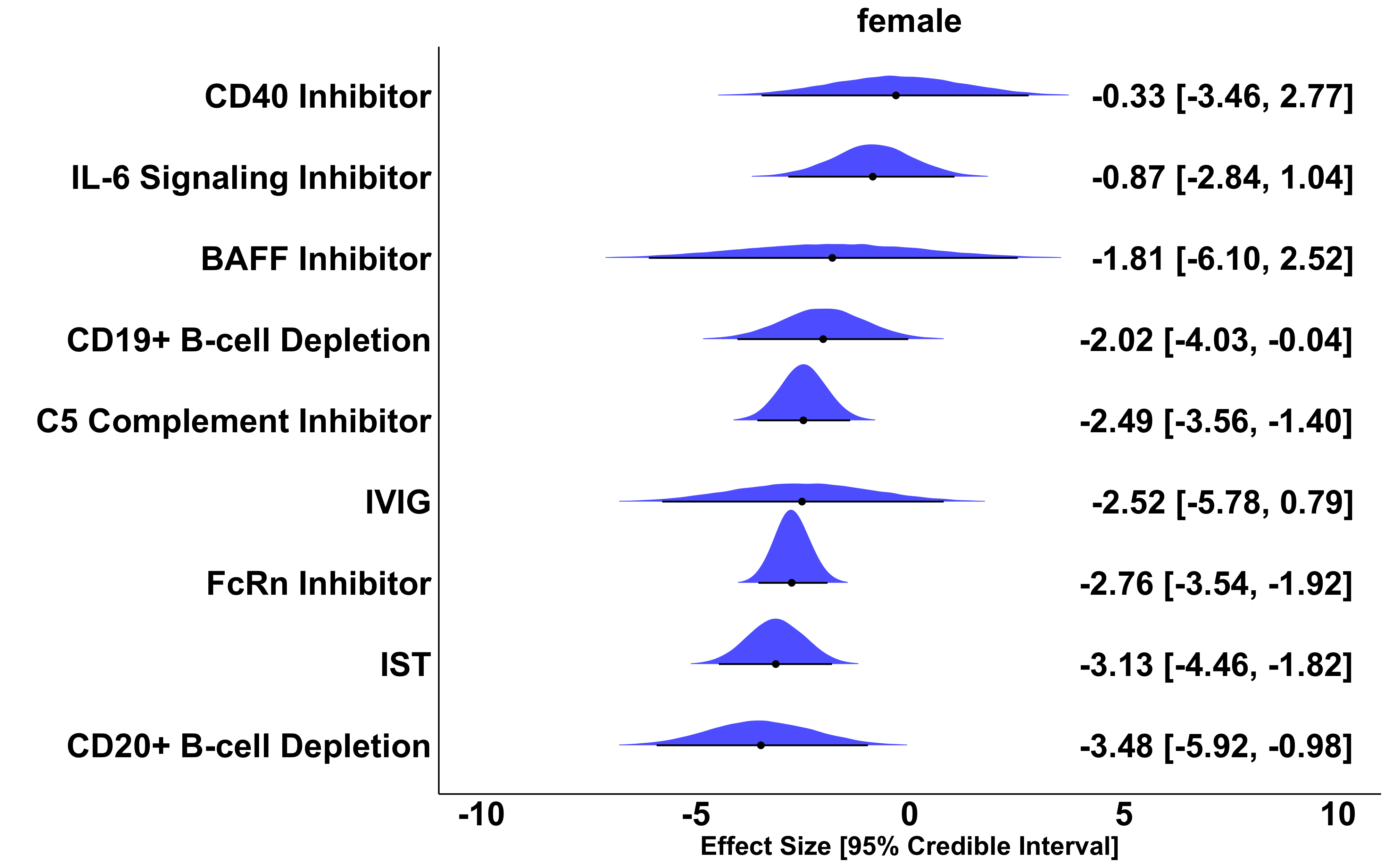

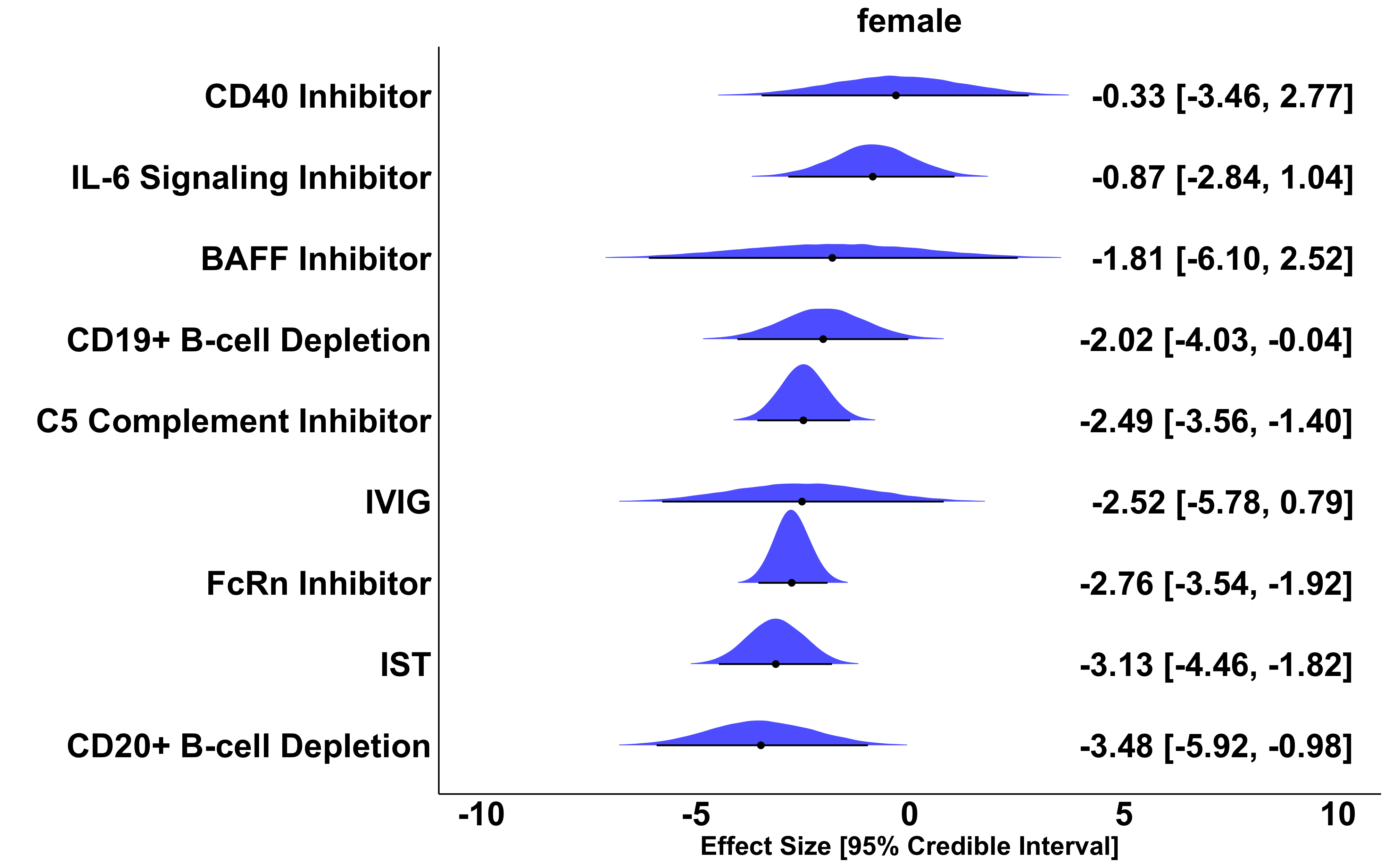

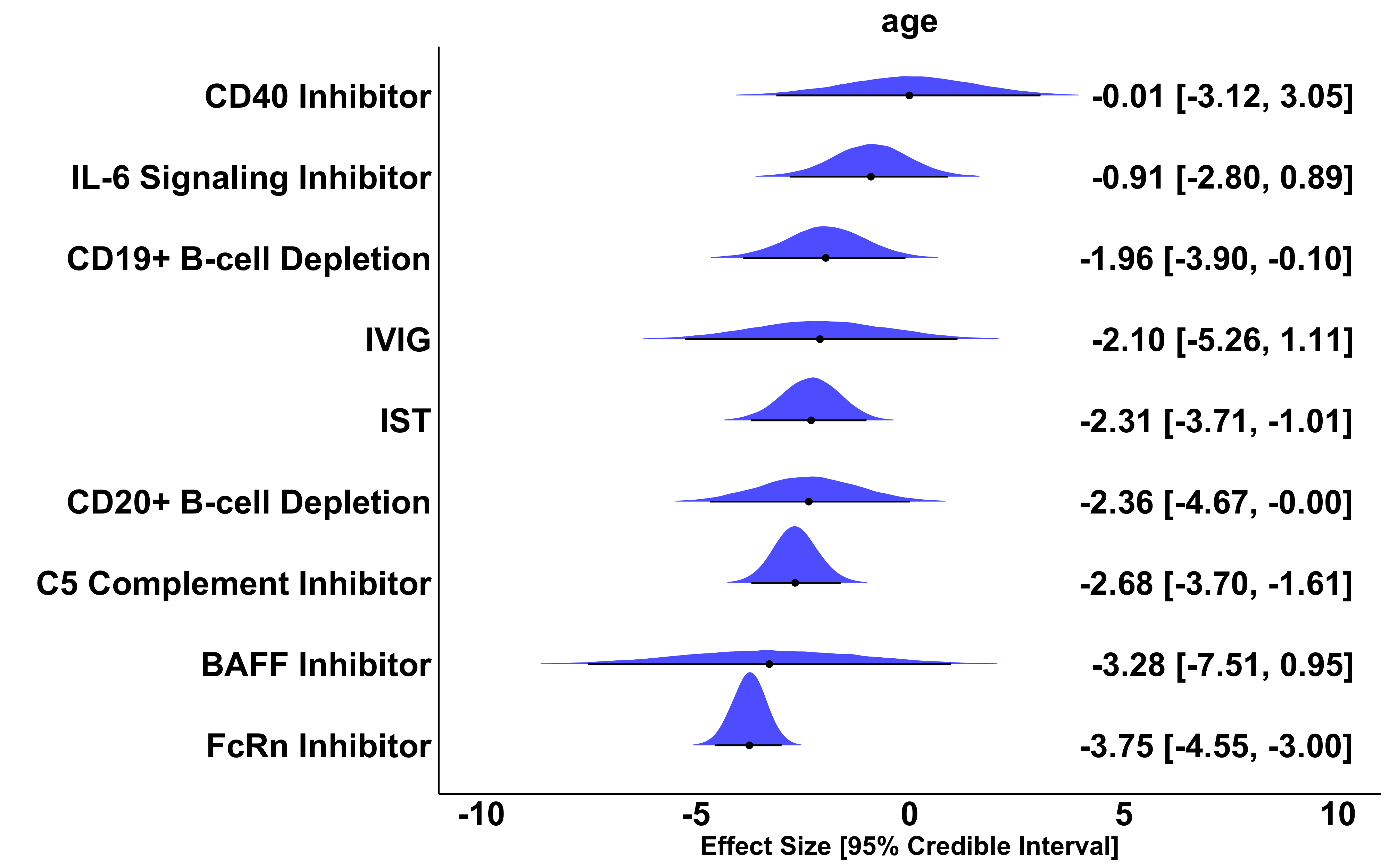

### **eFigure 6.** Effect size, 95% Credible Interval, and overlayed posterior distribution of QMG change from baseline compared to placebo across all placebo-controlled studies with available data adjusted for study-wide baseline QMG score, age at enrollment, and percentage of female participants. All adjusted effect sizes represent the treatment effect if all studies had the same value of the covariate (here we use the average across all studies; average baseline QMG: 15.2, average enrollment age: 52.2 years, average percentage of female participants: 54.0%). A negative effect size of x is interpreted as: the treatment reduces QMG x points more than placebo and standard of care.

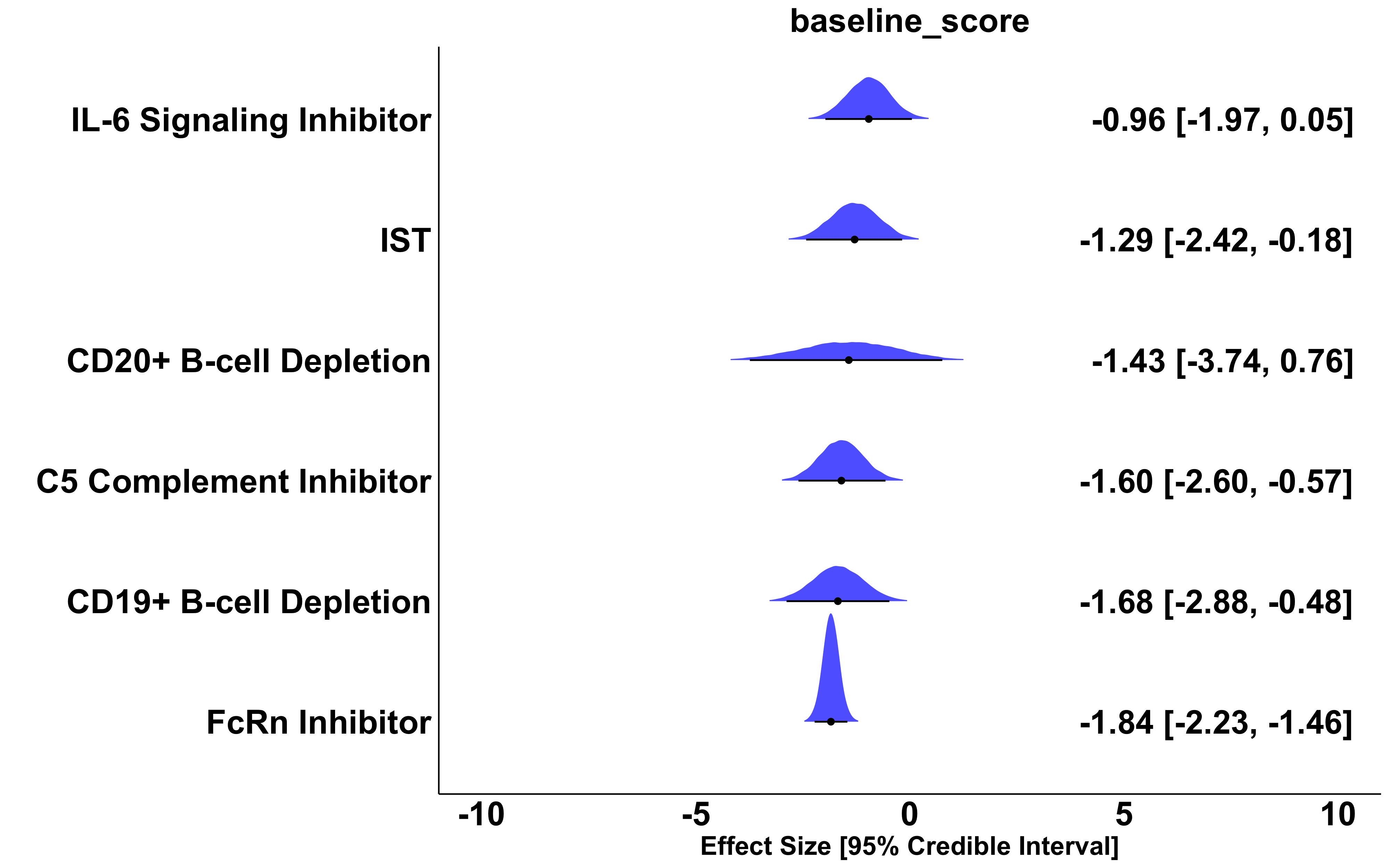

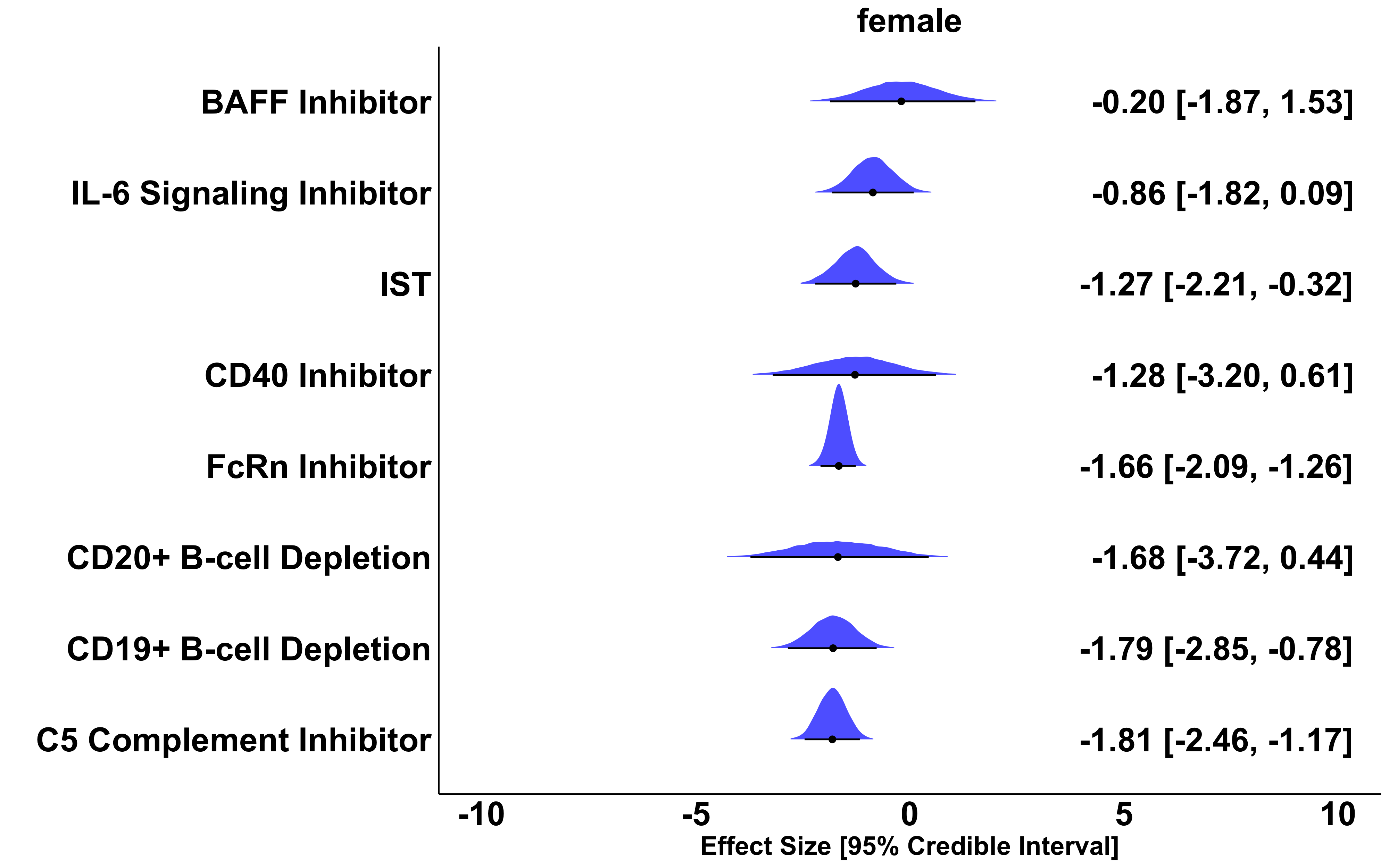

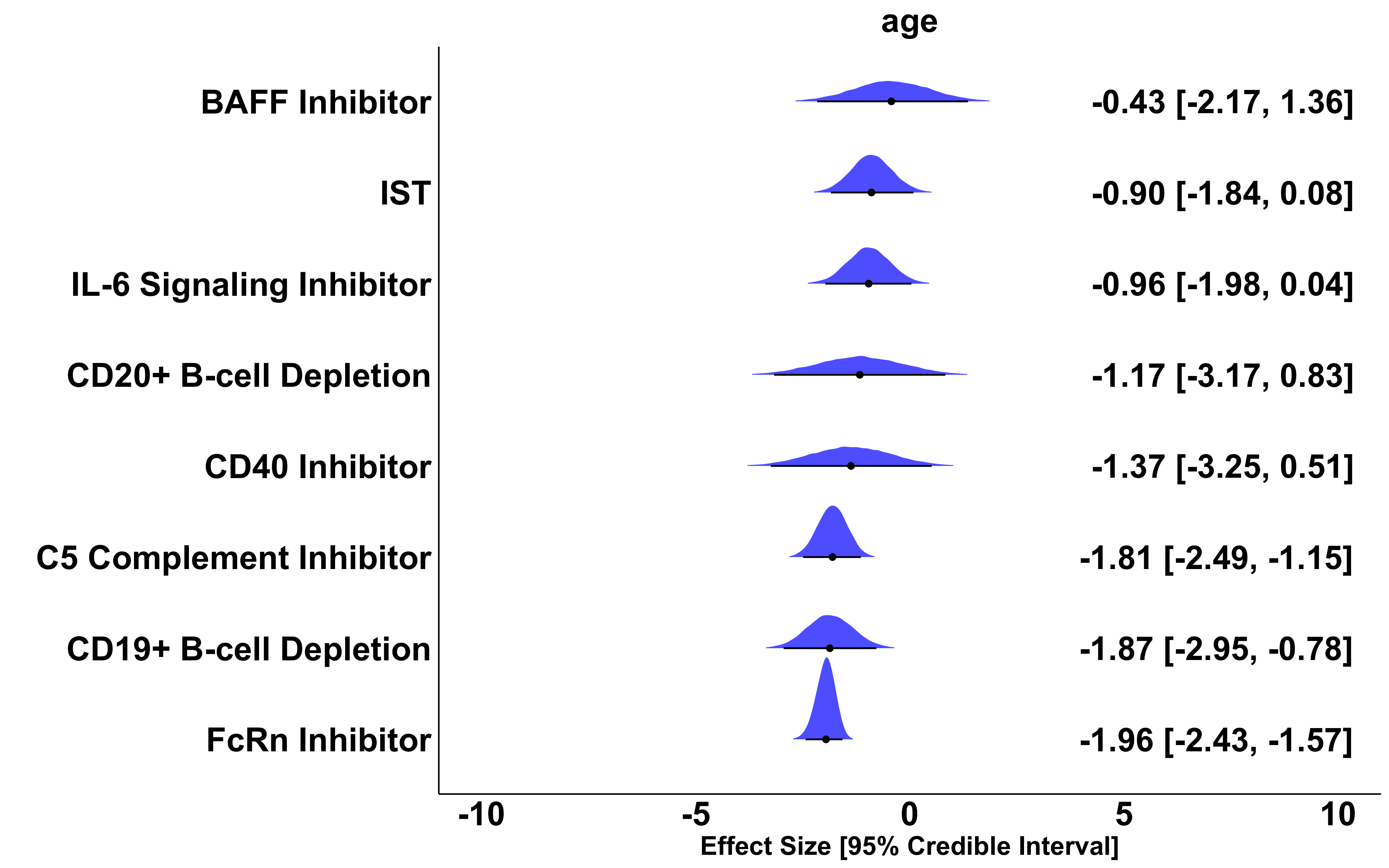

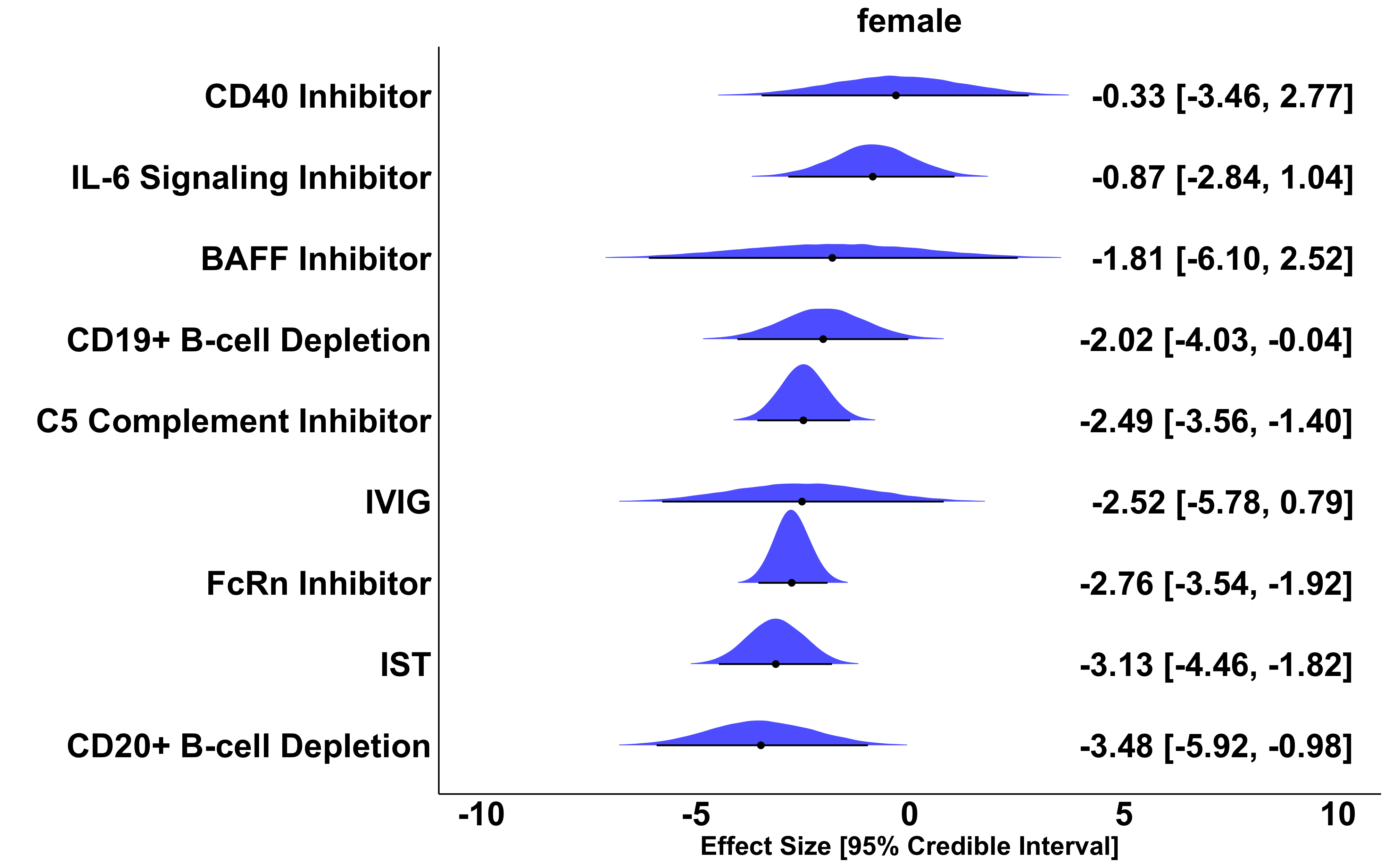

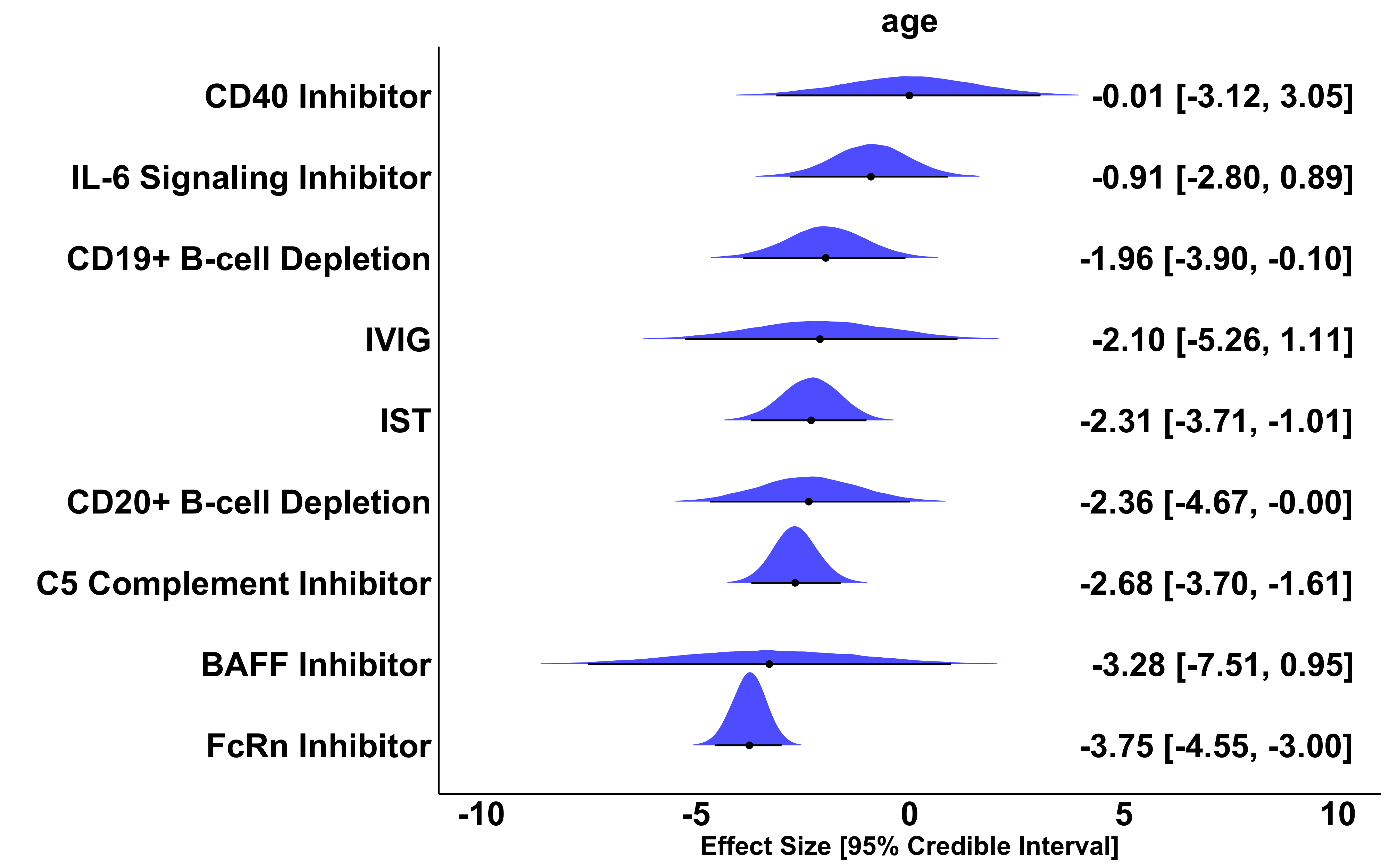

### **eFigure 7.** Effect size, 95% Credible Interval, and overlayed posterior distribution of MG-ADL change from baseline compared to placebo across all placebo-controlled studies with available data adjusted for study-wide baseline MG-ADL score, age at enrollment, and percentage of female participants. All adjusted effect sizes represent the treatment effect if all studies had the same value of the covariate (here we use the average across all studies; average baseline MG-ADL: 7.7, average enrollment age: 51.1 years, average percentage of female participants: 53.5%). A negative effect size of x is interpreted as: the treatment reduces MG-ADL x points more than placebo and standard of care.

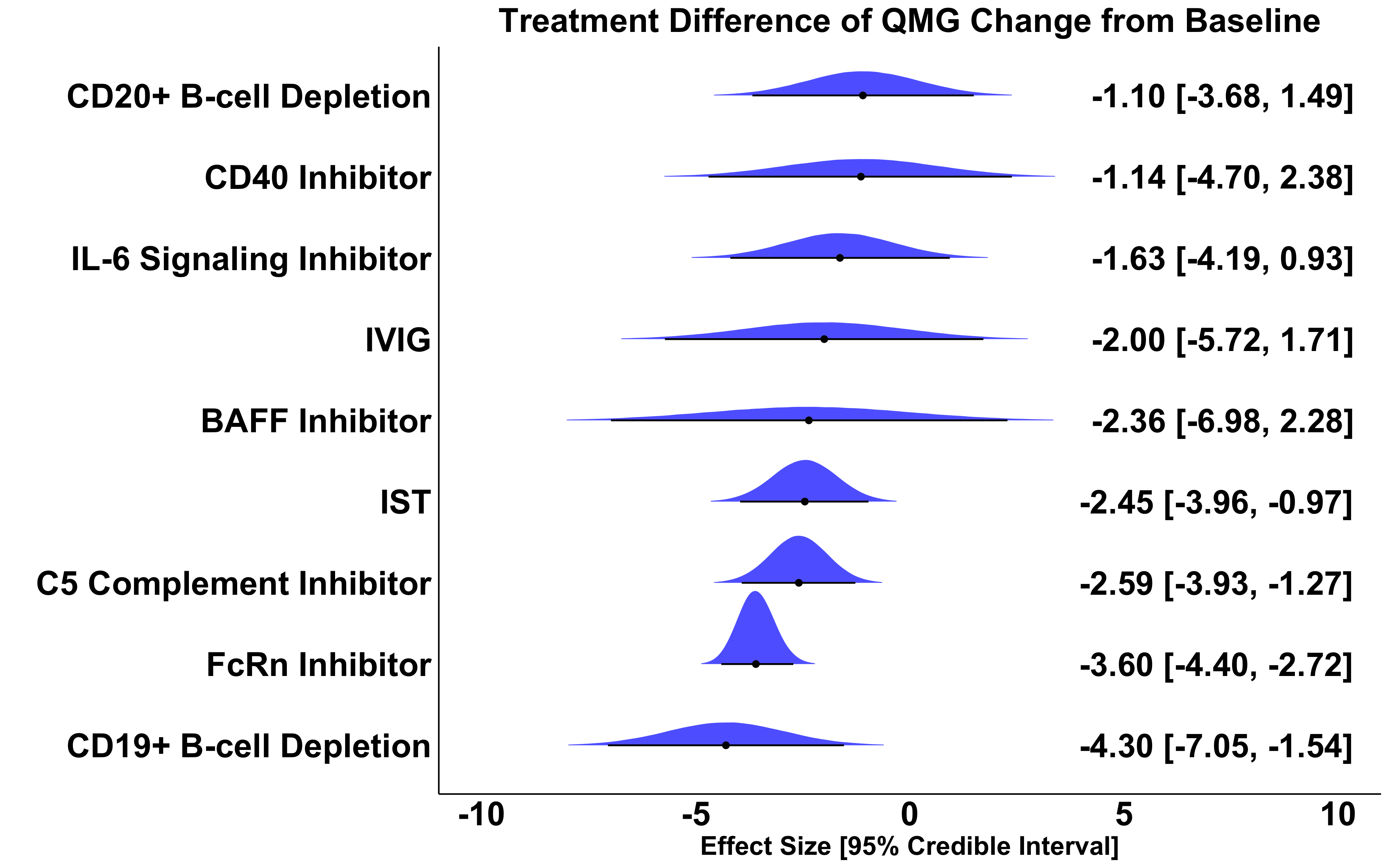

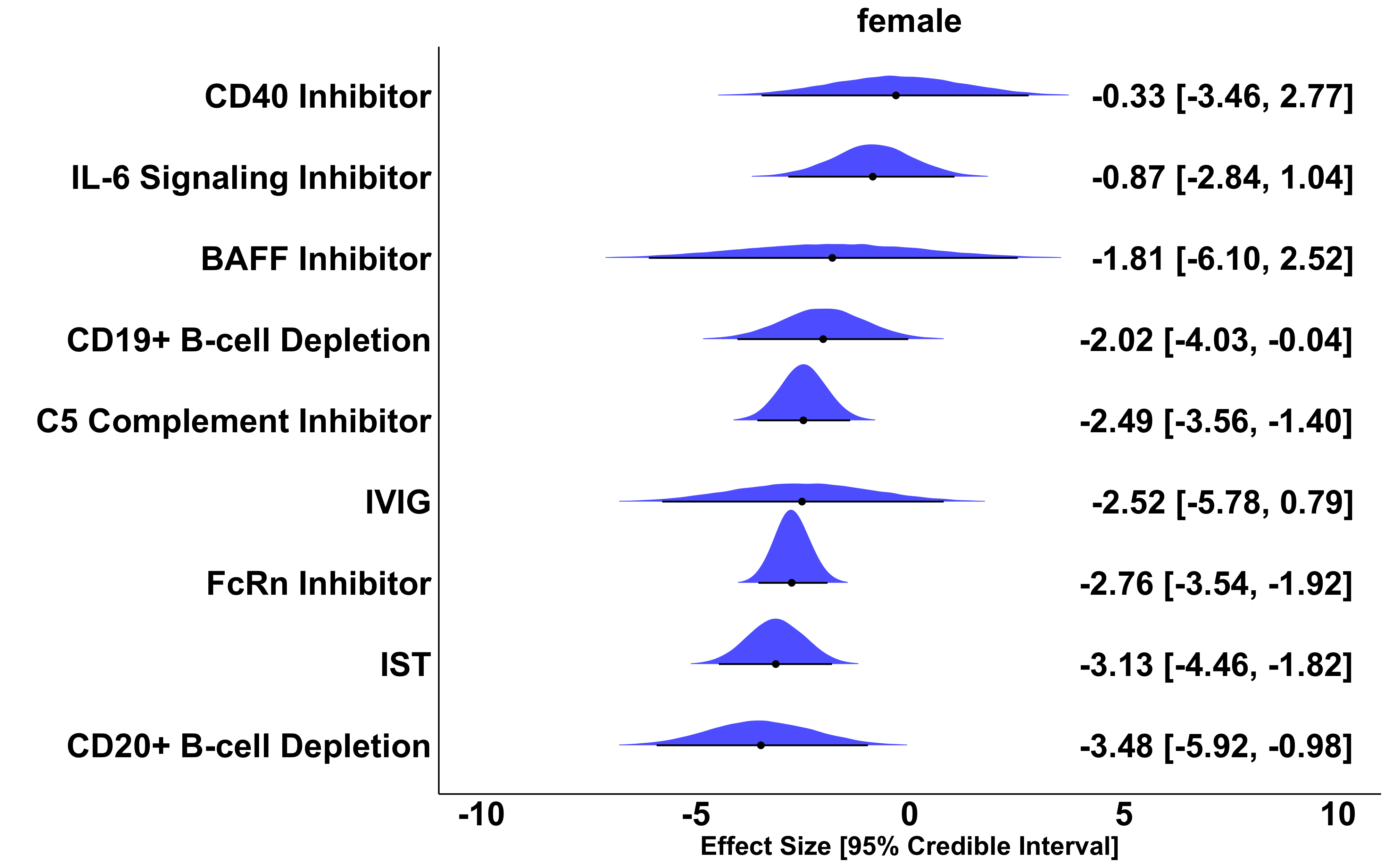

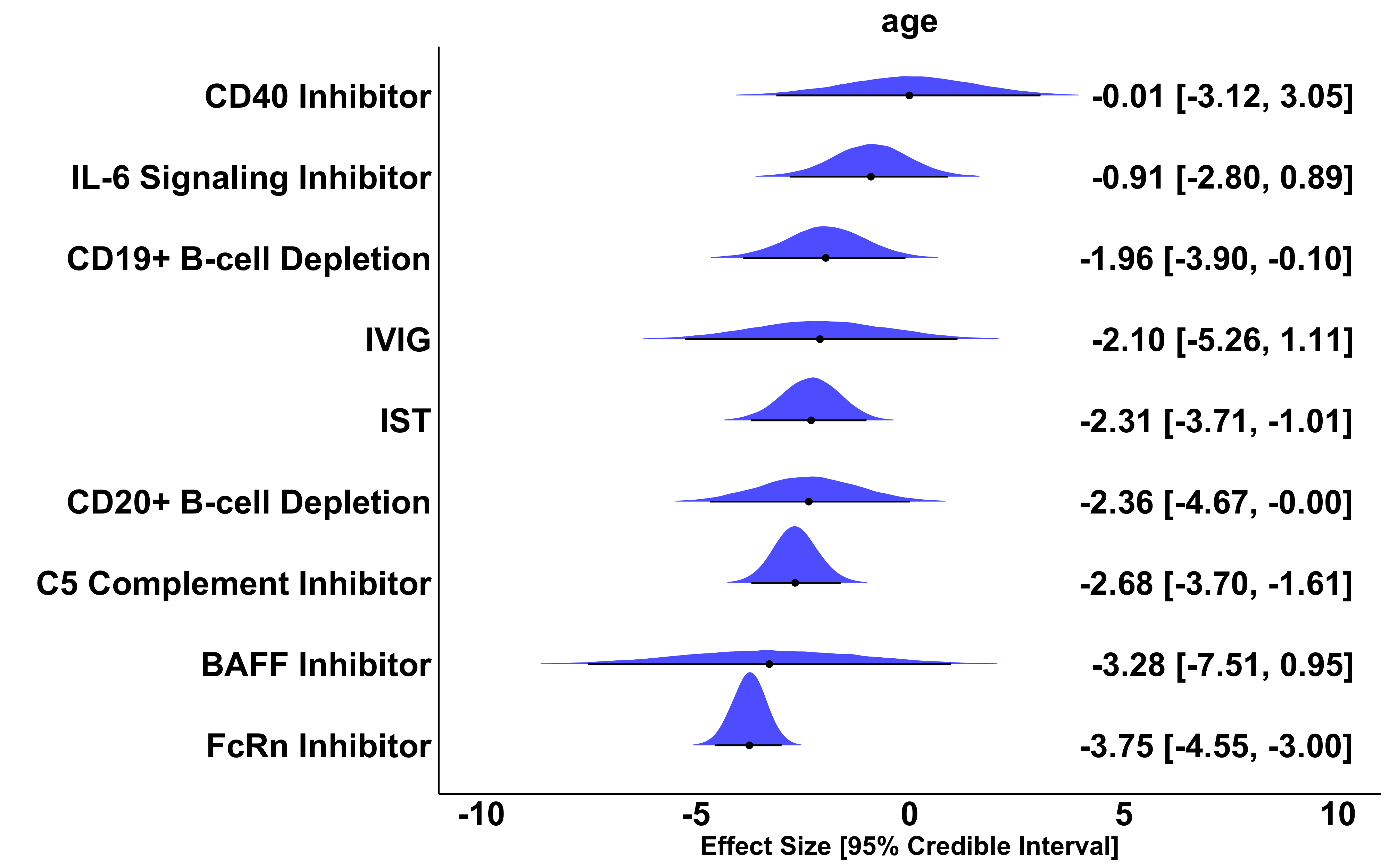
**eFigure 8.** Effect size, 95% Credible Interval, and overlayed posterior distribution of QMG change from baseline compared to placebo across all 27 placebo-controlled studies using 52-week CD19+ B-cell depletion therapy data. A negative effect size of x is interpreted as: the treatment reduces QMG x points more than placebo and standard of care.

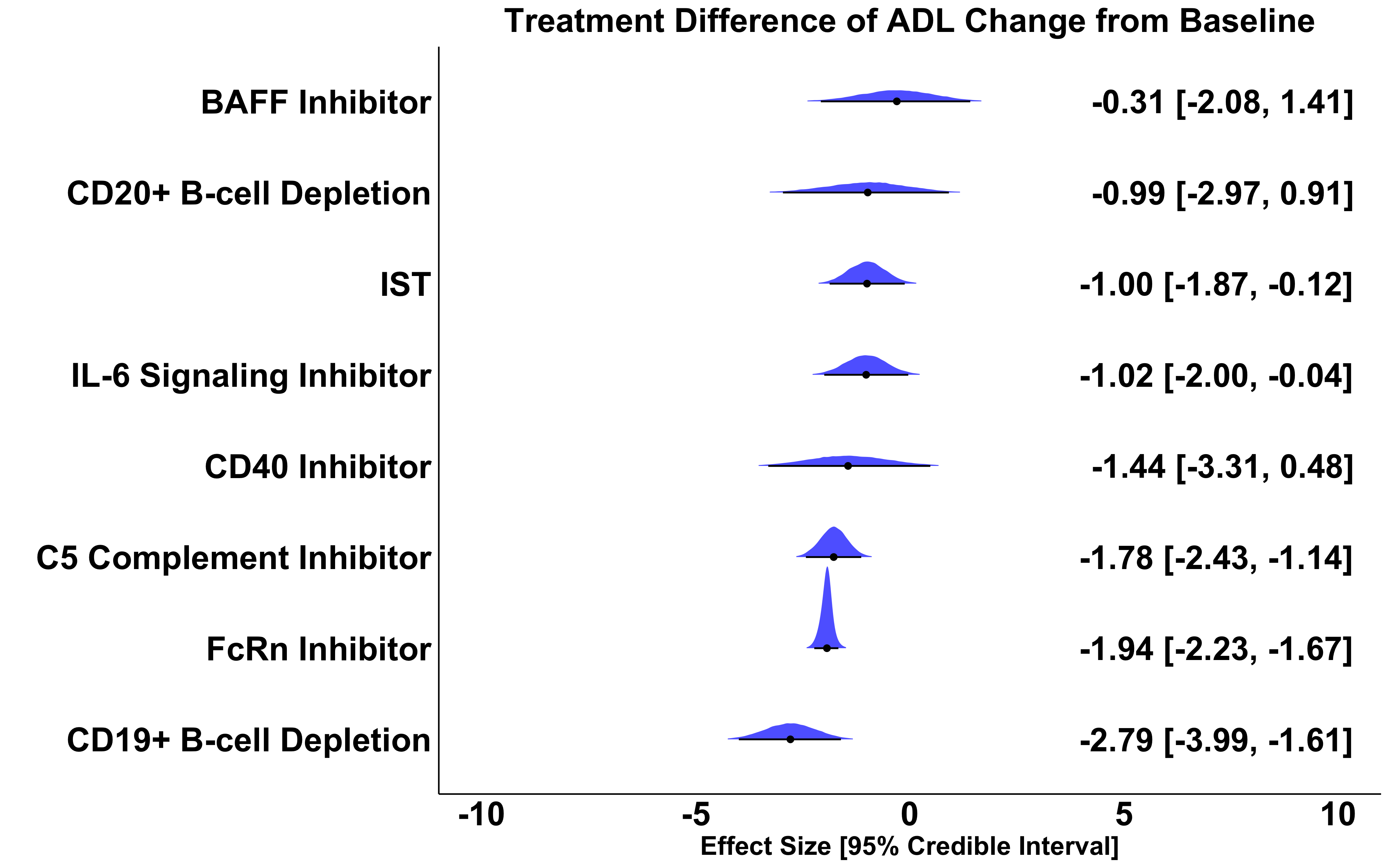

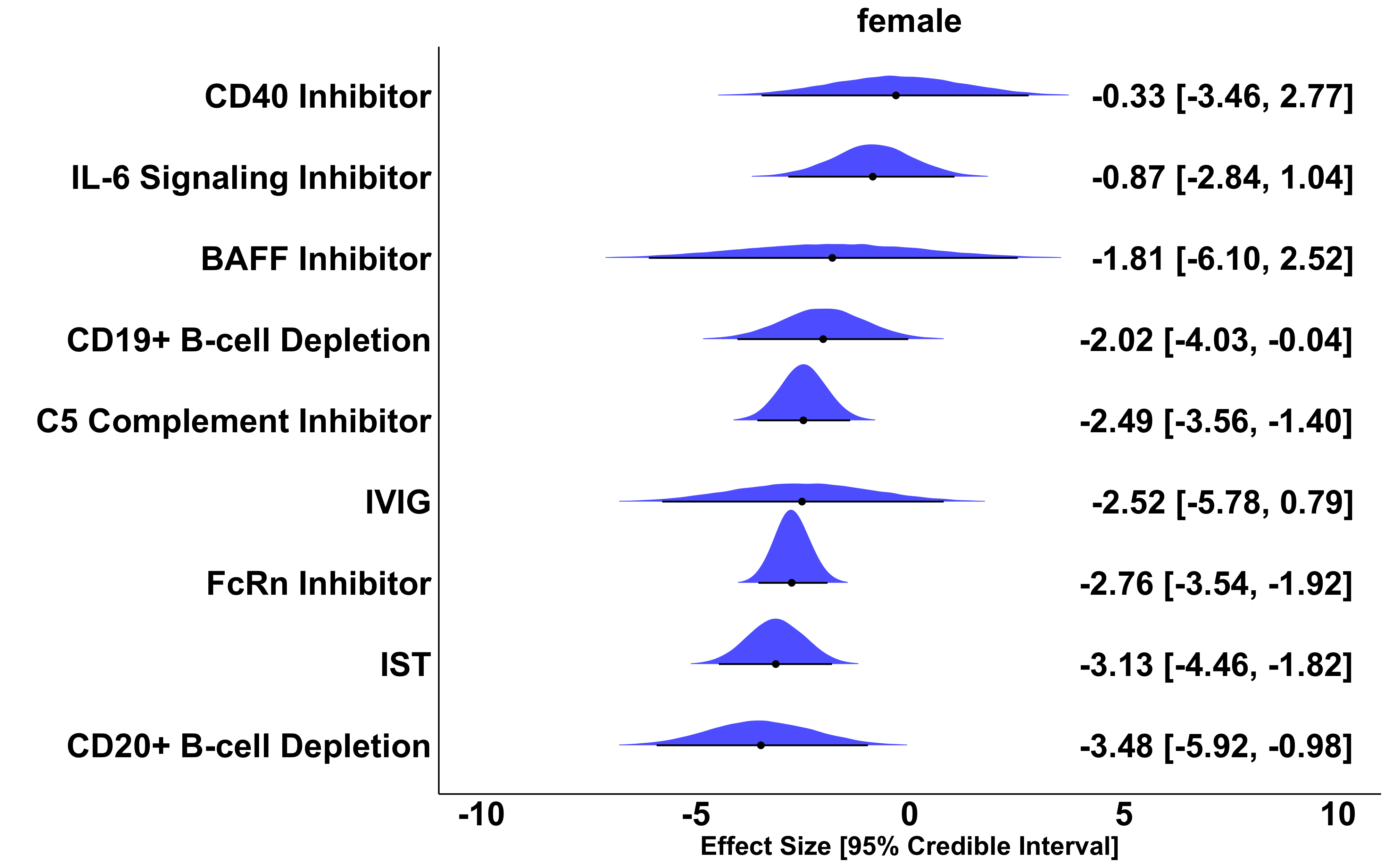

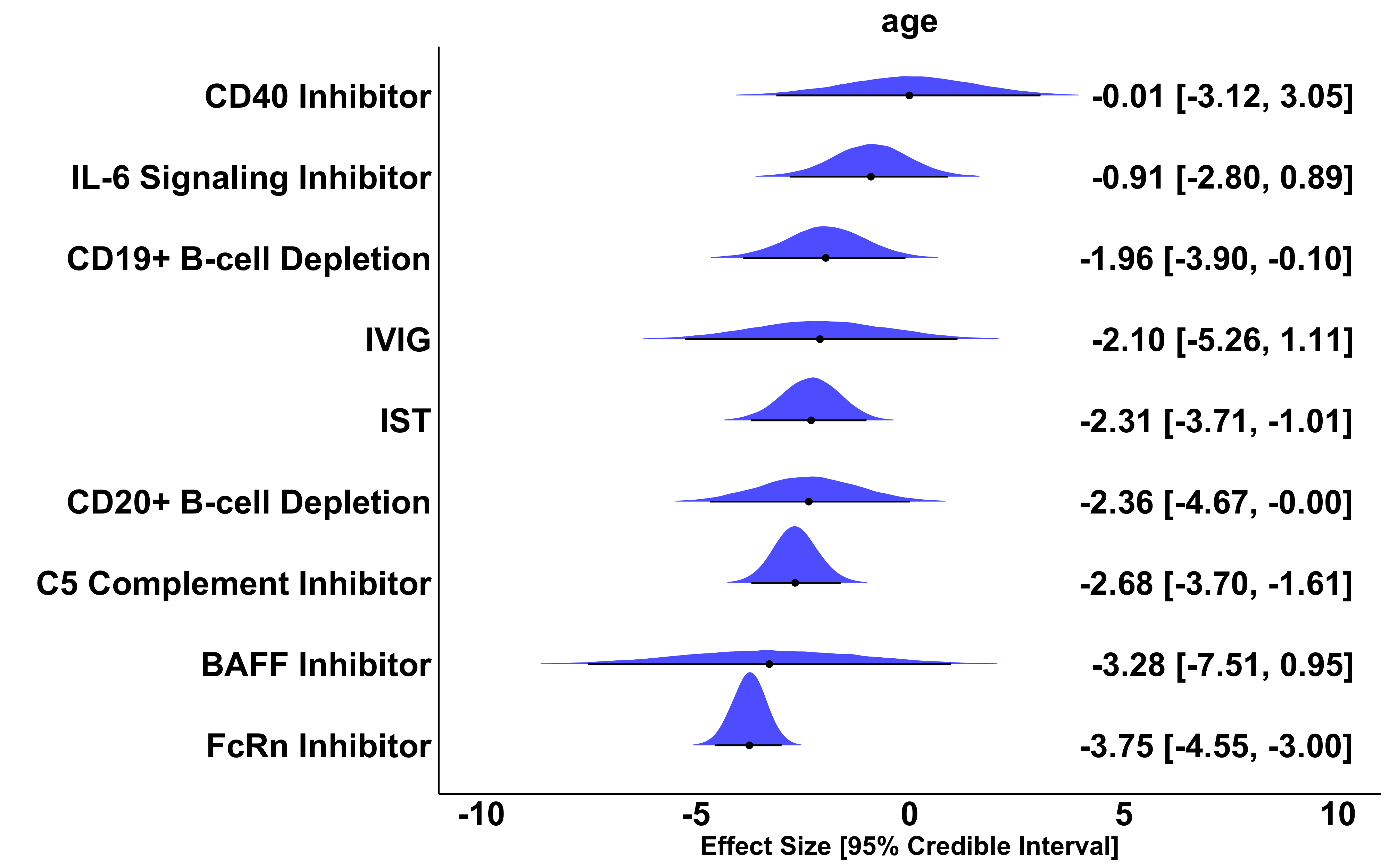
**eFigure 8.** Effect size, 95% Credible Interval, and overlayed posterior distribution of MG-ADL change from baseline compared to placebo across all 27 placebo-controlled studies using 52-week CD19+ B-cell depletion therapy data. A negative effect size of x is interpreted as: the treatment reduces MG-ADL x points more than placebo and standard of care.

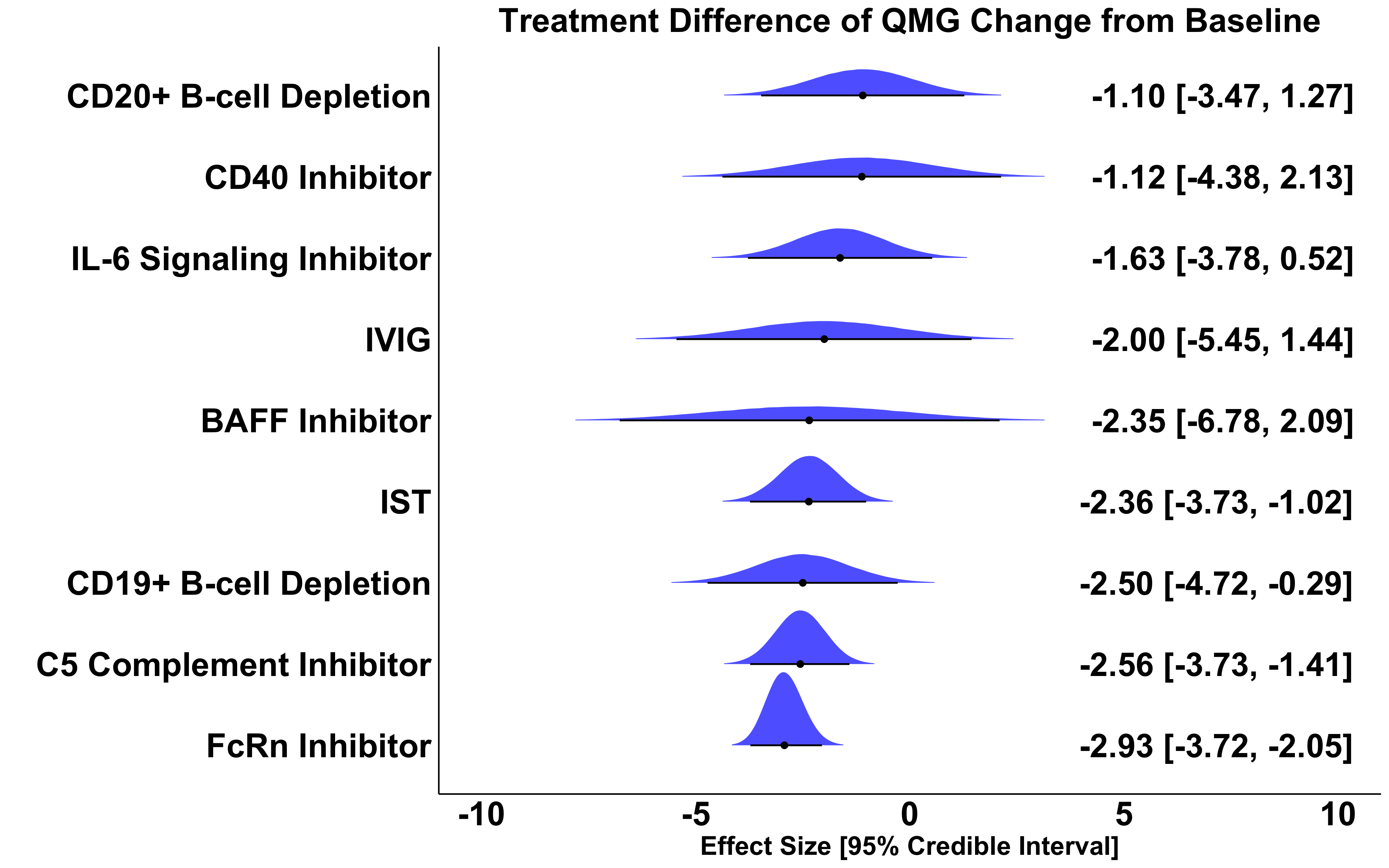

### **eFigure 10.** Effect size, 95% Credible Interval, and overlayed posterior distribution of QMG change from baseline compared to placebo across all 27 placebo-controlled studies excluding phase 2 batoclimab trial. A negative effect size of x is interpreted as: the treatment reduces QMG x points more than placebo and standard of care.

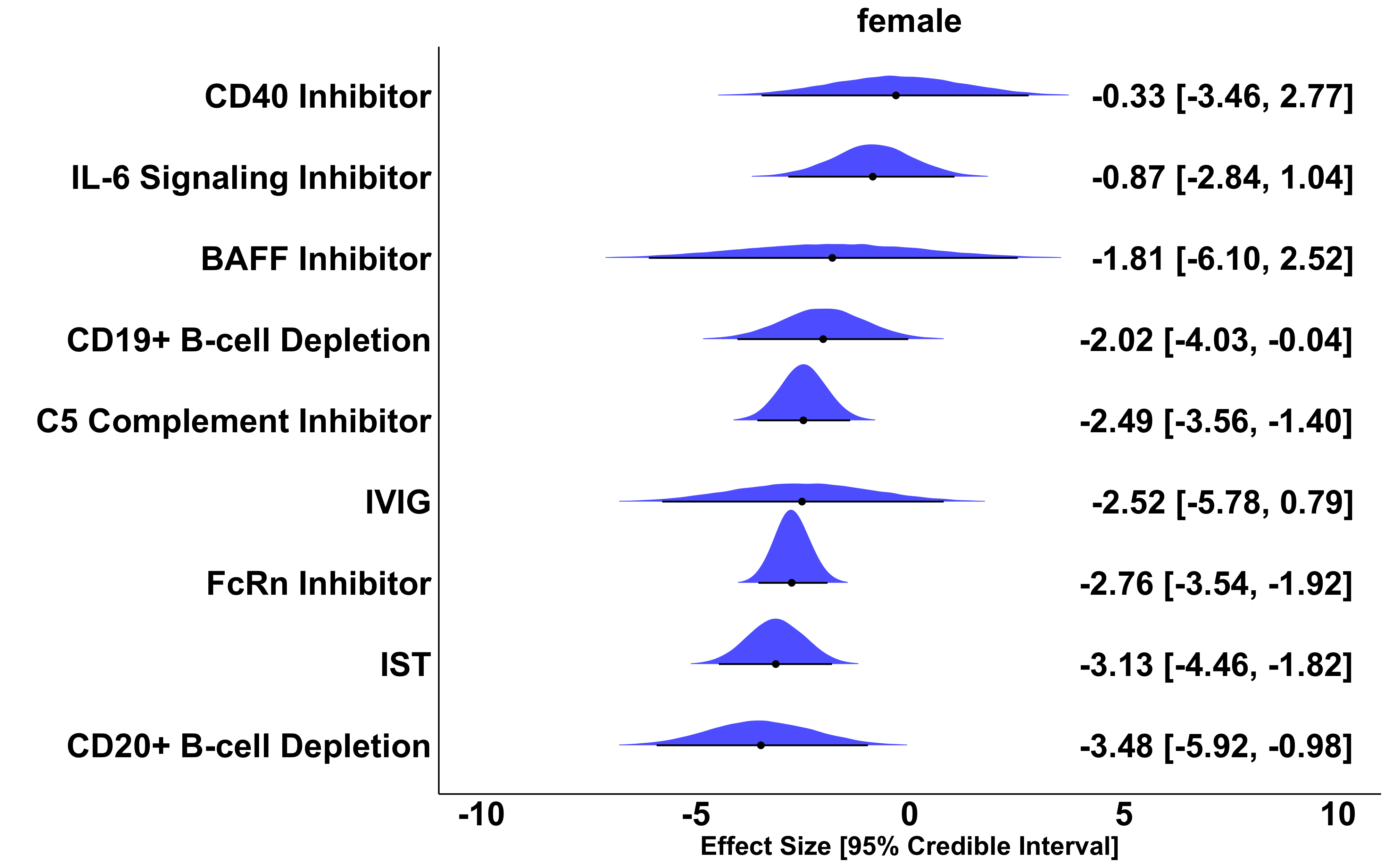

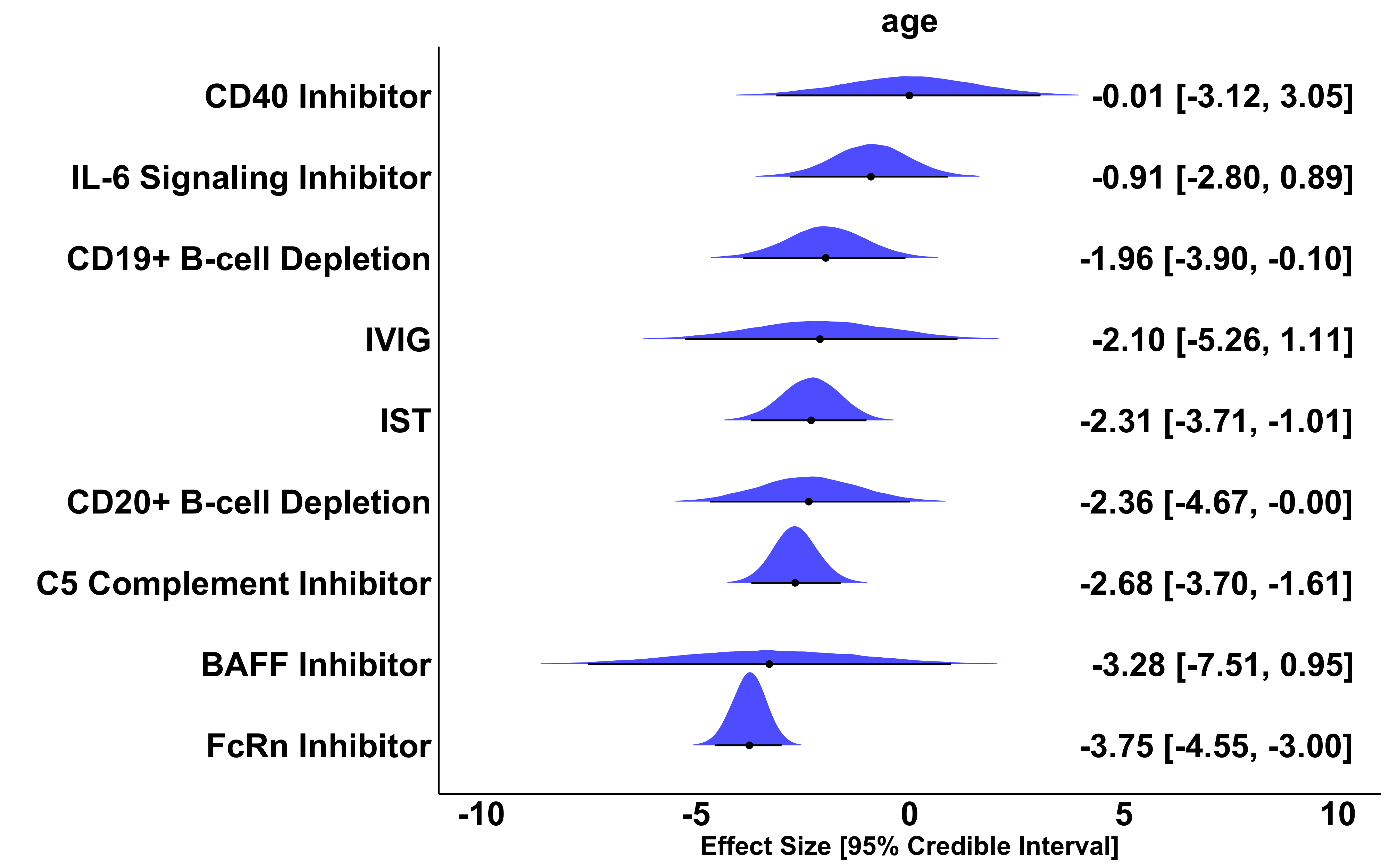

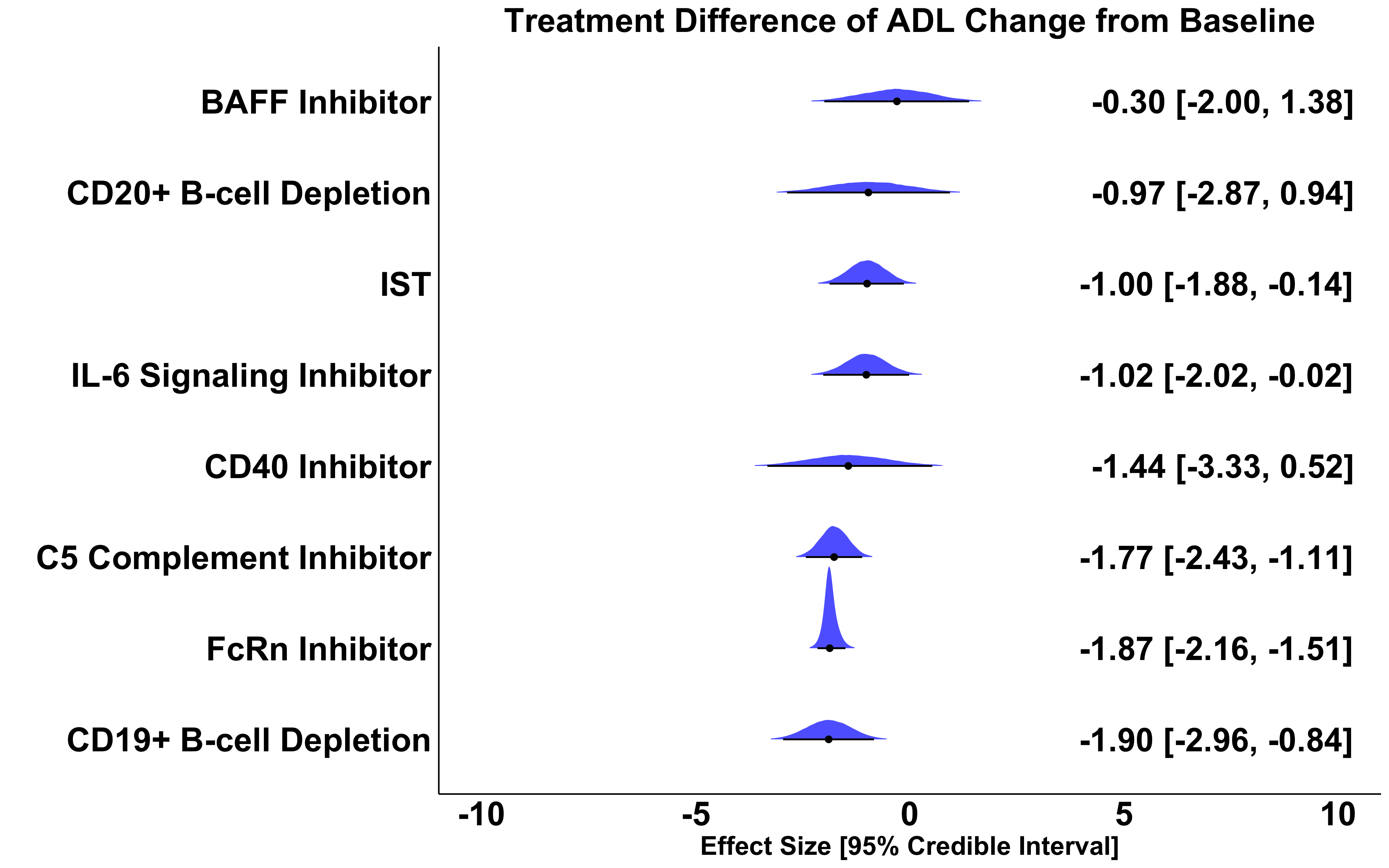

### **eFigure 11.** Effect size, 95% Credible Interval, and overlayed posterior distribution of MG-ADL change from baseline compared to placebo across all 27 placebo-controlled studies excluding phase 2 batoclimab trial. A negative effect size of x is interpreted as: the treatment reduces MG-ADL x points more than placebo and standard of care.

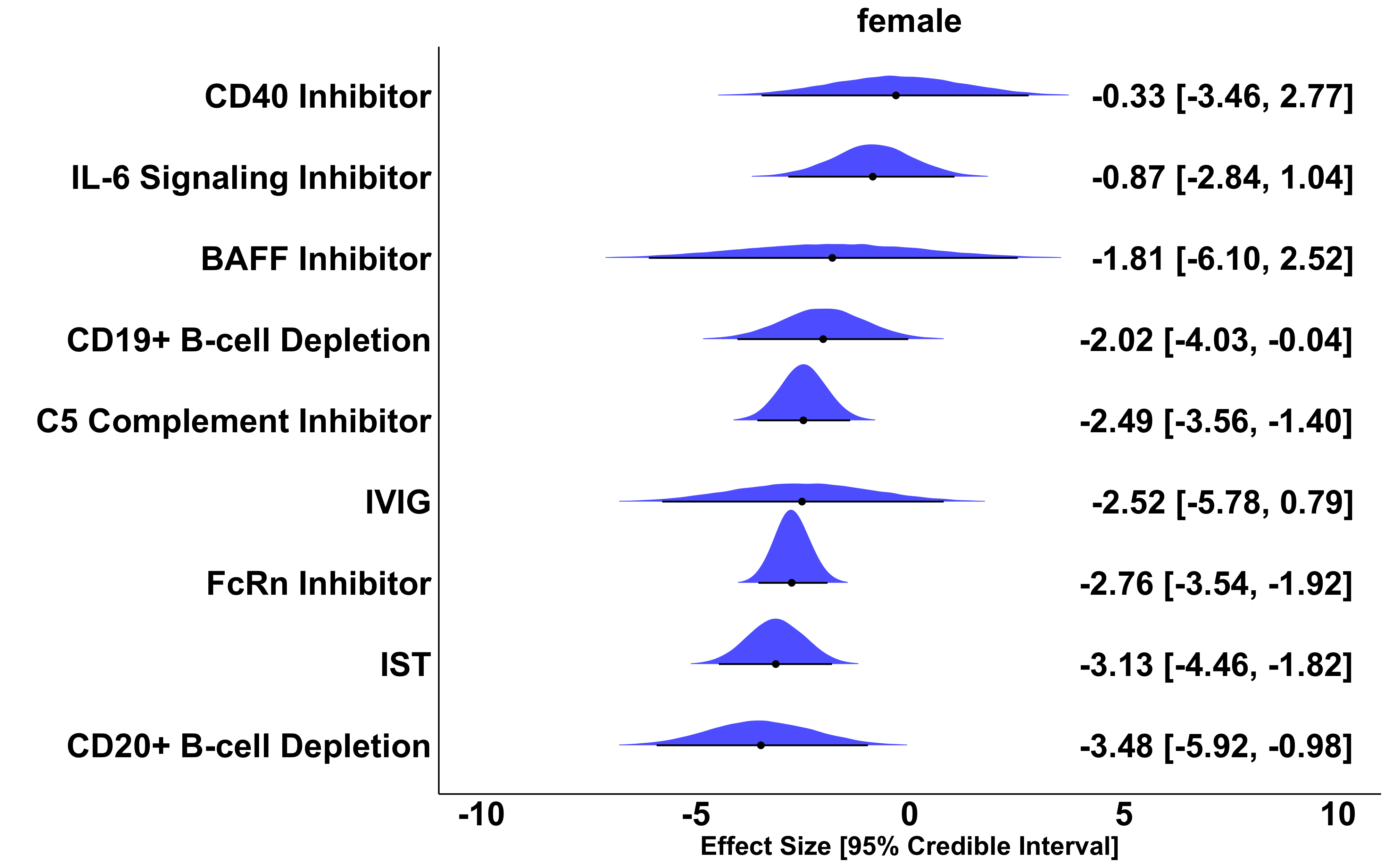

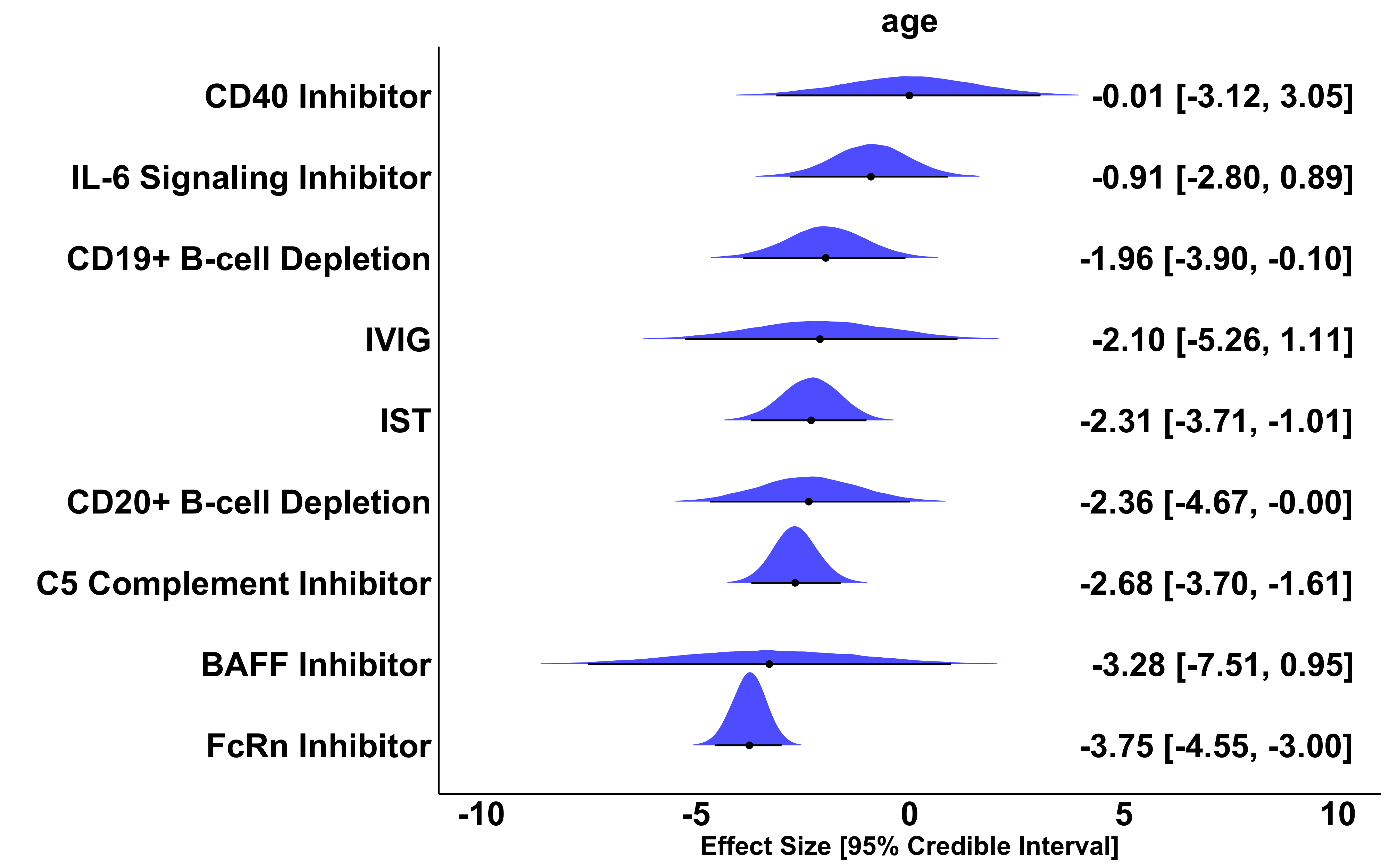

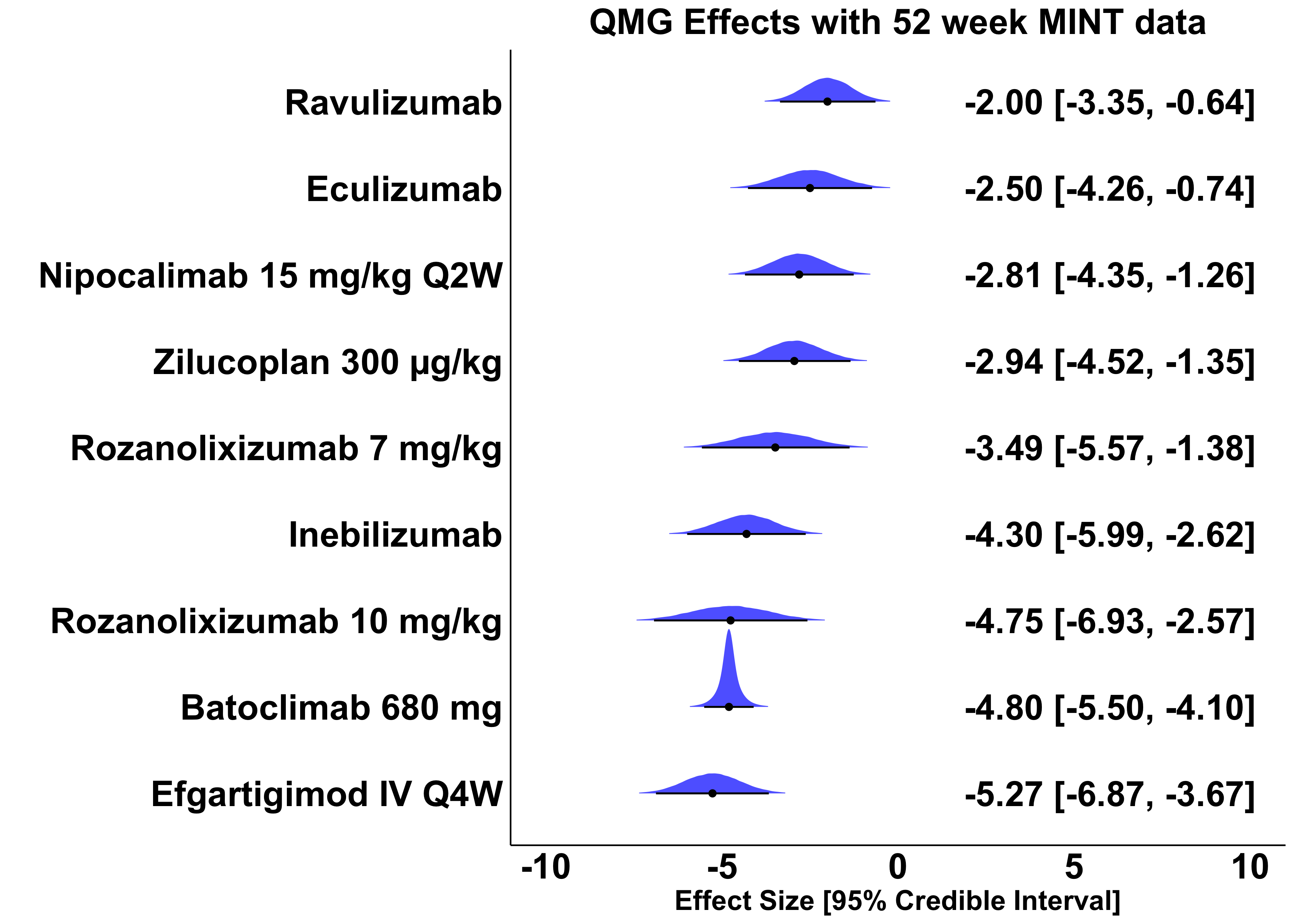
**eFigure 12.**
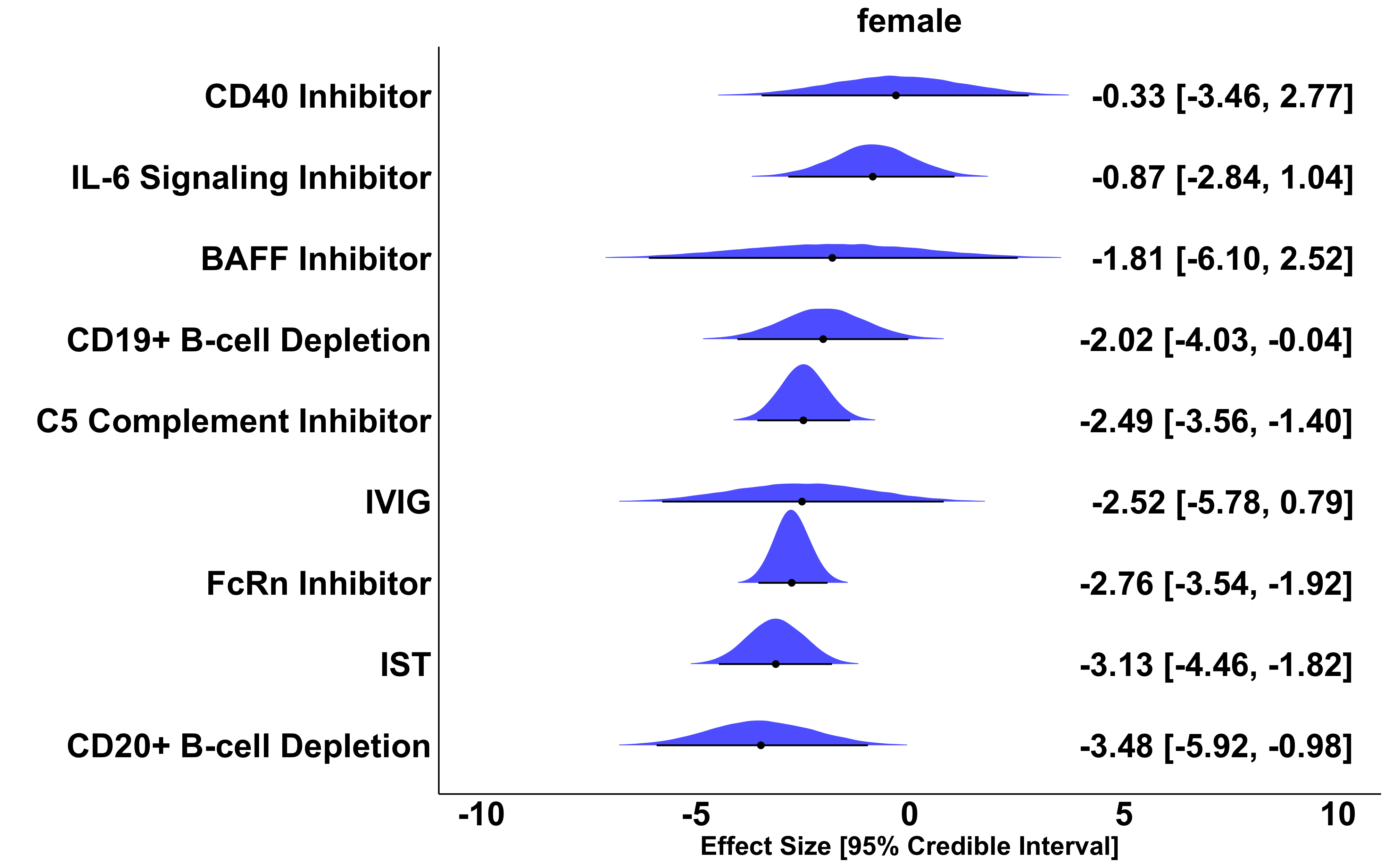

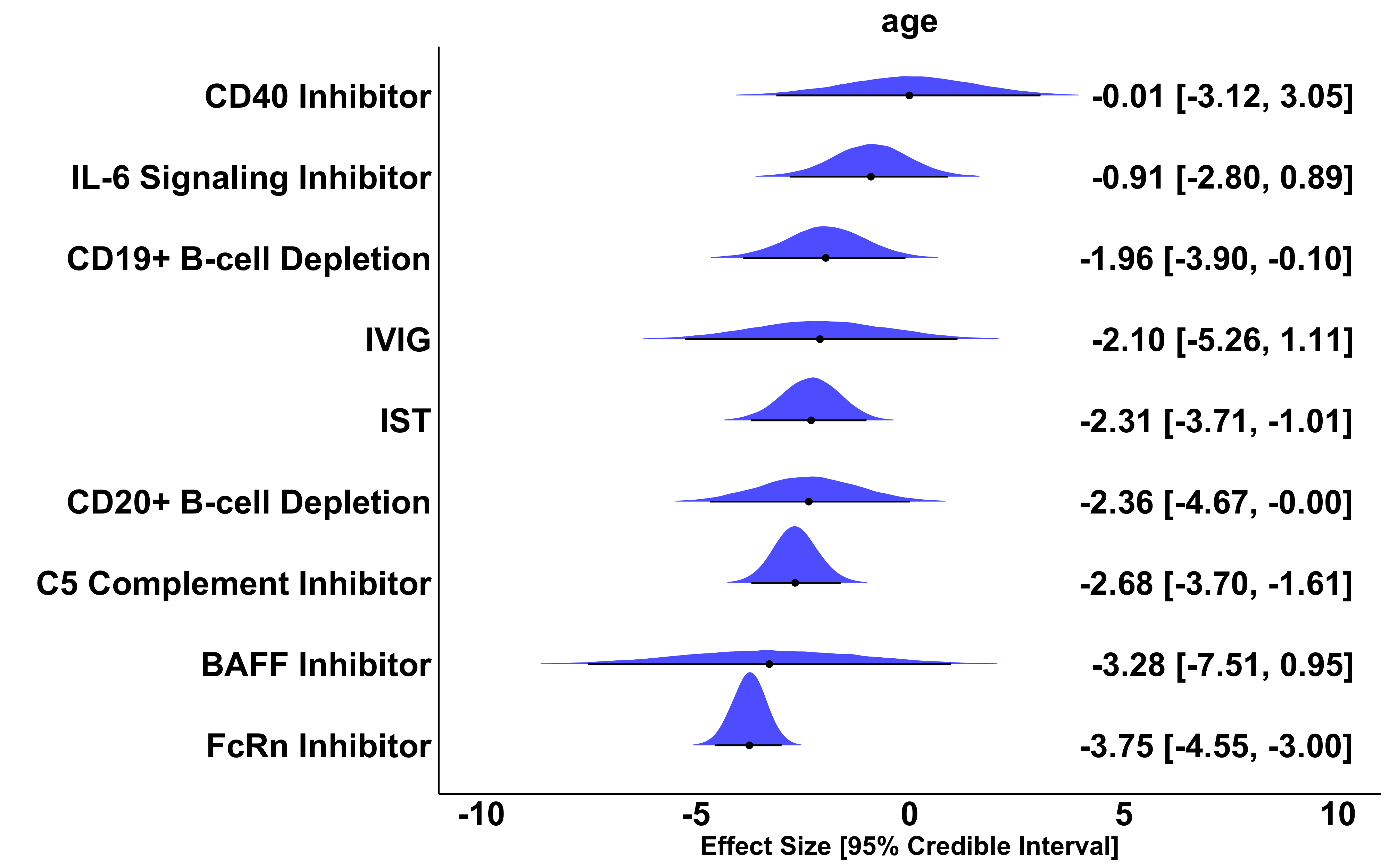
Effect size, 95% Credible Interval, and overlayed posterior distribution of QMG change from baseline compared to placebo across phase 3 trials with 52-week inebilizumab data. A negative effect size of x is interpreted as: the treatment reduces QMG x points more than placebo and standard of care.

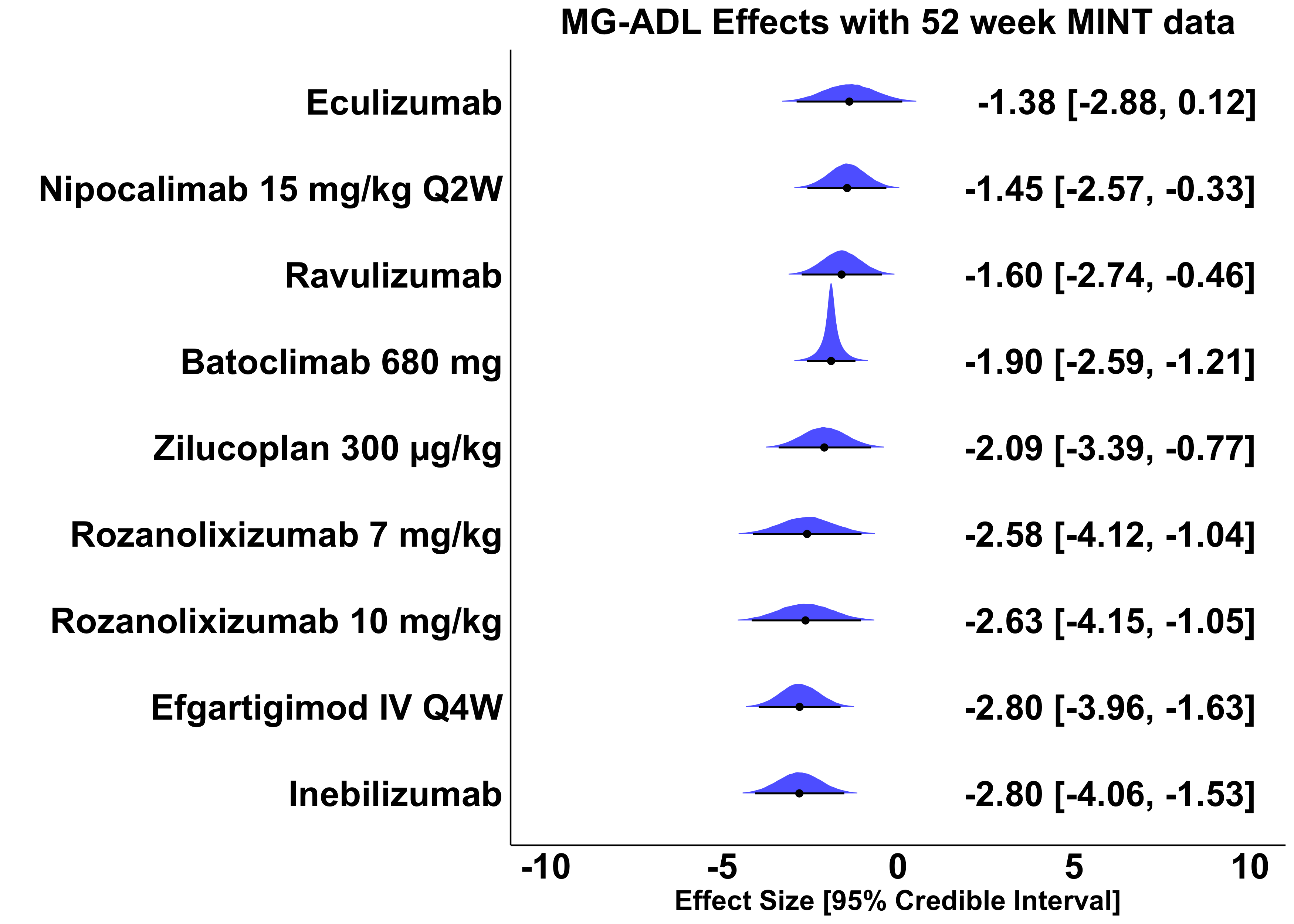

**eFigure 13.**
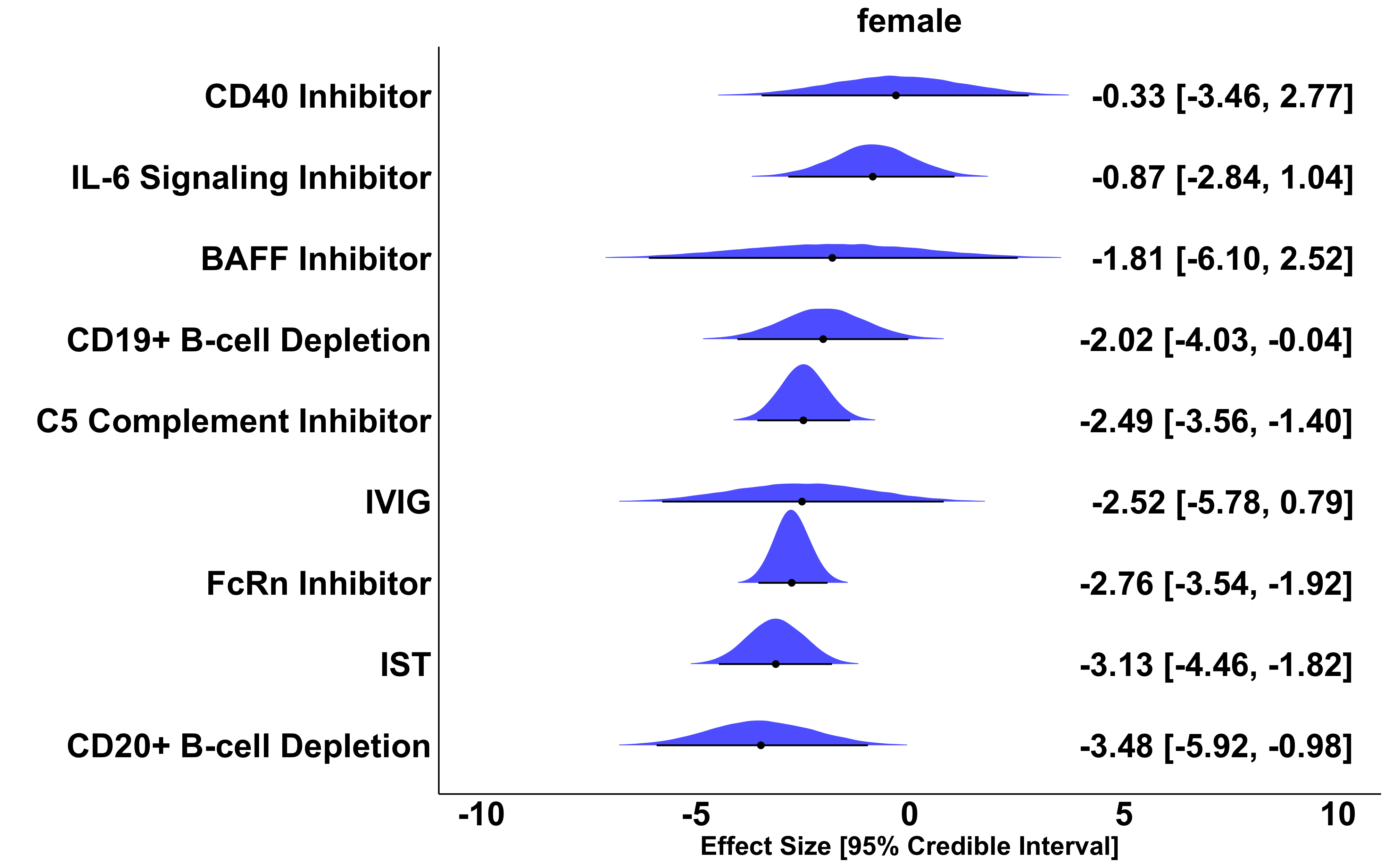

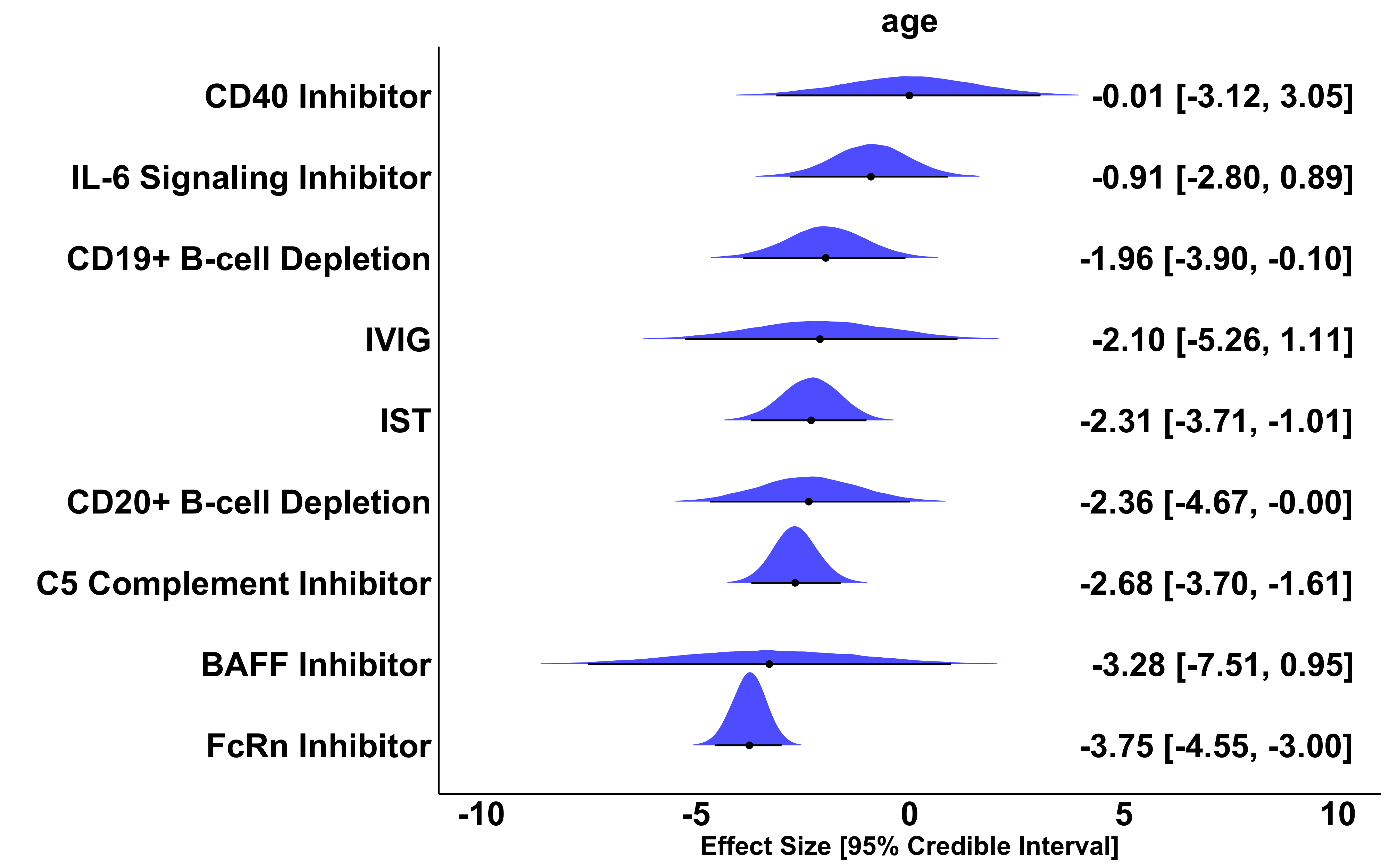
Effect size, 95% Credible Interval, and overlayed posterior distribution of MG-ADL change from baseline compared to placebo across phase 3 trials with 52-week inebilizumab data. A negative effect size of x is interpreted as: the treatment reduces MG-ADL x points more than placebo and standard of care.

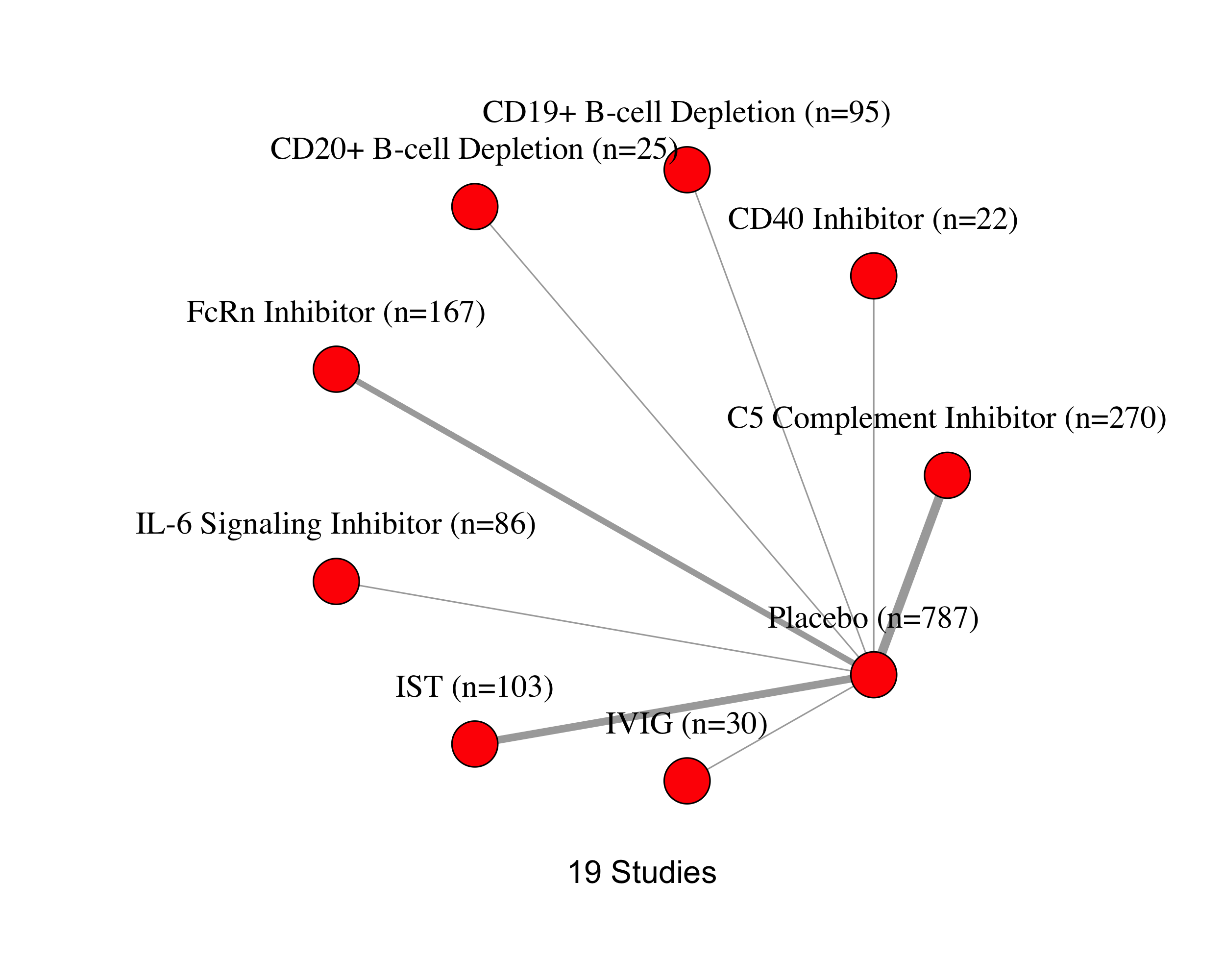

### **eFigure 14.** Network plot of included studies for analysis of QMG treatment effect in AChR+ patients.

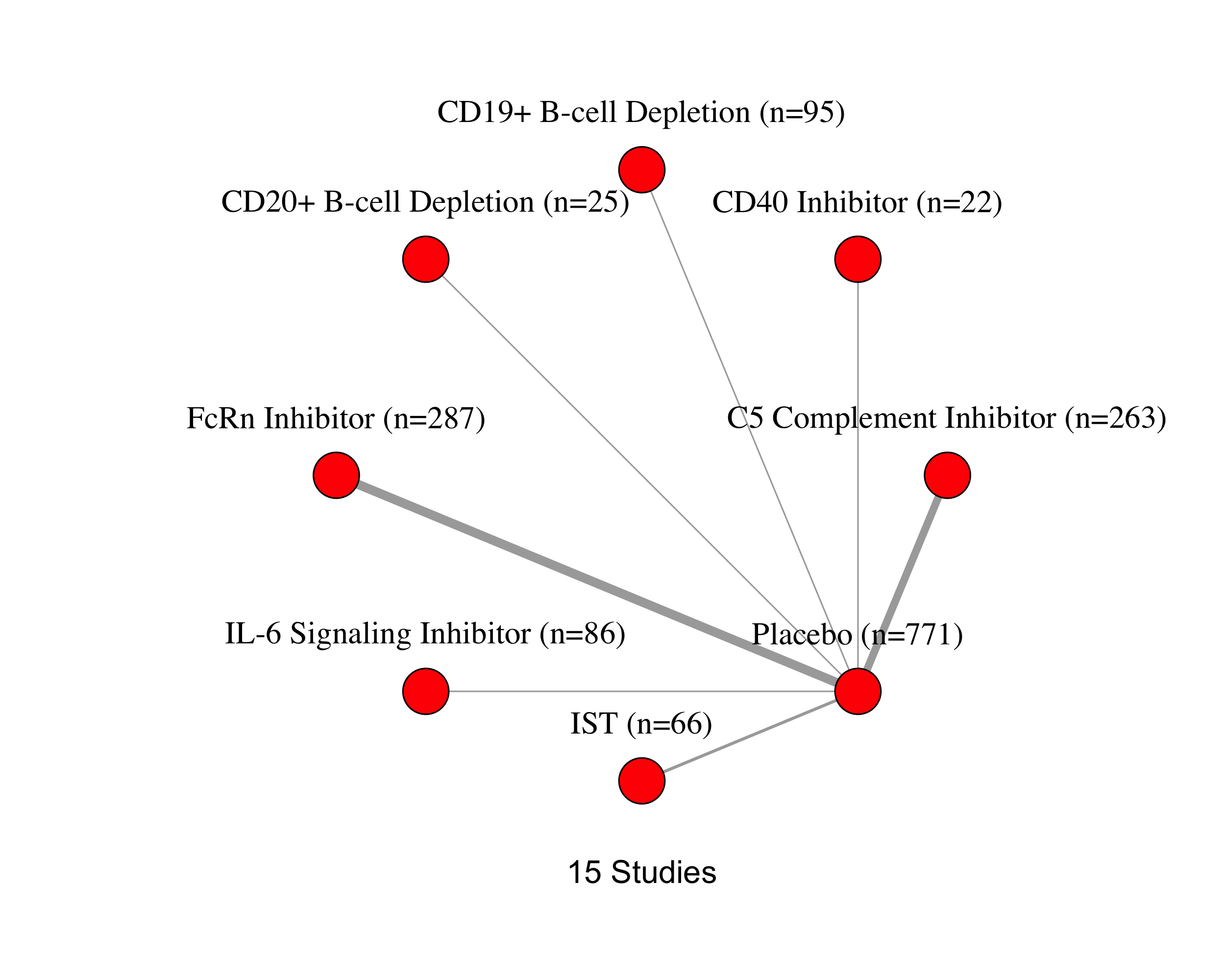

### **eFigure 15.** Network plot of included studies for analysis of MG-ADL treatment effect in AChR+ patients.

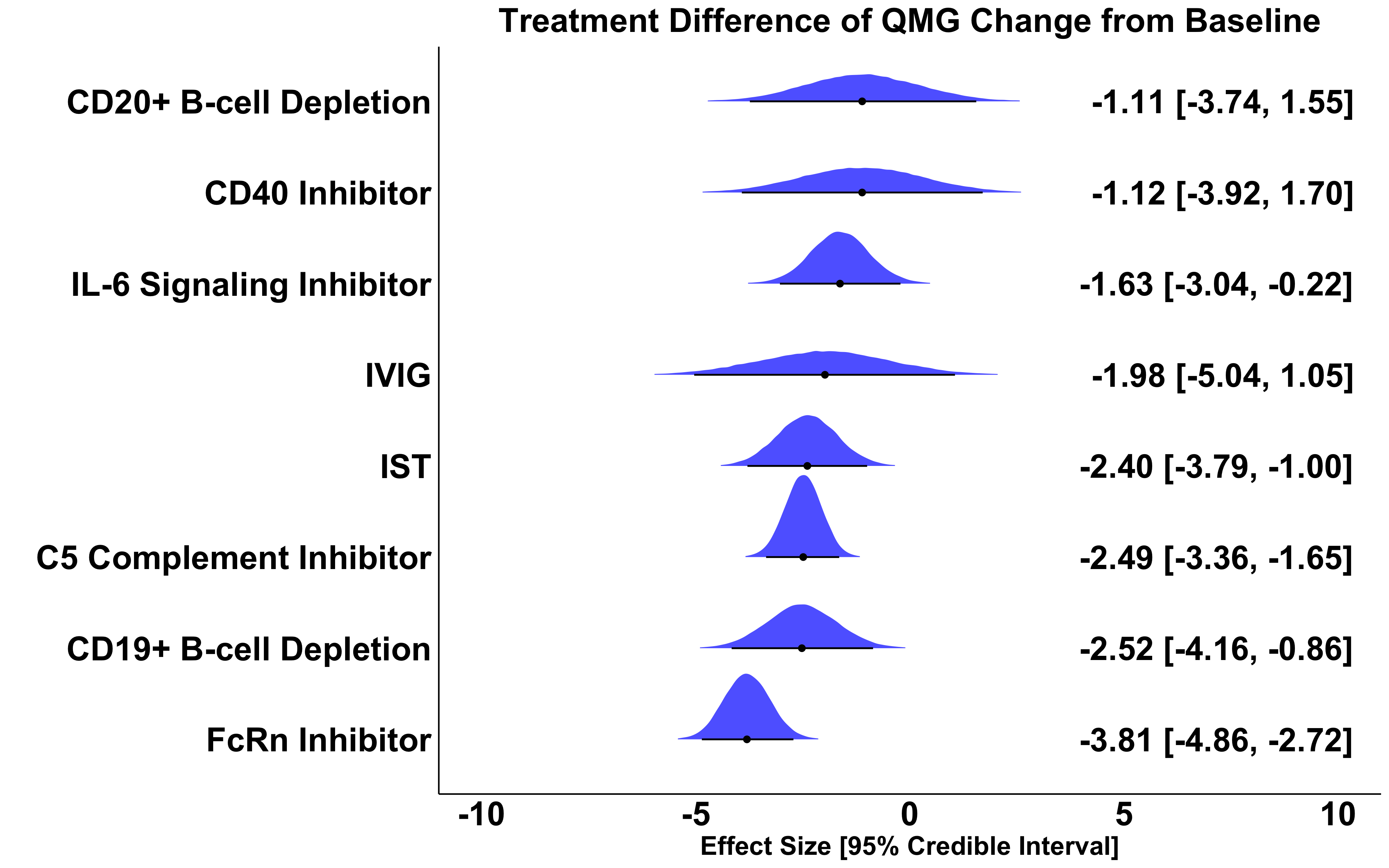

### **eFigure 16.** Effect size, 95% Credible Interval, and overlayed posterior distribution of QMG change from baseline compared to placebo in AChR+ patients. A negative effect size of x is interpreted as: the treatment reduces QMG x points more than placebo and standard of care.

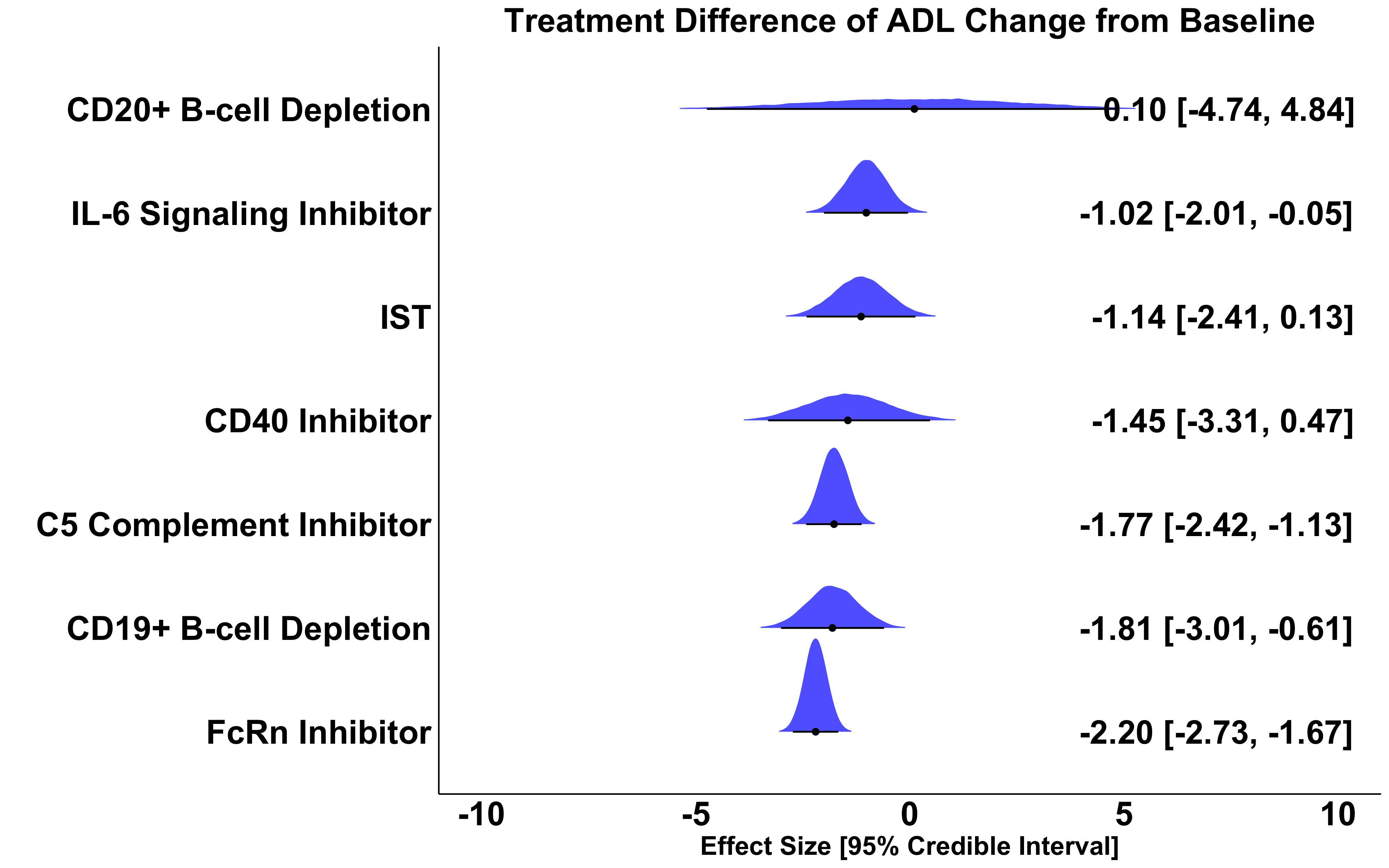

### **eFigure 17.** Effect size, 95% Credible Interval, and overlayed posterior distribution of MG-ADL change from baseline compared to placebo in AChR+ patients. A negative effect size of x is interpreted as: the treatment reduces MG-ADL x points more than placebo and standard of care.

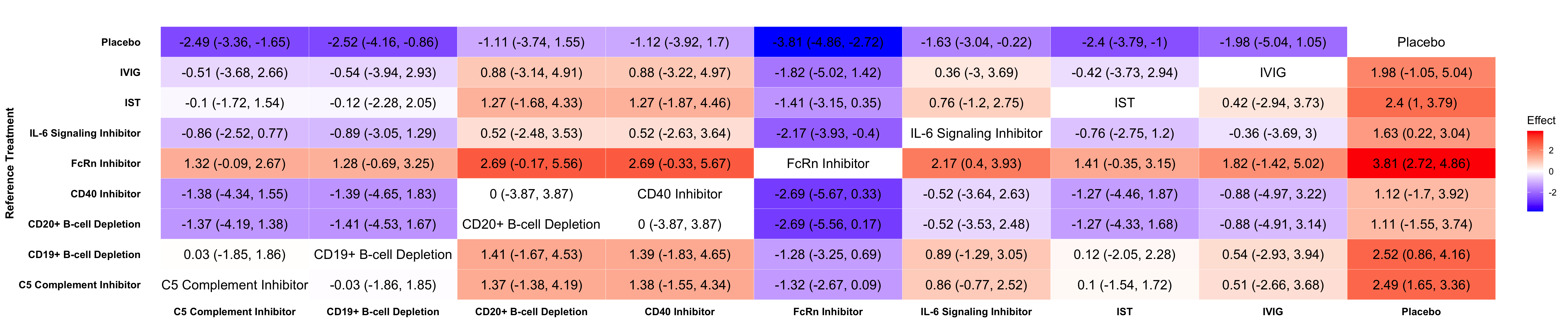

### **eFigure 18.** League table of relative treatment effect of QMG change from baseline across all pairs of treatments in AChR+ patients. Reference treatments are on the rows. A treatment effect of -1 in row A and column B is interpreted as: treatment B reduces the outcome measure by one more point than treatment A.

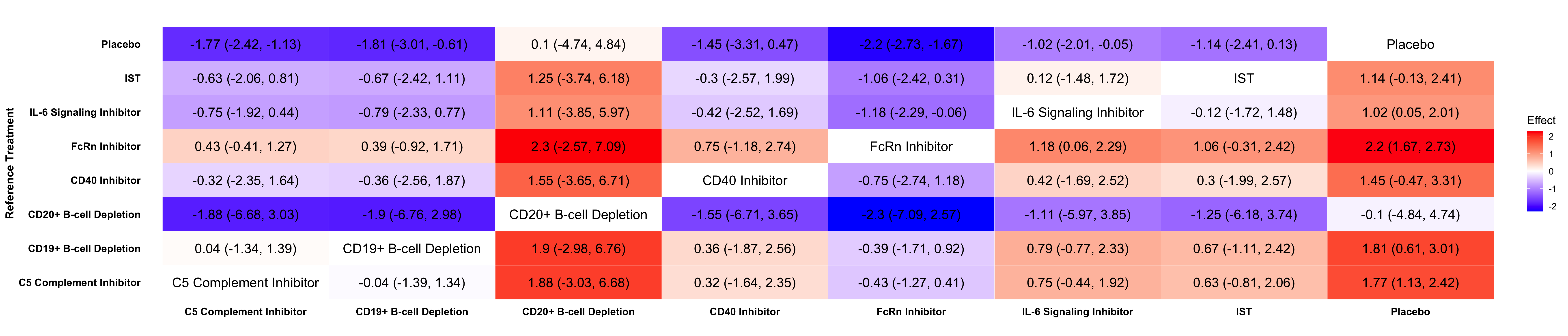

### **eFigure 19.** League table of relative treatment effect of MG-ADL change from baseline across all pairs of treatments in AChR+ patients. Reference treatments are on the rows. A treatment effect of -1 in row A and column B is interpreted as: treatment B reduces the outcome measure by one more point than treatment A.

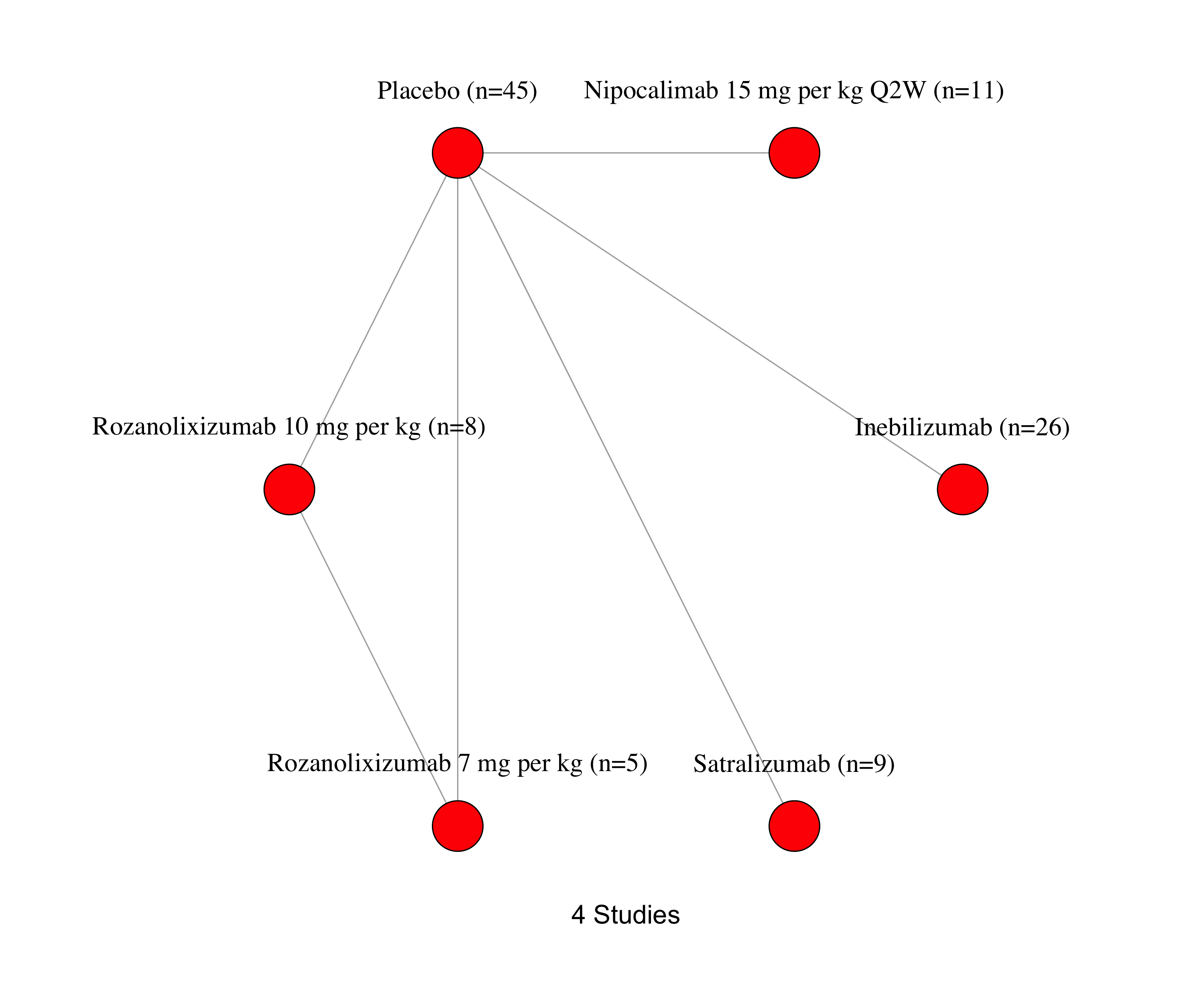

### **eFigure 20.** Network plot of included studies for analysis of MG-ADL treatment effect in MuSK+ patients.

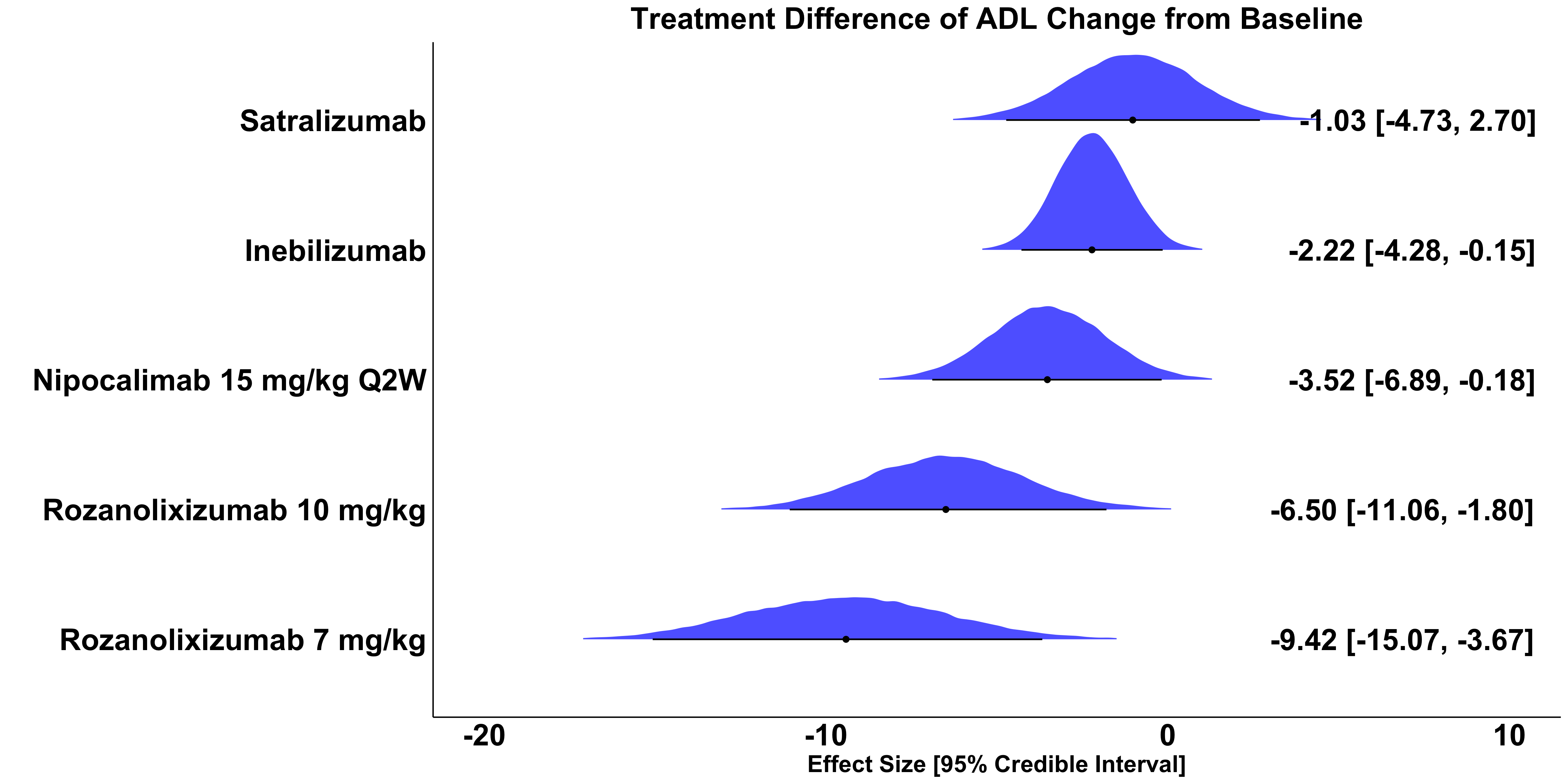

### **eFigure 21.** Effect size, 95% Credible Interval, and overlayed posterior distribution of MG-ADL change from baseline compared to placebo in MuSK+ patients. A negative effect size of x is interpreted as: the treatment reduces MG-ADL x points more than placebo and standard of care.

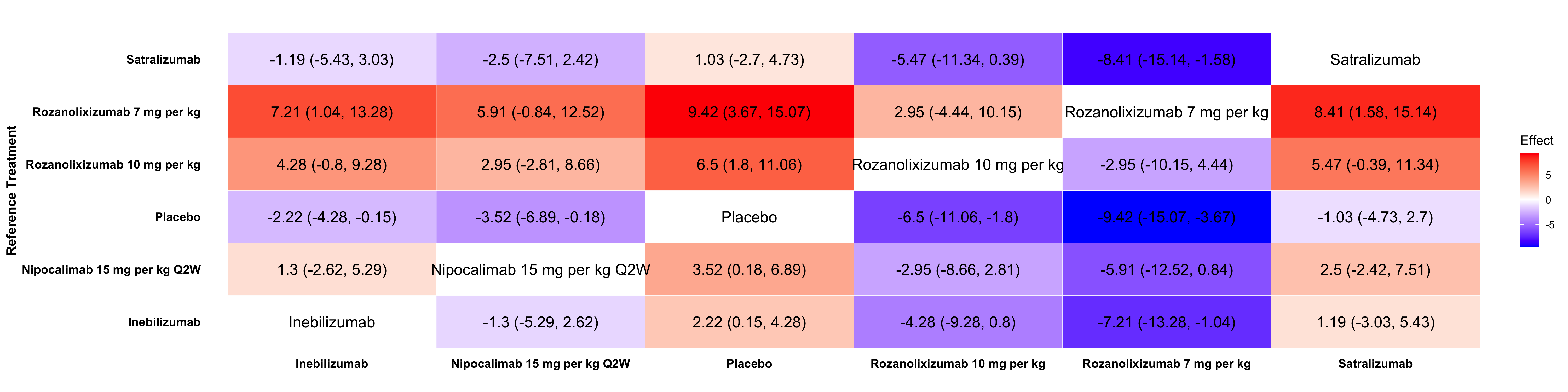

### **eFigure 22.** League table of relative treatment effect of MG-ADL change from baseline across all pairs of treatments in MuSK+ patients. Reference treatments are on the rows. A treatment effect of -1 in row A and column B is interpreted as: treatment B reduces the outcome measure by one more point than treatment A.

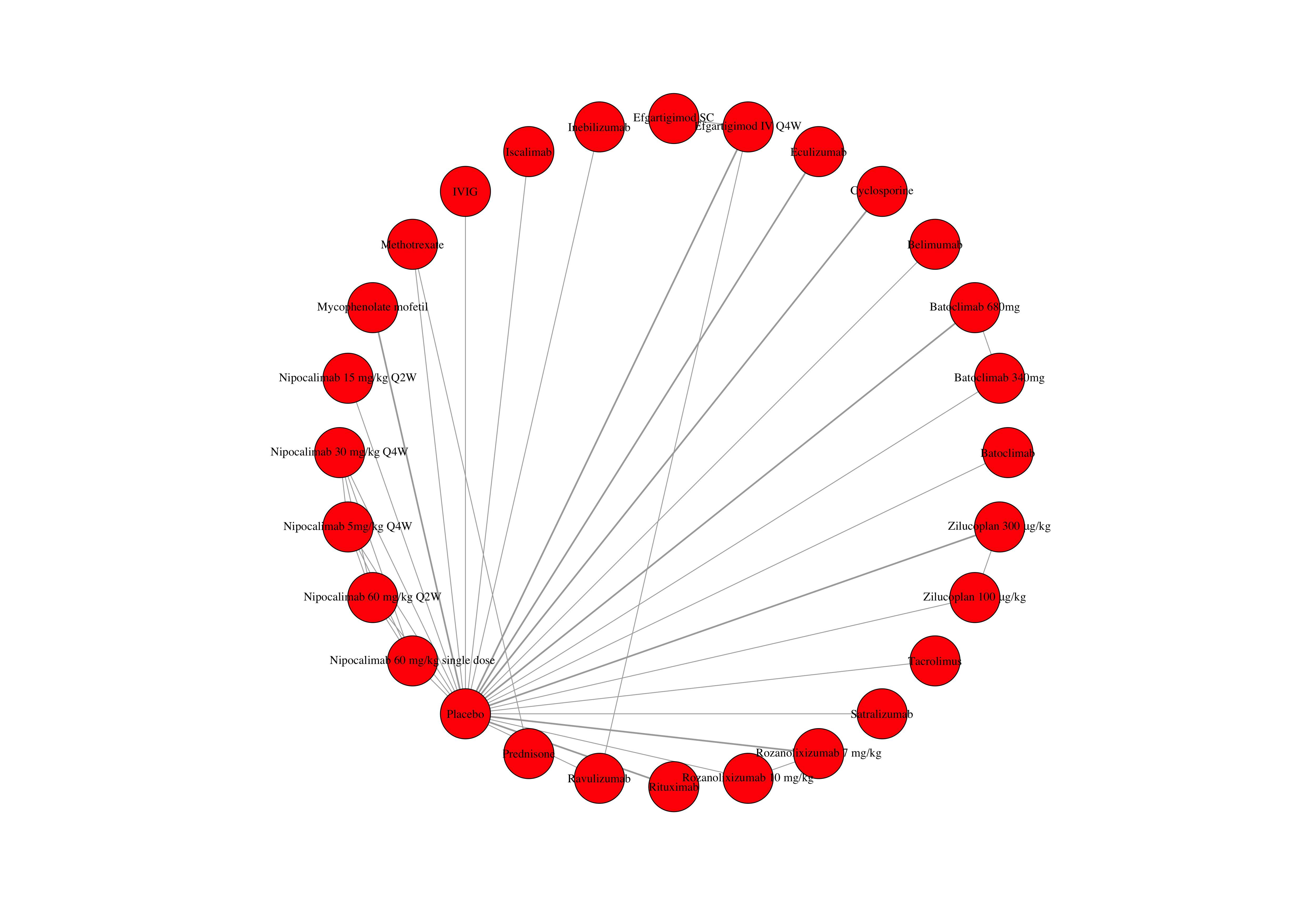

### **eFigure 23.** Network plot of included individual dosages for analysis of QMG treatment effect.

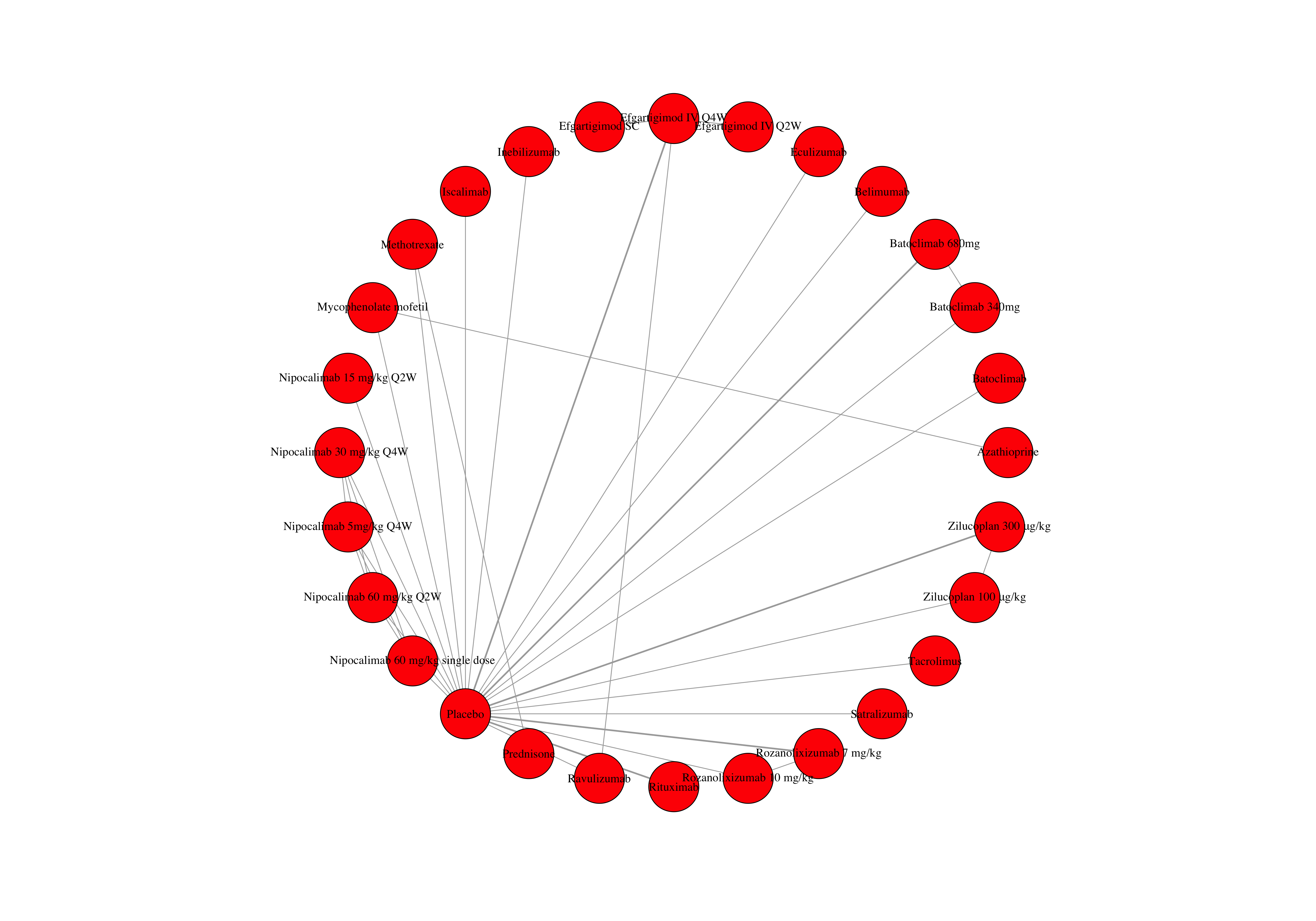

### **eFigure 24.** Network plot of included individual dosages for analysis of MG-ADL treatment effect.

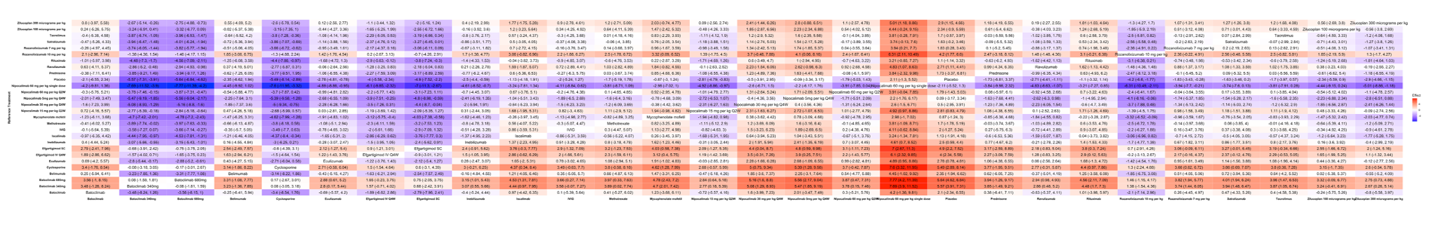

### **eFigure 25.** League table of relative treatment effect of QMG change from baseline across all pairs of treatments dosages. Reference treatments are on the rows. A treatment effect of -1 in row A and column B is interpreted as: treatment B reduces the outcome measure by one more point than treatment A.

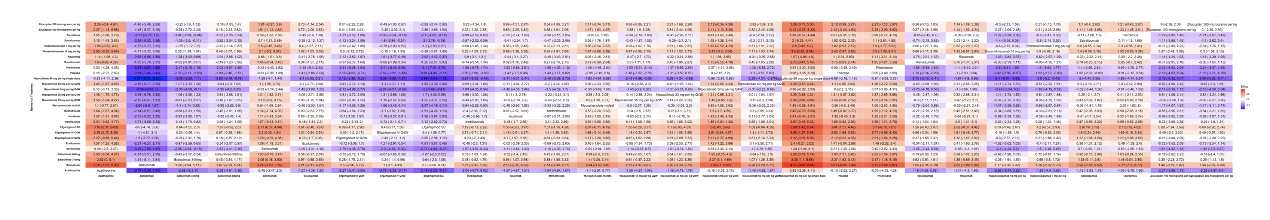

### **eFigure 26.** League table of relative treatment effect of QMG change from baseline across all pairs of treatments dosages. Reference treatments are on the rows. A treatment effect of -1 in row A and column B is interpreted as: treatment B reduces the outcome measure by one more point than treatment A.

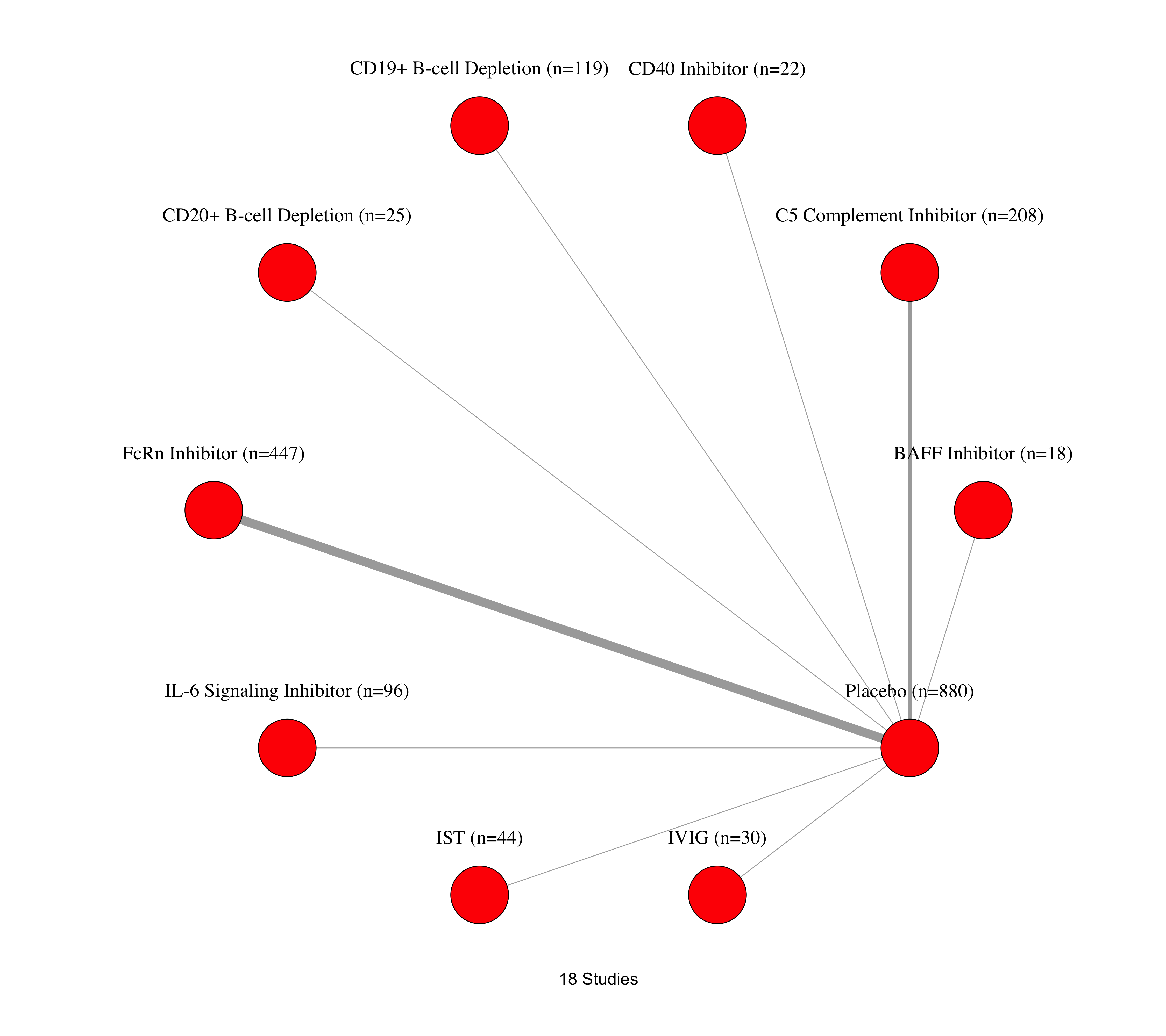

### **eFigure 27.** Network plot of included studies for analysis of treatment-related adverse events.

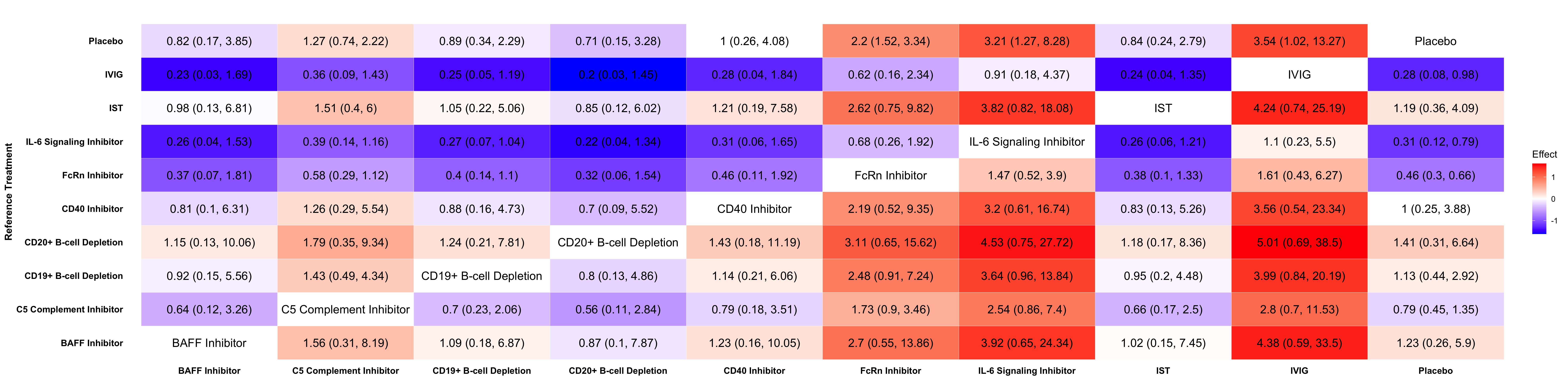

### **eFigure 28.** League table of relative odds of treatment-related adverse events. Each cell in the table is the odds of an adverse event from the treatment in the column compared to the treatment in the row. An odds ratio of 2 in row A and column B is interpreted as: patients treated with treatment B have two times increased odds of a treatment-related adverse event compared to patients treated with treatment A.

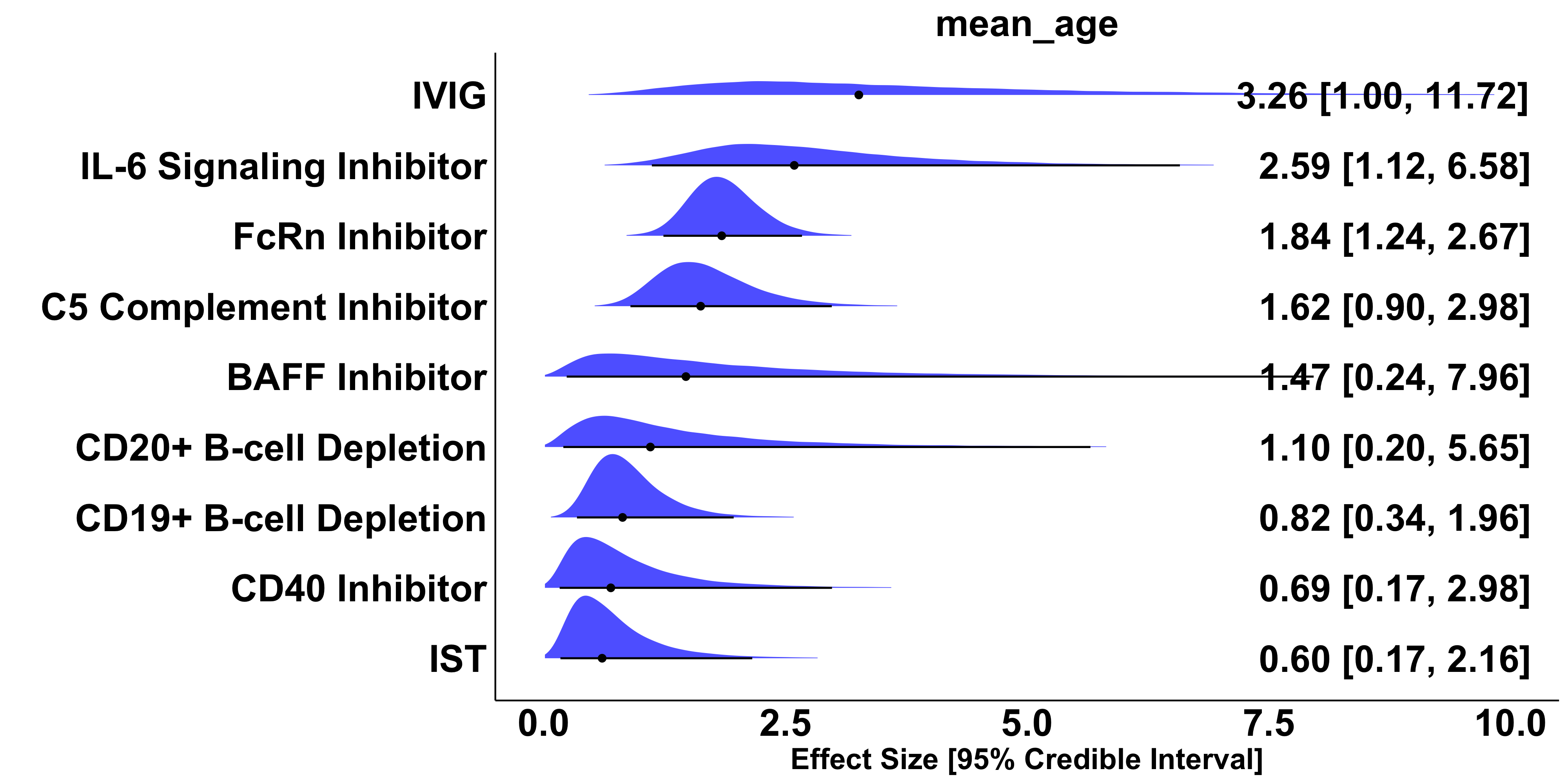

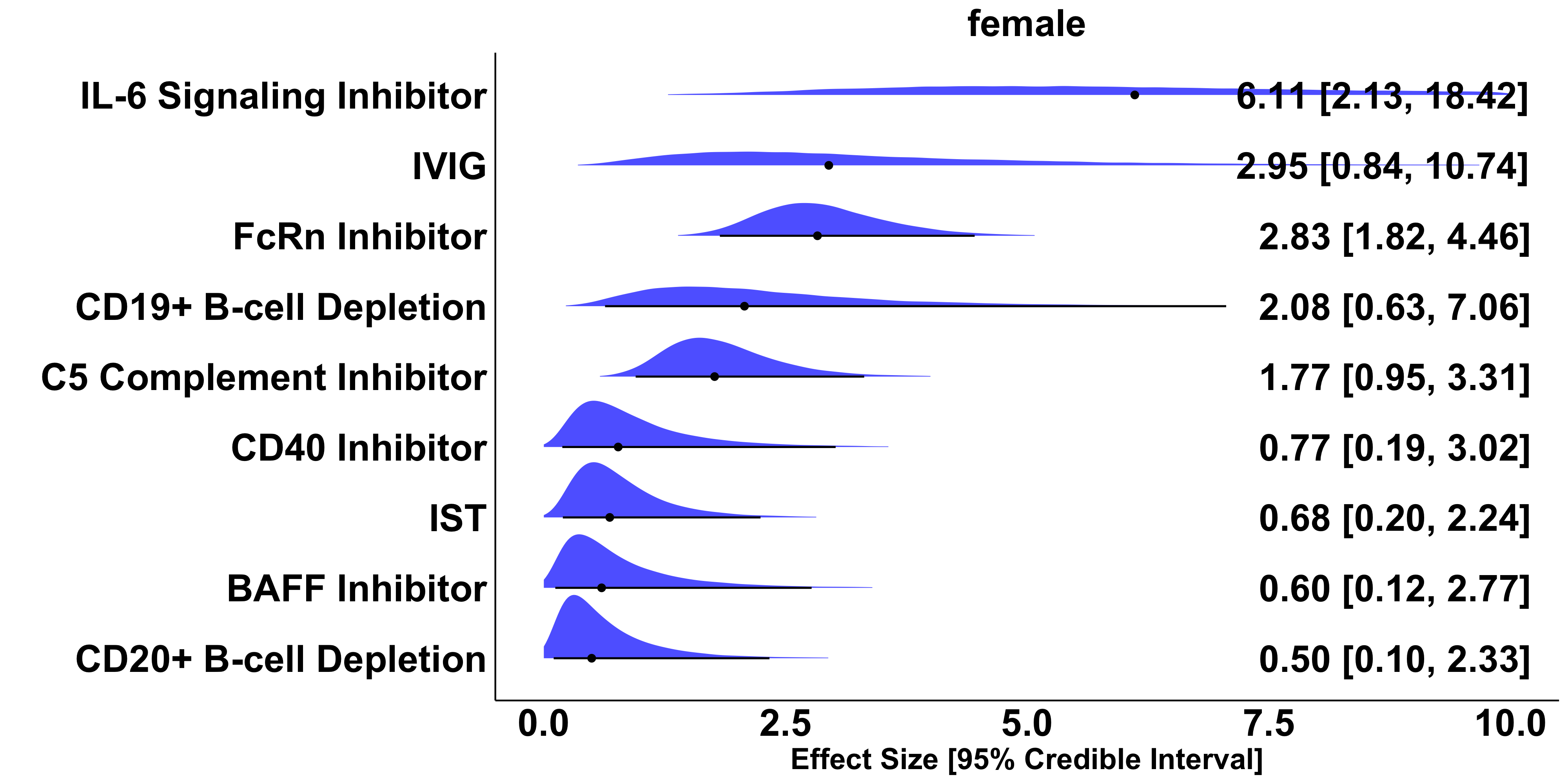

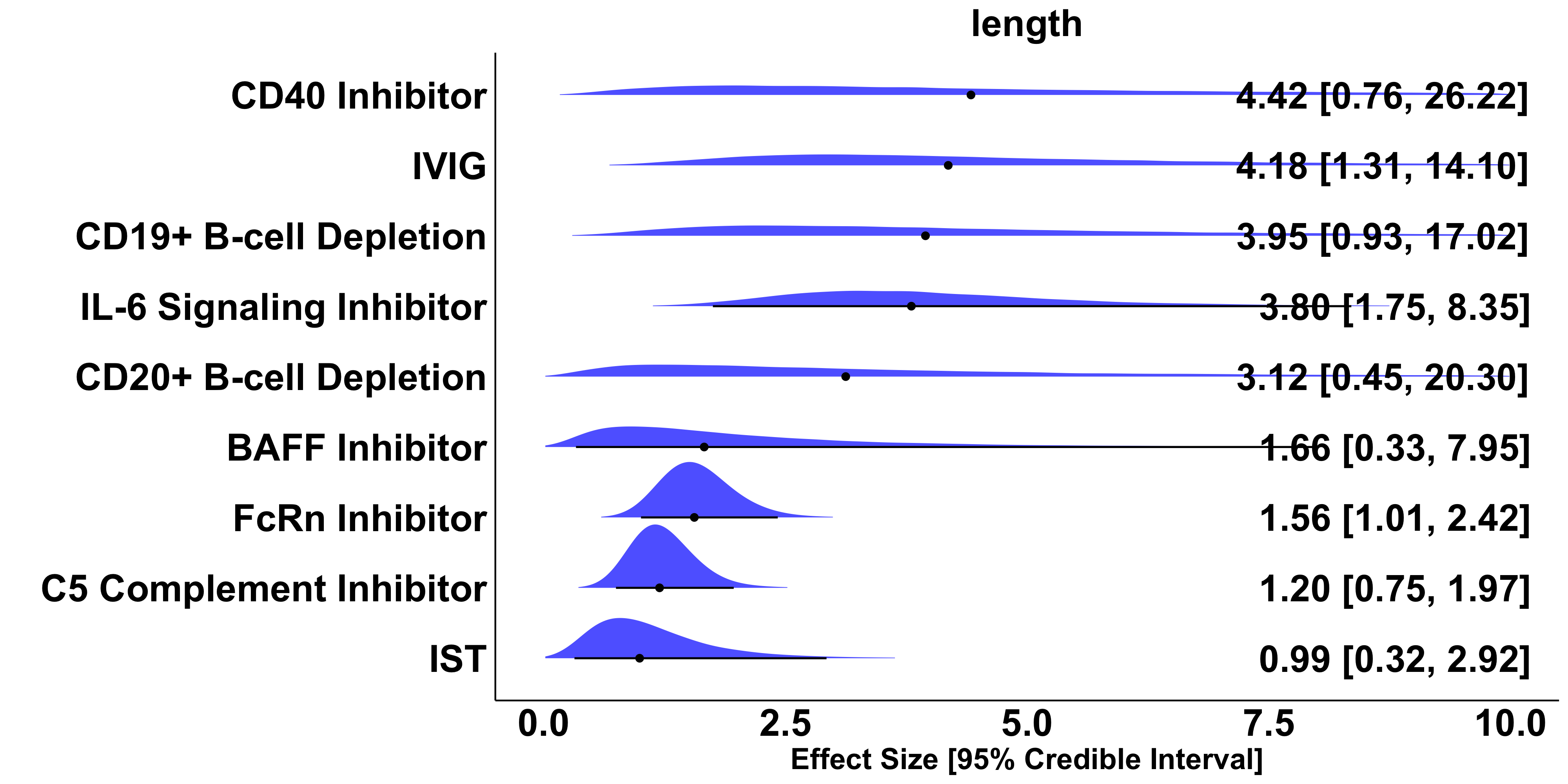

### **eFigure 29.** Effect size, 95% Credible Interval, and overlayed posterior distribution of odds of treatment-related adverse events compared to placebo. Odds ratios adjusted for study-wide age at enrollment, percentage of female participants, and study length in weeks. All adjusted effect sizes represent the treatment effect if all studies had the same value of the covariate. An odds ratio greater than 1 is interpreted as: patients have greater odds of treatment-related adverse events compared to placebo.
